# Towards Equitable Patient Subgroup Performance by Gene-Expression-Based Diagnostic Classifiers of Acute Infection

**DOI:** 10.1101/2022.04.24.22274125

**Authors:** Michael B. Mayhew, Uros Midic, Kirindi Choi, Purvesh Khatri, Ljubomir Buturovic, Timothy E. Sweeney

## Abstract

Host-response gene expression measurements may carry confounding associations with patient demographic characteristics that can induce bias in downstream classifiers. Assessment of deployed machine learning systems in other domains has revealed the presence of such biases and exposed the potential of these systems to cause harm. Such an assessment of a gene-expression-based classifier has not been carried out and collation of requisite patient subgroup data has not been undertaken. Here, we present data resources and an auditing framework for patient subgroup analysis of diagnostic classifiers of acute infection. Our dataset comprises demographic characteristics of nearly 6500 patients across 49 studies. We leverage these data to detect differences across patient subgroups in terms of gene-expression-based host response and performance with both our candidate pre-market diagnostic classifier and a standard-of-care biomarker of acute infection. We find evidence of variable representation with respect to patient covariates in our multi-cohort datasets as well as differences in host-response marker expression across patient subgroups. We also detect differences in performance of multiple host-response-based diagnostics for acute infection. This analysis marks an important first step in our ongoing efforts to characterize and mitigate potential bias in machine learning-based host-response diagnostics, highlighting the importance of accounting for such bias in developing diagnostic tests that generalize well across diverse patient populations.

## 1 Introduction

Detecting acute infection and sepsis is a challenging task with considerable impact on patient morbidity and mortality as well as healthcare utilization and expenditures [12]. Conventional diagnosis of acute infection can be slow and non-specific, relying on lab cultures to detect the presence of particular pathogens to guide subsequent treatment [36]. Host response diagnostics offer a potentially more timely and accurate means of performing this task, leveraging measurements of targeted biomarkers derived from patient blood to detect the presence and type of infection. For example, biomarkers such as procalcitonin (PCT) have, despite potential shortcomings in terms of clinical utility [18], been developed to detect and manage infection as well as predict adverse events such as patient mortality [13].

Recent studies have also demonstrated the potential of measuring expression of targeted mRNA transcripts from patient blood to diagnose a range of infectious diseases [38, 42, 1, 2]. Development of these host response signatures requires first identifying those genes strongly associated with a condition of interest in one or multiple studies. The latter, multi-cohort approach can enable discovery of generalizable signatures by including more samples from the target patient population across a wider range of geographies, healthcare delivery settings and infectious diseases.

However, in either the single- or multi-cohort case, characteristics of samples in the dataset can introduce or exacerbate confounding associations between patient variables (e.g. demographics) and outcomes of the classification task. Such variations in biomarker expression due to patient attributes may produce differences in a ML classifier’s performance with respect to those attributes. Thus, prior to clinical deployment, these diagnostic classifiers should ideally demonstrate high performance not only in a single mixed-population validation dataset but also across different patient subgroups. Characterizing these confounding effects is made more challenging by the requisite task of compiling information on patient attributes across a number of studies [5]. Indeed, a recent working group organized by the US Food & Drug Administration highlighted the need to enhance demographic data collection and communication efforts to enable such analyses [9].

Analyses of deployed ML systems for healthcare [4, 26, 5] as well as other domains [6, 3, 4] have exposed potential biases, revealing variations in system performance that correlate with demographic attributes of subjects in the dataset. These biases have highlighted the need for end-to-end assessments in the development and deployment of ML systems [32, 40] in order to mitigate potential harms and performance disparities across patient subgroups. These findings in the ML community dovetail with decades of research in population genetics, epidemiology and clinical science on the estimation of and accounting for sources of confounding or bias in statistical models of patient characteristics (e.g. [10, 35, 39]). As the use of ML becomes more prevalent in molecular diagnostics [7], the need for such auditing pipelines in molecular diagnostic classifier development is clear and, currently, unmet.

Here, we evaluate a pre-market, gene expression-based diagnostic classifier of acute infection across different patient sub-groups. To this end, we compile demographic patient attributes across 49 studies of nearly 6500 patients, many of which with clinically-adjudicated bacterial infection, viral infection or non-infectious inflammation. We provide these data to the research community as a resource for further diagnostic classifier development efforts (see Supplementary Material). In a similar vein as subgroup analyses of treatment effects [8], we develop Bayesian multi-level models to detect associations between patient demographic attributes, infection status and the expression of 29 host mRNAs previously selected for development of diagnostic classifiers of acute infection. In addition, drawing inspiration from recent work in the ML community on algorithmic bias, we evaluate the performance of previously developed diagnostic classifiers of acute infection across patient subgroups in a multi-cohort validation dataset. Our study highlights the important and currently under-appreciated role of subgroup analysis in auditing ML-based diagnostic systems.

## 2 Methods

### 2.1 Description of multi-cohort gene expression data

We initially identified 3159 samples from 42 studies that profiled whole blood of patients with acute infection or sepsis in the NCBI GEO and EBI ArrayExpress repositories (as described in [24]). Gene expression was profiled using a variety of technical platforms (primarily microarray). Studies were conducted in different geographic regions and with both adult and pediatric patients. These publicly available studies were used for diagnostic classifier training. For the purposes of external validation, we also profiled 931 patients across 15 studies using the NanoString SPRINT platform.

We collected ground truth labels of a patient’s infection status (**B** - bacterial infection, **V** - viral infection, **N** - non-infectious inflammation) for all samples. A breakdown of all samples across different patient characteristics appears in Table 1 while the infection status breakdown of our publicly available and NanoString-profiled studies appears in Supplementary Tables 1 and 2, respectively. Depending on available data, a patient’s infection status was determined differently for the training and validation studies. For training studies, we used the labels provided by each study. Derivation of these labels involved multi-clinician adjudication with or without positive pathogen identification or positive pathogen identification alone. In the event that adjudications were not directly provided by the study, we used pathogen test results from the study’s metadata/manuscripts to assign an appropriate class label. Validation study labels of infection status were assigned using pathogen test results alone or in combination with adjudication by a multi-clinician panel using all available clinical data. We did not include healthy controls in either our training or validation datasets.

**Table 1:** Composition of datasets of acute infection patients.

|  | Overall<br>(N=3533) | Bacterial<br>Infection (N=1388) | Viral<br>Infection (N=1218) | Non-infectious<br>Inflammation (N=947) |
| --- | --- | --- | --- | --- |
| **Female** | 1604 (45.1%) | 626 (45.1%) | 511 (42.0%) | 467 (49.3%) |
| **Median age in years (range)** | 26 (0-100) | 52 (0-99) | 17 (0-100) | 20 (0.2-92) |
| **Age Bracket** |  |  |  |  |
| Child (<12yo) | 1257 (35.4%) | 394 (28.4%) | 526 (43.2%) | 337 (35.6%) |
| Young Adult ($\geq 12$yo, <40yo) | 831 (23.4%) | 155 (11.2%) | 385 (31.6%) | 291 (30.7%) |
| Middle-Aged ($\geq 40$yo, <65yo) | 717 (20.2%) | 335 (24.1%) | 172 (14.1%) | 210 (22.2%) |
| Senior ($\geq 65$yo) | 748 (21.1%) | 504 (36.3%) | 135 (11.1%) | 109 (11.5%) |
| **Race/Ethnicity** |  |  |  |  |
| Asian | 449 (12.6%) | 82 (5.9%) | 309 (25.4%) | 58 (6.1%) |
| Black or African descent | 316 (8.9%) | 113 (8.1%) | 113 (9.3%) | 90 (9.5%) |
| Hispanic | 423 (11.9%) | 125 (9.0%) | 110 (9.0%) | 188 (19.9%) |
| White | 1724 (48.5%) | 873 (62.9%) | 462 (37.9%) | 389 (41.1%) |
| Other | 165 (4.6%) | 37 (2.7%) | 50 (4.1%) | 78 (8.2%) |
| Unknown | 476 (13.4%) | 158 (11.4%) | 174 (14.3%) | 144 (15.2%) |
| **Geographic Region** |  |  |  |  |
| Australia & New Zealand | 36 (1.0%) | 16 (1.2%) | 8 (0.7%) | 12 (1.3%) |
| Northern America | 1667 (46.9%) | 565 (40.7%) | 540 (44.3%) | 562 (59.3%) |
| Northern Europe | 529 (14.9%) | 329 (23.7%) | 199 (16.3%) | 1 (0.1%) |
| Southeastern Asia | 322 (9.1%) | 60 (4.3%) | 262 (21.5%) | 0 (0%) |
| Southern Asia | 27 (0.8%) | 0 (0%) | 27 (2.2%) | 0 (0%) |
| Southern Europe | 409 (11.5%) | 187 (13.5%) | 128 (10.5%) | 94 (9.9%) |
| Western Europe | 370 (10.4%) | 179 (12.9%) | 54 (4.4%) | 137 (14.5%) |
| Unknown | 193 (5.4%) | 52 (3.7%) | 0 (0%) | 141 (14.9%) |

### 2.2 Description of multi-cohort demographic data

To analyze associations between patient demographic variables and diagnostic markers as well as assess differences in performance across patient subgroups of classifiers of acute infection, we collected demographic information on 1) age, 2) biological sex, 3) race, and 4) ethnicity (Hispanic vs. Non-Hispanic). For the publicly available studies, these data were not immediately available and, when available, mainly comprised age and sex. For the vast majority of studies for which we did not have complete demographic data, we corresponded with each of the study’s authors to request the data (see Acknowledgements). A summary of the collected demographic data for each of the public and NanoString-profiled studies appears in Supplementary Tables 3 and 4, respectively. Sex codings were harmonized across studies to take the values ‘female’ or ‘male’. Some race groups had insufficient sample sizes for some of our downstream analyses, owing to low frequencies in the overall patient population. As such, patients from the following groups were given the race designation ‘Other’: a) patients of designated mixed race, b) ‘Native American or Alaskan Native’ patients, c) ‘Native Hawai’ian or Other Pacific Islander’ patients, d) patients already given the ‘Other’ race designation. To ensure consistent groupings and adequate sample sizes for downstream analyses, we re-coded race designations in the following way (following the FDA reporting taxonomy; [9]): 1) patients with ‘Asian’, ‘Indian sub-continent’ or ‘SE Asian’ designations were called ‘Asian’, 2) patients with ‘Black’, ‘African American’, or ‘Afro-caribbean’ designations were called ‘Black or African descent’, 3) patients with ‘Caucasian’ or ‘White’ designations were called ‘White’. In addition, we merged race and ethnicity into a single variable (‘RaceEthnicity’) by replacing the race designation of “Hispanic” patients with their ethnicity, when available. Of the 615 total patients assigned a RaceEthnicity of ‘Hispanic’, 293 listed race as ‘White’ (47.6%), 251 listed race as ‘Unknown’ (40.8%), 52 listed race as ‘Other’ (8.4%), 13 listed race as ‘Black or African descent’ (2.1%) and 6 listed race as ‘Asian’ (1%). This process resulted in six different RaceEthnicity subgroups: 1) *Asian*, 2) *Black or African descent*, 3) *Hispanic*, 4) *Other*, 5) *White* and 6) *Unknown* (in cases where RaceEthnicity information was unavailable). Besides race/ethnicity and sex, we created an additional categorical demographic variable (‘AgeBracket’) based on patient age: 1) ‘Child’ - *<* 12 years old, 2) ‘Young Adult’ - *≥* 12 and *<* 40 years old, 3) ‘Middle-Aged’ - *≥* 40 and *<* 65 years old, 4) ‘Senior’ - *≥* 65 years old.

### 2.3 Creation of patient subgroup analysis datasets

We generally limited our analyses to those patients with available demographic information. Specifically, we retained patient samples with age information and known biological sex. A total of 2622 (83%) of the initial 3159 training samples and all of the 931 validation samples met these criteria. For our Bayesian multi-level analysis of association between demographic variables and marker expression, we fitted our model to the combination of training and validation samples with age and sex information (N=3553). For our patient subgroup analysis of the pre-trained IMX-BVN-3 classifier, we removed the ‘Other’ and ‘Unknown’ RaceEthnicity groups as they lacked patients with viral infection or bacterial infection, respectively. We then evaluated the IMX-BVN-3 classifier on this subset (N=846) of the 931 validation samples. The IMX-BVN-3 classifier had been previously trained with the 3159 training samples. In our comparison of patient subgroup performance between IMX-BVN-3 and PCT, we only evaluated diagnostic classifiers in the subset of the 846 validation patients for which we had measured PCT (N=752).

### 2.4 Description of the IMX-BVN-3 diagnostic classifier

To assess performance of diagnostic classifiers of acute infection across different patient subgroups, we used the pre-trained Inflammatix BVN-3 classifier (IMX-BVN-3). Here, the abbreviation BVN represents the three infection statuses: B - bacterial, V - viral, N - non-infected.

The 29 mRNA markers used as input for the IMX-BVN-3 classifier were, in part, derived from previous work (described in [24]) but also selected *de novo* based on whether markers could be readily profiled by the loop-mediated isothermal amplification-based assay platform we’ve targeted for our diagnostic tests [16]. Tthe set of 29 markers consisted of IFI27, ZDHHC19, TGFBI, CTSB, GADD45A, CTSL1, DEFA4, PDE4B, FURIN, GNA15, OASL, C9orf95, CEACAM1, OLFM4, HLA-DMB, RAPGEF1, PER1, ARG1, KCNJ2, BATF, ISG15, KIAA1370, JUP, PSMB9, C3AR1, S100A12, LY86, and HK3.

The IMX-BVN-3 classifier is an ensemble of 10 multi-layer-perceptron neural networks. The hyperparameters for each ensemble member were tuned using the Hyperband hyperparameter search algorithm [22]. The three predicted probabilities (bacterial, viral and non-infected) computed by the ensemble members were averaged to generate the final model outputs. We performed model selection by evaluating each hyperparameter configuration used to train the classifier by multi-class AUROC (mAUC; [14]) in five-fold grouped cross-validation [24] with the publicly available microarray training data. The highest-performing hyperparameter configuration was used to train the final classifier, and this trained classifier was then applied to the independent NanoString data for validation. The hyperparameters being tuned included the number of layers; the number of nodes in each hidden layer; the optimization learning rate; the activation type; whether dropout was enabled and with what rate; whether batch-normalization was enabled; the number of training iterations; the mini-batch size; and weight regularization parameters. We also applied to the NanoString validation data the standardization transformation derived from and applied to the IMX-BVN-3 training data.

### 2.5 Bayesian multi-level regression modeling of diagnostic marker expression

The 29 markers of the IMX-BVN-3 classifier were selected for their high degree of association with patient infection status. To determine whether the markers were also significantly associated with biological (e.g. demographic attributes) or technical (e.g. assay platform) effects and whether accounting for these variables affected marker association with infection status, we specified and fit a series of 20 regression models (see Supplementary Material). Owing to computational limitations, we fit regressions to each marker individually. Prior to model fitting, we standardized each marker’s expression values to have zero mean and unit standard deviation. We also transformed patient age (square-root transformation followed by standardization).

We considered two types of regression models. For the first type of model (complete pooling - CP), we only included population-level (fixed) effects for different combinations of all of (or subsets/interactions of) 1) age, 2) sex, 3) race/ethnicity, 4) assay platform, and 5) infection status. For the second type of model (partial pooling - PP), we also included group-level (random) effects of sex and age with study used as the grouping variable. We limited our investigation of study-level effects to sex and age as some studies consisted of patients of only one race/ethnicity and were assayed on a single technical platform. To ensure identifiability of the effects, we used the following reference patient categories for each categorical covariate type: infection status - bacterial infection, sex - female, race/ethnicity - White, platform - GPL10558.

We performed Bayesian inference, approximating and summarizing posterior distributions of all model parameters by sampling using the No-U-Turn approach [17]. We further evaluated the relative ability of each of the 20 regression models to predict expression for all 29 markers by computing model stacking weights [41] with the Pareto-smoothed importance sampling approximation of leave-one-out cross-validation (LOO-PSIS; [37]). Additional details of the 20 regression models, Bayesian inference, and implementation appear in Supplementary Material.

### 2.6 Metrics of diagnostic classifier performance in validation

We evaluated the IMX-BVN-3 diagnostic classifier as well as the standard-of-care biomarker, PCT, on all patients in the validation set as well as on demographic subgroups of patients. We assessed the performance of classifiers in two ways: *within* group and *across* group. For *within* group analyses using IMX-BVN-3, we computed mAUC. For the three-class BVN classification task, mAUC is defined as the average of the AUROCs for each of the six pairwise comparisons using each of a classifier’s three predicted probabilities (e.g. bacterial- vs. viral-infected patients using bacterial predicted probabilities). For *within* group analyses of different patient demographic subgroups, all pairwise class comparisons used to compute a subgroup’s mAUC were based on the corresponding predicted probabilities of patients from that subgroup. For example, the ‘Black or African descent’ mAUC is based on pairwise class comparisons solely from ‘Black or African descent’ patients.

To develop our *across* group analyses, we used the xAUC metric [19]. Briefly, 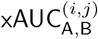 generalizes the AUC to identify pairwise group disparities in classifier performance by comparing score distributions of class *i* and *j* from patients in demographic subgroups *A* and *B*, respectively. For example, one would compute 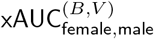 by determining whether randomly selected bacterial scores from female bacterial patients were higher on average than those from male viral patients. In our analyses, we compute both 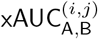 from pairwise comparisons as well as a multi-class generalization which we call xmAUC_A,B_. For the latter, we take the average of all 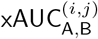 based on the six pairwise class comparisons, analogous to within-group mAUC:

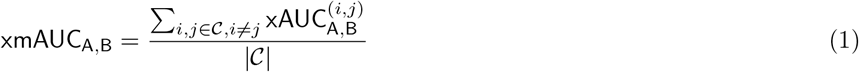

To assess pairwise across-group performance disparities, we compute ΔxAUC^(*i,j*)^_A,B_ [19]:

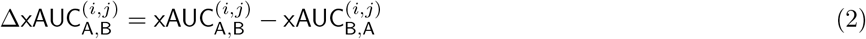

We also compute the multi-class generalization, ΔxmAUC_A,B_ as follows:

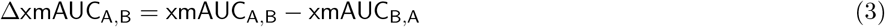

Values of these measures closer to 0 would indicate comparable performance for a given classifier across patients of subgroups A and B while deviations from 0 would indicate the presence of performance disparity for the given patient subgroups.

For the comparison of IMX-BVN-3 and PCT, we also performed both *within* and *across* group analyses, using the binary AUROC for separation of patients with bacterial infections from patients with non-bacterial inflammation (i.e. viral infections + non-infectious inflammation). We call this metric BActerial-vs-Other (‘BAO’) AUROC.

For all performance metrics, we computed 95% credible intervals using the Bayesian bootstrap [33, 11]. In brief, we follow the algorithm of Gu et al. [11] to generate posterior samples of the ROC curve for each pairwise class comparison (either within or across group). We generate 5000 posterior samples of each ROC curve. We then compute the desired AUROC-based performance metric (including averages of AUROCs) from each ROC curve sample in order to create posterior samples of each performance measure. We derive 95% credible intervals from these posterior samples by computing and reporting the 2.5th and 97.5th quantiles of all samples.

## 3 Results

### 3.1 Data survey of gene expression studies of acute infection reveals variability and gaps in patient representation

We compiled gene expression data from studies of acute infection and sepsis made publicly available on the NCBI GEO and EBI ArrayExpress databases. Details of this systematic search have been published previously [24]. This process resulted in an initial dataset of 2958 adult and pediatric patient samples across 44 studies (Supplementary Table 1). To validate diagnostic classifiers trained on these data, we profiled an additional 931 adult patients across 15 studies using the NanoString platform (Supplementary Table 2). Each patient sample was adjudicated for presence of a bacterial infection, a viral infection or non-infectious inflammation. Nearly two-thirds of the publicly available studies (N=28, 64%) corresponding to 2284 patients (77%) were profiled using Illumina assays (GPL6102, GPL6947, GPL10558) with GPL10558 being the most prominent platform. The remainder of the publicly available studies were profiled using Affymetrix (GPL570, GPL6244, GPL13158, GPL13667; N=12 studies, 27%; 470 patients, 16%) and, to a much lesser extent, Agilent (GPL10332, GPL20844; N=4 studies, 9%; 204 patients, 7%) assay platforms. For 2622 of the initial 2958 patient samples (89%; across 34 out of the 44 public studies), we were able to collect age and sex (with possibly unknown race/ethnicity; Supplementary Table 3). We compiled the same demographic information for the studies profiled on NanoString (Supplementary Table 4).

A summary of the collected data for which at least age and sex information was available (N=3553 patients) appears in Table 1. The majority of patient samples were profiled in the UK, continental Europe and the USA (N=2975, 84%), with relatively fewer patients profiled in Southern and Southeastern Asia (N=349, 10%) and Australia & New Zealand (N=36, 1%). The 193 patients of unknown geographical origin were either profiled in the UK or South Africa; the listed corresponding manuscript for the relevant study (GSE22098) did not actually contain the 193 samples, making resolution of their origin difficult. No training or validation studies took place in Africa, Central & South America, the Middle East, or Central & East Asia. Of the 3553 patients with available sex and age information, approximately 45% of patients (N=1604) were female, with roughly 13% (N=449), 9% (N=316), 12% (N=423), 5% (N=165), and 49% (N=1724) self-reporting as Asian, Black or African descent, Hispanic, Other, or White, respectively.

In the publicly available data used for classifier training, the proportion of patients of each infection type was fairly similar with roughly 34% (N=898) having a bacterial infection, 37% (N=983) having a viral infection, and 28% (N=741) having non-infectious inflammation (Supplementary Figure 3). We note that bacterial infections were relatively under-represented in the ‘Young Adult’ subgroup samples and relatively over-represented among ‘Senior’ patient samples of our training data. Viral infections were over-represented among patients with the ‘Asian’ RaceEthnicity designation, likely owing to the inclusion of multiple studies focused on viral infection from SE Asia (Table 1 and Supplementary Figure 1). We also found that the NanoString datasets were more skewed in their composition towards bacterial infections (53%; N=490) with 25% (N=235) and 22% (N=206) of patients having viral infection and non-infectious inflammation, respectively.

We found that some studies did not record race/ethnicity due to national regulations prohibiting collection of such data when it was not directly used in the outcome of the study (e.g. in France or Canada) or due to organization-level ethical considerations. Consequently, race/ethnicity information shows the highest levels of missingness in our database, particularly from Australia and continental Europe (Supplementary Table 3). Of the 3553 publicly available and NanoString-profiled patient samples with age and sex information, 476 (13%) samples did not have recorded race/ethnicity. Conversely, reporting and availability of race/ethnicity information was considerably higher for studies conducted in Southeast Asia, the UK and USA, potentially due, at least in part, to encouragement from regulatory bodies to do so [9].

### 3.2 Bayesian multi-level regression modeling identifies associations between demographic characteristics and expression of acute infection biomarkers

The marker expression used to train our diagnostic classifiers could carry associations with any of the patient demographic attributes potentially leading to bias in the training and performance of those classifiers. To assess the presence of these demographic associations, we developed a series of 20 multi-level regression models that: 1) incorporate fixed effects of different combinations of demographic covariates, including interactions between infection status and age, sex or race/ethnicity (models denoted ‘CP’) and 2) include random effects by pooling estimates of the effects of age and sex on marker expression across studies (models denoted ‘PP’). We also account for the effects of assay platform on marker expression in our regression models. We characterize uncertainty in our estimates of fixed and random effects by conducting Bayesian inference, using samples from the posterior distributions of effect coefficients to determine whether effects are significantly different from 0 (posterior samples overlap 0 barely or not at all).

Some regression models, based on different combinations of covariates (interactions and random and/or fixed effects) may better predict marker expression than others. We use a Bayesian approximation to leave-one-out cross-validation performance (LOO-PSIS; [37]) to assign weights to the different regression models for each marker; higher weight values indicate higher performance in predicting marker expression for a left-out sample using that model [41]. Overwhelmingly, we find that models that include random effects for age and sex receive the highest weights across all 29 markers (Suppl. Figure 4). These findings suggest that the associations between age/sex and marker expression are similar (though heterogeneous) across studies and that variation in marker expression isn’t as well explained by simpler models that only include fixed effects for demographic variables or assay platform. We also find that the models with highest weight for most markers are those random effects models that also include interaction terms with infection status, indicating that patient demographic status may modify the association between infection status on marker expression (Suppl. Figure 4).

To further investigate these associations, we visualized samples of the posterior distributions of population-level regression coefficients from the largest model (‘PP, All + Age:Inf + Sex:Inf. + RaceEthn:Inf.’; see Suppl. Material). In Figure 1, we show examples of these model inferences for three of the 29 selected markers: IFI27, S100A12, and CEACAM1. We could consider a marker unbiased, at least empirically, if that marker showed significant effects for ‘Noninfected’ and ‘Viral’ status (credible intervals for those terms that do not include 0), relative to the reference status, ‘Bacterial’ encoded in the intercept term, as well as no significant associations for the other covariates. Not surprisingly, we detect significant associations with infection status for nearly every marker (Figure 1, Suppl. Figures 5-33). However, we also detect significant associations with age as well as with race/ethnicity and their interaction terms for multiple markers (Supplementary Table 14), suggesting diferences in host response across patient subgroups. In Figure 1C, for example, we see significant interaction effects for CEACAM1 expression in Hispanic viral and noninfected patients. Given the sign of the interaction effects and assuming we hold other covariates fixed, these interaction effects indicate that CEACAM1 expression would more poorly separate Hispanic noninfected and viral-infected patients from Hispanic bacterial-infected patients than, say, for other patient subgroups. Indeed, we observed several markers for which there was no significant main effect for a subgroup but there were significant interaction effects (e.g. LY86 - Suppl. Figure 24, RAPGEF1 - Suppl. Figure 30), again indicating differences in host response across patient subgroups. Our analysis also identified patterns of association with age as well as race/ethnicity similar to Simpson’s paradox (Supplementary Table 16). For example, as shown in Figures 1B, age is overall positively associated with S100A12 expression but negatively associated if infection status is taken into account. We see other patterns of association with age with multiple markers (e.g. Figure 1C though not in Figure 1A), and these findings are consistent with a recent study that noted differences by age group as well as by sex in gene expression of host responses to COVID-19 [23]. We generally observe milder associations between sex (or its interaction terms with infection status) and marker expression.

In addition, though our data have undergone a co-normalization procedure to mitigate gene expression differences due to assay platform (see Supplementary Material), we still detect significant platform effects for several markers. Of the platforms represented in our multi-cohort dataset, GPL570, an Affymetrix platform and one of the oldest in use across the studies, as well as NanoString tended to show significant associations with expression for multiple (N=13) markers. Taken together, our Bayesian multi-level modeling analyses have highlighted potential sources of bias in our data, heterogeneity in host response that might necessitate additional marker selection as well as the need for further improvements to our co-normalization procedures.

**Figure 1:**
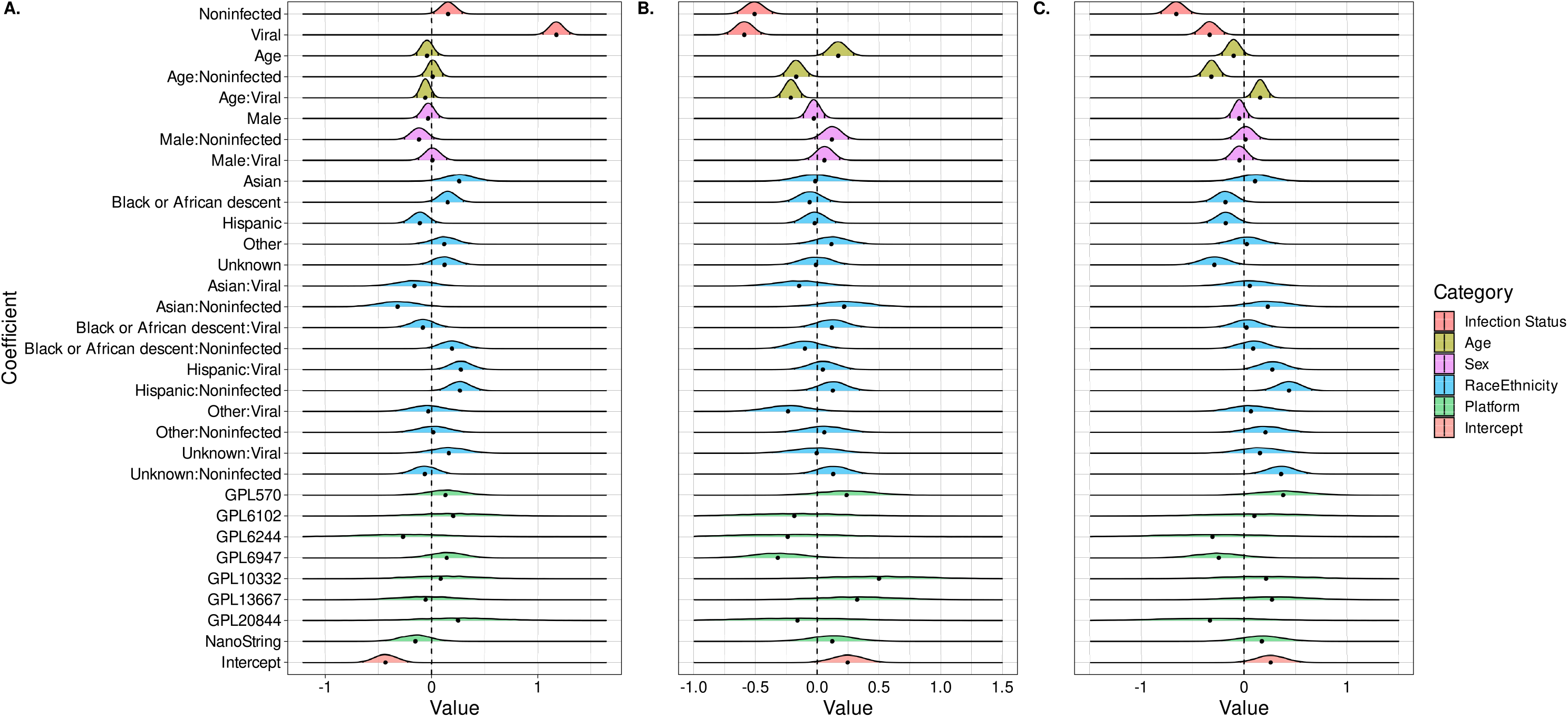
Bayesian multi-level modeling of associations between patient characteristics and markers of acute infection. Density plots of the posterior distributions of regression coefficients from the ‘PP, All + Sex:Inf. + Age:Inf. + RaceEthn:Inf.’ model for example markers, IFI27 (**A**.), S100A12 (**B**.), and CEACAM1 (**C**.). The small black lines within the density plots indicate the 2.5th and 97.5th quantiles of the posterior and the black dot corresponds to the posterior median.

### 3.3 Performance of diagnostic classifiers of acute infection varies across patient subgroups

Our Bayesian multi-level modeling analysis detected significant associations between patient demographic attributes and the markers previously selected for development of the IMX-BVN-3 diagnostic classifier. In the same spirit as previous ‘off-the-shelf’ analyses for bias in performance of pre-trained ML classifiers [3], we evaluated our IMX-BVN-3 classifier across different patient subgroups in a subset of our NanoString-profiled patient samples (N=846). We first computed multi-class AUROC (mAUC) performance measures for the full set of NanoString-profiled patients (‘Overall’) as well as *within* each patient subgroup. As the name suggests, within-group performance is based on comparisons of classifier score distributions taken from a single patient group (e.g. viral scores for young adult viral infections vs. young adult non-infectious inflammation). We show the results of this analysis in Figure 2A (also in Suppl. Table 5). We note that inference results in this and following sections will appear as ‘posterior median of performance metric, (95% credible interval)’. We find that within-group mAUC is generally comparable across the different patient subgroups, with some notable exceptions. In particular, we find that IMX-BVN-3 performs significantly worse for young adult (≥ 12yo, *<* 40yo) patients compared with middle-aged (≥ 40yo, *<* 65yo) patients young adult mAUC: 0.807, (0.778, 0.834); middle-aged mAUC: 0.868, (0.846, 0.887); Suppl.Table 5). We also find that IMX-BVN-3 performance for Asian patients in the NanoString validation set is significantly higher than for other race/ethnicity subgroups (mAUC: 0.967, (0.931, 0.988); Suppl. Table 5). However, we attribute this difference in performance to the fact that the overwhelming majority of samples (N=27) in this relatively smaller subgroup (N=39) are viral infections which have proven to be easier to distinguish from the other infection classes.

**Figure 2:**
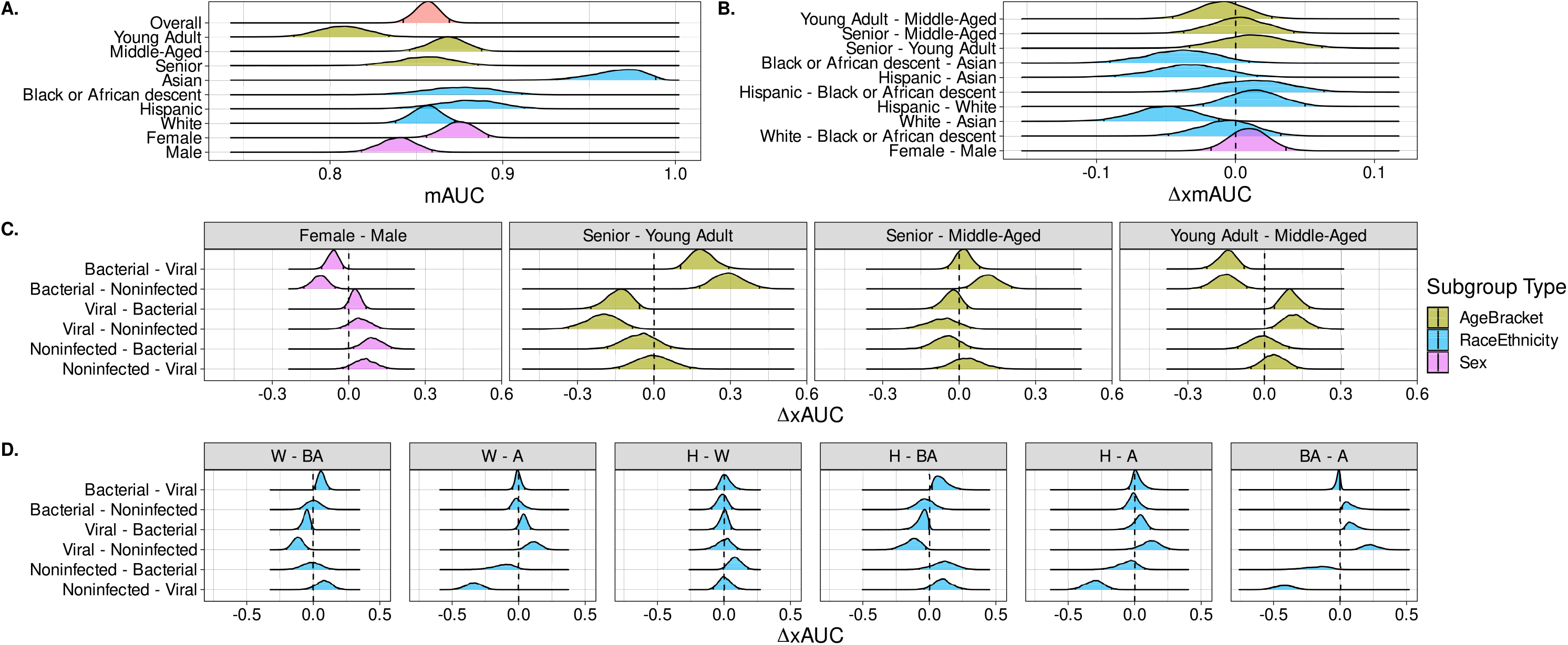
Performance of the IMX-BVN-3 diagnostic classifier within and across patient demographic subgroups. Shown are density plots of Bayesian bootstrap posterior distributions of the relevant measures of performance. Small black lines within the density indicate the 2.5th and 97.5th quantiles of the posterior sample. The density plots depict within-group measures of multi-class classification performance (mAUC; **A**.), composite measures of across-group performance (**B**.) and individual pairwise across-group performance measures (e.g. bacterial vs. viral AUROC) (**C**. and **D**.). For the across-group measures of performance, shifts of the posterior distribution away from 0 (dashed line) indicate performance disparities of the classifier for that comparison. A - Asian, BA - Black or African descent, H - Hispanic, W - White.

Recent studies have shown that performance differences in within-group AUC may not indicate disparities in classifier performance across groups [19]. So, we further analyzed IMX-BVN-3 using a multi-class generalization of the ΔxAUC metric (which we call ΔxmAUC; see Methods) to assess differences in classifier performance *across* patient subgroups. Deviations of ΔxmAUC away from 0 would indicate disparity in IMX-BVN-3 performance for the indicated subgroup comparison. We find, for the most part, no such disparities by this measure (Figure 2B., Suppl. Table 6; nearly all 95% CIs include 0). We again attribute differences in performance between Asian and White patients to the makeup of the relatively smaller Asian patient subgroup.

An evaluation of ΔxmAUC alone might mask disparities at the level of the individual pairwise comparisons (i.e. ΔxAUC) upon which ΔxmAUC is based. In other words, performance disparity may exist for, say, separation of female bacterial patients from viral patients using a classifier’s bacterial scores even though the composite measure may indicate no disparity in performance across female and male patients. When we evaluated IMX-BVN-3 in terms of ΔxAUC for each component pairwise comparison in xmAUC, we detected differences in performance between patient subgroups (Figure 2C and D, Suppl. Table 9). In particular, we find that IMX-BVN-3 bacterial scores produce poorer separation of female bacterial patients from male viral and non-infected patients (ΔxAUC^(*B,V*)^: -0.061, (−0.105, -0.019); ΔxAUC^(*B,N*)^: -0.112, (−0.174, -0.051); Suppl. Table 9). Conversely, IMX-BVN-3 bacterial scores produce relatively better separation of male bacterial patients from female viral and non-infected patients. We see stronger trends with age, as young adult patients of any infection status appear more difficult to distinguish from middle-aged and senior patients. Specifically, we find IMX-BVN-3 bacterial scores lead to relatively worse separation of young adult patients from middle-aged and senior viral and non-infected patients 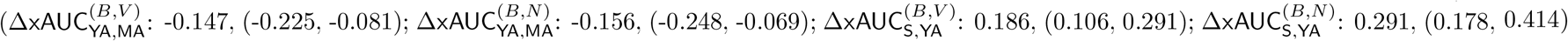; Suppl. Table 9). We also find significant performance disparities using IMX-BVN-3 viral scores: relatively better separation of young adult viral patients from middle-aged and senior bacterial and non-infected patients 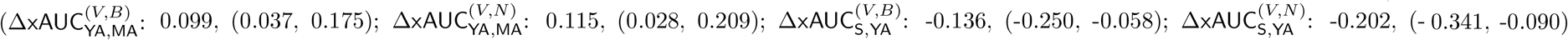; Suppl. Table 9). In addition, we observed significant differences in performance across patients of different race/ethnicity subgroups (Suppl. Table 9) such as: 1) better bacterial-viral separation using bacterial scores for White versus Black or African descent patients 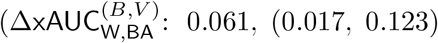, 2) worse viral-non-infected separation using viral scores for White versus Black or African descent patients 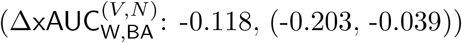, 3) worse separation of viral infection from other infection types for Hispanic versus Black or African descent patients and 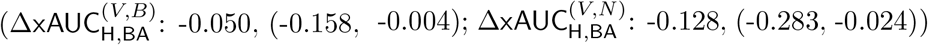 4) better bacterial-viral separation using bacterial scores for Hispanic vs. Black or African descent patients 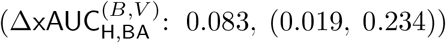. Finally, we find that despite general increases in overall and within-group performance over the three generations of the IMX-BVN classifier series, the performance disparities we observe with IMX-BVN-3 have persisted (Suppl. Figure 34, Suppl. Tables 7 & 8). While we show results for IMX-BVN-3’s non-infected scores, we note that only the bacterial and viral scores will be used as part of our planned diagnostic test report.

We extended our analysis to a biomarker, PCT, to determine whether existing clinical tools show similar within- or across-group performance disparities. Using a subset of the NanoString-profiled patients for which PCT was also collected (N=752), we computed PCT’s (and IMX-BVN-3’s) ability to separate bacterial infection from the combination of viral infections and non-infectious inflammation using the relevant within- or across-group variant of AUROC (Suppl. Tables 10 & 11). We detect significant differences between IMX-BVN-3 and PCT in overall within-group performance (IMX-BVN-3 overall mAUC: 0.887, (0.863, 0.909); PCT overall mAUC: 0.834, (0.805, 0.862); Figure 3A). We also find that, similar to IMX-BVN-3, PCT shows significant across-group performance disparities (Figure 3B, Suppl. Table 11). Specifically, PCT resulted in poorer separation of female and young adult bacterial-infected patients from male and middle-aged/senior non-bacterial-infected patients, respectively (ΔxAUC_F,M_: -0.095, (−0.154, -0.036); ΔxAUC_YA,MA_: -0.221, (−0.315, -0.134); ΔxAUC_S,YA_: 0.272, (0.167, 0.384)). Comparisons between IMX-BVN-3 and PCT across subgroups were nonsignificant, suggesting any biases in IMX-BVN-3 with respect to race may also be present in PCT (Figure 3B). A separate analysis based on an FDA-cleared gene-expression signature of infected-vs-non-infected patient status revealed similar disparities (SeptiCyte Lab^*T M*^ ; Suppl. Figures 35 & 36, Suppl. Tables 12 & 13), indicating that the potential for differences in patient subgroup performance is not limited to any one host-response-based diagnostic of acute infection. In general, these results underline the importance of inspecting performance differences across patient subgroups for any new tool both alone and in comparison to existing tools.

**Figure 3:**
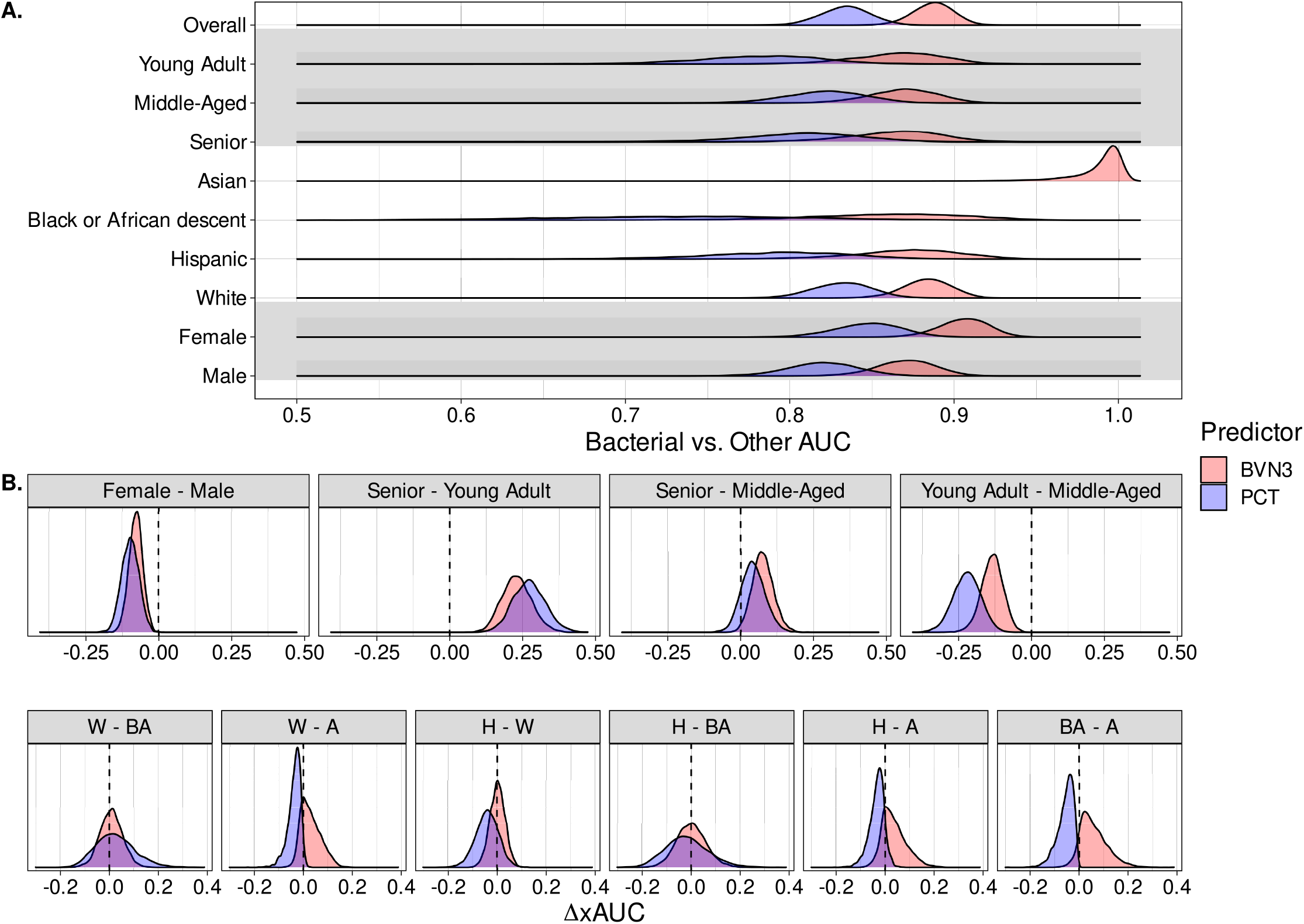
Demographic subgroup performance of IMX-BVN-3 diagnostic classifier compared with PCT. Shown are density plots of Bayesian bootstrap posterior distributions of within- (**A**.) and across- (**B**.) group measures of bacterial vs. other (viral + non-infected) classification performance. Within-group PCT performance in Asian patients does not appear as every posterior sample was 1.0, preventing rendering of the density plot. Likewise, appearance of posterior density greater than 1.0 for IMX-BVN-3 in Asian patients is an artifact of the density estimation; nearly every posterior sample was 1.0. A - Asian, BA - Black or African descent, H - Hispanic, W - White.

## 4 Discussion & Conclusion

We have developed an end-to-end framework for auditing of gene expression-based diagnostic classifiers of acute infection, drawing on recent advancements in machine learning. We uncovered variability in patient representation in our multi-cohort dataset and demographic associations with a targeted panel of host-response biomarkers as well as assessed pre-trained diagnostic classifier performance both within and across patient subgroups. As observed previously, we found that classifier performance disparities across patient subgroups could be missed by solely considering within-group performance. In addition, we found that these disparities could be missed when focusing on summary measures of performance and ignoring performance between each pair of classes in the multi-class classification setting. Moreover, despite significantly better overall performance with IMX-BVN-3, we detected performance disparities across patient subgroups for both IMX-BVN-3 and standard-of-care biomarkers for acute infection like PCT. As part of our efforts, we created an extensive data resource of demographic attributes for nearly 2700 patients with acute infection and non-infectious inflammation, profiled across a range of geographical regions, healthcare settings and assay platforms. We freely provide this resource to the community to aid further development of diagnostic classifiers and to ensure their consistent performance across diverse patient subgroups.

In the course of creating this resource, we found (as some have previously observed [5]) that some countries, organizations, and even individual investigators did not collect certain demographic information (e.g. race/ethnicity) due to ethical concerns. However, we contend as others have that, without such information, it would be impossible to assess effects of such demographic attributes on biomarker expression or to evaluate disparities in diagnostic classifier performance across patient subgroups. Indeed, our and others’ analyses indicate that patient demographic attributes are necessary for efforts to characterize and mitigate effects of these variables on biomarker expression and, consequently, on diagnostic classifier performance. We don’t claim to know the way forward for investigators in countries or organizations that forbid collection of these data, but offer this study as another piece of evidence that the harms of avoiding their collection might outweigh the benefits of preventing any potential misuse.

In our multi-cohort data survey and analysis, we revealed potential sources of bias in host-reponse-based diagnostics of acute infection. However, we stopped short of proposing remedial solutions. We did so for two reasons. First, a thorough investigation of how to mitigate the bias we observed would comprise a study in its own right and would thus be beyond the scope of this initial work. Second, and relatedly, the way to approach bias mitigation in our setting is not entirely clear. Recent advances in ML have proposed at least two sets of approach (group fairness and individual fairness), each with their own drawbacks and advantages. Group fairness approaches enforce similar model performance in expectation across subgroups by introducing ‘fairness’ penalties into classifier training [15]. However, satisfying group fairness constraints comes at the cost of overall performance, potentially leading to greater harm for patients [25, 28]. Moreover, different definitions of group fairness can be at odds with one another, making it impossible to simultaneously satisfy all of them [6, 30]. Individual fairness is based on the idea that the score produced by a classifier for an individual patient of a given class should not differ if that patient had belonged to the same class but a different subgroup. Broadly speaking, these frameworks draw on techniques from causal inference [27, 31] to account for the causative relationships between sensitive attributes (e.g. demographics), features and outcomes and to enable evaluation of the effect of changes in a sensitive attribute on performance of an ML system [21, 20, 29]. In general, we are encouraged by developments in causal inference and representation learning [34] that can allow for modeling and accounting for sources of bias in host-response marker expression. Investigation of these approaches is the subject of ongoing and future work.

There were several limitations of our study. To ensure adequate subgroup sample sizes for our analyses, we had to group together patients of potentially distinct geographic and ethnic origins (e.g. Black or African Americans with Black British patients of Caribbean descent) or remove some groups entirely (e.g. ‘Native Hawaiian or Other Pacific Islander’). We acknowledge that our results might be different with sufficiently larger sample sizes and analysis of more fine-grained patient subgroups. We believe this study represents our best effort given the available data. In a related manner, analysis of specific patient subgroups in our NanoString validation set was infeasible due to a lack of sufficient sample sizes and infection status representation. In particular, we note the lack of cases of bacterial infection and non-infectious inflammation among our Asian patient subgroup. This study helped to expose these shortages, and we are actively addressing them with targeted establishment of new clinical study sites. Another limitation of our study was in our multi-level modeling where we fit our series of regression models to each marker separately. Associations could still exist between markers even after accounting for technical platform and demographic attributes.

These data and techniques now comprise a general framework to audit pre-deployment characteristics of marker sets and candidate classifiers for all our diagnostic tests, whatever their stage of development. Implementing this framework has made us more cognizant of the need to record and account for demographic information in order to ensure high and consistent diagnostic classification performance across all segments of patient populations we hope to serve. The finding that disparities may already exist in currently used biomarker tools is also important, as it offers a minimum level of acceptable equality; a new tool may not be perfect, but it should at least be better than standard of care. We believe that as ML systems continue to appear in contexts of high-stakes decision-making, the societal impact of these systems must be continuously investigated and monitored.

## Data Availability

All demographic data from publicly available studies used in the present study are available upon reasonable request to the authors. These data will be made freely available to all as part of supplementary material upon peer-reviewed publication. Demographic data from one public study (GSE40165) was provided by the Oxford University Clinical Research Unit for analysis purposes only and cannot be redistributed.

## 5 Acknowledgements

We sincerely thank the following authors who responded to our requests for data and, without whom, this analysis would not have been possible:

- GSE40012 - Dr. Grant Parnell (Univ. of Sydney)
- GSE57065, GSE77791 - Dr. Marie Angelique Cazalis (Biomerieux)
- GSE82050 - Dr. Klaus Schughart (Helmholtz Centre for Infection Research), Dr. Ben Tang (Univ. of Sydney, Nepean Hospital)
- E-MTAB-3162 - Dr. Henk-Jan van den Ham (ENPICOM)
- GSE65682 - Dr. Brendon Scicluna (University of Malta)
- GSE27131 - Dr. Jan-Erik Berdal (University of Oslo)
- GSE13015, GSE69528 - Dr. Damien Chaussabel (Sidra Medicine), Dr. Darawan Rinchai (The Rockefeller University)
- E-MTAB-5273, E-MTAB-5274 - Dr. Katie Burnham (Wellcome Sanger Institute)
- GSE72810, GSE73461 - Dr. Jethro Herberg (Imperial College London)
- GSE63881 - Dr. Chisato Shimizu (Univ. of California - San Diego), Dr. Jane Burns (Univ. of California - San Diego, Rady Children’s Hospital)
- GSE66099 - Dr. Hector Wong (Cincinnati Children’s Hospital)
- GSE68310 - Dr. Luis Franco (NIH/NIAMS)
- GSE40165 - Ms. Evelyne Kestelyn & Dr. Bridget Wills (Oxford University Clinical Research Unit)
- GSE61821 - Dr. Long Hoang (AstraZeneca)
- E-MTAB-5638 - Dr. Ignacio Martin-Loeches (Trinity College Dublin)
- All authors who previously made patient demographic information publicly available.

We dedicate this work to the memory of Dr. Hector Wong, friend of Inflammatix and champion of sepsis research.

## Acronyms

AUROC: area under the receiver operating characteristic curve. 3, 4, 7–9
mAUC: multi-class AUC. 3, 4, 7–9
xAUC: across-group (binary) AUC. 4, 7, 9
xmAUC: across-group multi-class AUC. 4, 7, 9
BVN: bacterial-viral-noninfected. 3
CI: credible interval. 7
CP: complete pooling. 3, 5
LOO-PSIS: leave-one-out cross-validation approximation by Pareto-smoothed importance sampling. 3, 7
ML: machine learning. 1, 2, 7, 11
PCT: procalcitonin. 1, 3, 4, 9, 10
PP: partial pooling. 3, 5–7
ROC: receiver operating characteristic. 4

