## Supplementary Material for "Towards Equitable Patient Subgroup Performance by Gene-Expression-Based Diagnostic Classifiers of Acute Infection"

#### 1 Data pre-processing details

##### Within-study normalization & co-normalization

We downloaded expression data of 42 studies (comprising 3159 patients) from the NCBI GEO and EBI ArrayExpress databases. To identify and remove duplicate samples, we compared pairs of samples from different studies based on raw fluorescence signal.

For normalization, we then divided samples into pools based on platform, sample type (whole blood or buffy coat extraction) and age group (adult, pediatric; adult - >18yo). This procedure resulted in 14 pools. For samples profiled on either Agilent or Illumina microarray platforms, we corrected non-normalized expression data of individual samples by applying R's [10] limma::backgroundCorrect function (using the 'normexp' method) followed by log2 transformation. We then normalized pools of samples by applying R's limma::normalizeBetweenArrays function (using the 'quantile' method). Affymetrix array samples were normalized in pools using either the affy::rma or oligo::rma function in R with custom BrainArray CDF files (http://brainarray.mbni.med.umich.edu, Version 24, ENTREZG) in which each probeset corresponds to one NBCI Entrez gene.

For each Illumina and Agilent platform, and for each gene, probes designed for the gene were identified by aligning probe sequences to RefSeq transcript sequences. If two or more probes were retained for a gene, we computed the mean of their (log2-transformed) expression values. We discarded the probes for ZDHHC19 in Agilent arrays as it was deemed inefficient; instead, after the co-normalization step (described below), we imputed the expression values for ZDHHC19 from the remaining 28 genes in our panel by fitting a linear regression on all samples profiled on the Illumina platform.

931 samples and 30 healthy control samples were assayed in-house with Nanostring SPRINT. We normalized data between samples with housekeeping-gene (CDIPT, KPNA6, RREB1, YWHAB) normalization followed by log2(x)+1 transformation.

To account for technical and biological heterogeneity across the expression studies, we co-normalized all public microarray data to in-house Nanostring data, using healthy controls. In contrast to the procedure used in [8], we developed a different approach to co-normalization based on matching of the medians of the healthy control expression distributions for each marker. Specifically, for each gene, we calculated the median of expression values for 30 NanoString-profiled healthy controls as well as the median of expression values for the healthy controls in each pool. We then added the difference between the median of NanoString-profiled healthy control expression and the median of each pool's healthy control expression to each gene's expression values for all samples in the pool.

##### Annotating patient infection status

In addition to patient gene expression measurements, we collected labels for patient infection status. Samples were labeled as Bacterial, Viral, Non-infected, Unknown and Healthy controls, based on: 1) "Characteristics" data, when available in the NCBI GEO/ArrayExpress record for the study, 2) data provided in the study's original manuscript or supplementary material, or 3) sample title or description metadata taken from the study's NCBI GEO/ArrayExpress record. Data for Unknown samples were pre-processed along with the rest of the samples, but were not used for subsequent analysis. Non-infected determinations did not include healthy controls.

##### Selection of single patient-temporal replicate samples from GlueBuffyHCSS longitudinal study data

Previous datasets used to train the IMX-BVN-3 classifier included multiple samples per non-infected patient from the GlueBuffyHCSS study [12, 13]. These samples had been collected at different time points in their hospital stay. To facilitate our analyses, we selected a single time point as the representative sample for a given non-infected patient. We first identified the unique IDs for patients in the GlueBuffyHCSS study with either a bacterial infection or noninfectious inflammation. This procedure resulted in 45 unique patients with bacterial infection and 74 unique patients with noninfectious inflammation. We then performed propensity matching with the MatchIt package [6], pairing each non-infected patient with a single infected patient based on age, sex, race, and ethnicity (binary; Hispanic origin or not). As we had more non-infected than infected patients, some infected patients were paired with more than one non-infected patient. We then selected the time point from each non-infected patient that was closest in hours to the 'time since injury' of their paired infected patient. We used the expression data corresponding to this time point as the representative expression for that non-infected sample in our analyses. Removing temporal replicate samples in this way resulted in 2958 total public study samples for our downstream analyses compared with the 3159 public study samples used to train IMX-BVN-3.

### 2 Bayesian multi-level regression model details

#### Model specification

To assess the degree of association between different sample covariates (demographic and technical) and the expression of our 29 diagnostic markers, we specified and fit a series of regression models, including different combinations of fixed (main and/or interaction) and group effects. We modeled the expected marker expression for the $i$th patient sample and the $j$th marker ($\mathsf{M}_{i,j}$) as a linear combination of effects from patient covariates (Age - $\mathsf{A}_i$, Sex - $\mathsf{S}_i$, RaceEthnicity - $\mathsf{R}_i$), infection status ($\mathsf{F}_i$) and/or assay platform ($\mathsf{P}_i$). Models that only contained fixed/interaction effects were denoted 'CP' for 'complete pooling'. In the case of models with 'partial pooling' ('PP') or group effects, we used study (as given by their unique study ID) as the grouping variable. In our notation, we assume that categorical covariates are one-hot-encoded binary vectors with the reference category taking a 0 value for each item in the vector. For example, with bacterial infection being the reference category, a patient with viral infection would have a covariate vector, $\mathsf{F}_i = [[1], [0]]$. Correspondingly, the coefficient vector for infection status ($\boldsymbol{\beta}_{\mathsf{F}}$) would be two-dimensional. For all models, we assume that the noise term, $\epsilon$ is independent across samples, independent across markers, and Gaussian-distributed with zero-mean. We also estimate a marker-specific intercept term, $\alpha_j$ that represents the given marker's mean expression for patients with a set of reference covariates values patient covariates. We used this reference formulation to enable identifiability of the effects, and we used bacterial infection, female, White, average standardized age, and GPL10558 as the references values in our analyses. Prior to model fitting, we standardized each marker's expression values to have zero mean and unit standard deviation. We also transformed patient age (square-root transformation followed by standardization). Posterior summaries of the effects from model 'PP, All + Age:Inf. + Sex:Inf. + RaceEthn:Inf.' appear in Supplementary Figures 5-33 and Supplementary Table 14. Specific details of the terms included in each model appear below:

**'CP, Demo.' - fixed effects for patient demographics & assay platform**

$$\mathsf{M}_{i,j} = \alpha_j + \beta_{j,\mathsf{A}}\mathsf{A}_i + \boldsymbol{\beta}'_{j,\mathsf{R}}\mathsf{R}_{\boldsymbol{i}} + \boldsymbol{\beta}'_{j,\mathsf{S}}\mathsf{S}_{\boldsymbol{i}} + \boldsymbol{\beta}'_{j,\mathsf{P}}\mathsf{P}_{\boldsymbol{i}} + \boldsymbol{\beta}'_{j,\mathsf{F}}\mathsf{F}_{\boldsymbol{i}} + \epsilon$$

**'CP, Platform' - fixed effects for assay platform only**

$$\mathsf{M}_{i,j} = \alpha_j + \boldsymbol{\beta}'_{j,\mathsf{P}}\mathsf{P}_{\boldsymbol{i}} + \epsilon$$

**'CP, Inf.' - fixed effects for infection status only**

$$\mathsf{M}_{i,j} = \alpha_j + \boldsymbol{\beta}'_{j,\mathsf{F}}\mathsf{F}_{\boldsymbol{i}} + \epsilon$$

**'CP, Inf. + Platform' - fixed effects for infection status & assay platform**

$$\mathsf{M}_{i,j} = \alpha_j + \boldsymbol{\beta}'_{j,\mathsf{P}}\mathsf{P}_{\boldsymbol{i}} + \boldsymbol{\beta}'_{j,\mathsf{F}}\mathsf{F}_{\boldsymbol{i}} + \epsilon$$

**'CP, All' - fixed effects for patient demographics, infection status & assay platform**

$$\mathsf{M}_{i,j} = \alpha_j + \beta_{j,\mathsf{A}}\mathsf{A}_i + \boldsymbol{\beta}'_{j,\mathsf{R}}\mathsf{R}_{\boldsymbol{i}} + \boldsymbol{\beta}'_{j,\mathsf{S}}\mathsf{S}_{\boldsymbol{i}} + \boldsymbol{\beta}'_{j,\mathsf{P}}\mathsf{P}_{\boldsymbol{i}} + \boldsymbol{\beta}'_{j,\mathsf{F}}\mathsf{F}_{\boldsymbol{i}} + \epsilon$$

**'CP, All + Sex:Inf.' - fixed effects for demographics, infection status & assay platform; interaction terms between infection status & sex**

$$\mathsf{M}_{i,j} = \alpha_j + \beta_{j,\mathsf{A}}\mathsf{A}_i + \boldsymbol{\beta}'_{j,\mathsf{R}}\mathsf{R}_{\boldsymbol{i}} + \boldsymbol{\beta}'_{j,\mathsf{S}}\mathsf{S}_{\boldsymbol{i}} + \boldsymbol{\beta}'_{j,\mathsf{P}}\mathsf{P}_{\boldsymbol{i}} + \boldsymbol{\beta}'_{j,\mathsf{F}}\mathsf{F}_{\boldsymbol{i}} + \boldsymbol{\beta}'_{j,\mathsf{F}:\mathsf{S}}\mathsf{F}_{\boldsymbol{i}} : \mathsf{S}_{\boldsymbol{i}} + \epsilon$$

**'CP, All + Age:Inf.' - fixed effects for demographics, infection status & assay platform; interaction terms between infection status & age**

$$\mathsf{M}_{i,j} \quad = \quad \alpha_j + \beta_{j,\mathsf{A}}\mathsf{A}_i + \boldsymbol{\beta}'_{\boldsymbol{j},\mathsf{R}}\mathsf{R}_{\boldsymbol{i}} + \boldsymbol{\beta}'_{\boldsymbol{j},\mathsf{S}}\mathsf{S}_{\boldsymbol{i}} + \boldsymbol{\beta}'_{\boldsymbol{j},\mathsf{P}}\mathsf{P}_{\boldsymbol{i}} + \boldsymbol{\beta}'_{\boldsymbol{j},\mathsf{F}}\mathsf{F}_{\boldsymbol{i}} + \boldsymbol{\beta}'_{\boldsymbol{j},\mathsf{F:A}}\mathsf{F}_{\boldsymbol{i}} : \mathsf{A}_i + \epsilon$$

**'CP, All + RaceEthn:Inf.' - fixed effects for demographics, infection status & assay platform; interaction terms between infection status & race/ethnicity**

$$\mathsf{M}_{i,j} \quad = \quad \alpha_j + \beta_{j,\mathsf{A}}\mathsf{A}_i + \boldsymbol{\beta}'_{\boldsymbol{j},\mathsf{R}}\mathsf{R}_{\boldsymbol{i}} + \boldsymbol{\beta}'_{\boldsymbol{j},\mathsf{S}}\mathsf{S}_{\boldsymbol{i}} + \boldsymbol{\beta}'_{\boldsymbol{j},\mathsf{P}}\mathsf{P}_{\boldsymbol{i}} + \boldsymbol{\beta}'_{\boldsymbol{j},\mathsf{F}}\mathsf{F}_{\boldsymbol{i}} + \boldsymbol{\beta}'_{\boldsymbol{j},\mathsf{F:R}}\mathsf{F}_{\boldsymbol{i}} : \mathsf{R}_{\boldsymbol{i}} + \epsilon$$

**'CP, All + Sex:Inf. + Age:Inf.' - fixed effects for demographics, infection status & assay platform; interaction terms between infection status & both sex and age**

$$\mathsf{M}_{i,j} \quad = \quad \alpha_j + \beta_{j,\mathsf{A}}\mathsf{A}_i + \boldsymbol{\beta}'_{\boldsymbol{j},\mathsf{R}}\mathsf{R}_{\boldsymbol{i}} + \boldsymbol{\beta}'_{\boldsymbol{j},\mathsf{S}}\mathsf{S}_{\boldsymbol{i}} + \boldsymbol{\beta}'_{\boldsymbol{j},\mathsf{P}}\mathsf{P}_{\boldsymbol{i}} + \boldsymbol{\beta}'_{\boldsymbol{j},\mathsf{F}}\mathsf{F}_{\boldsymbol{i}} + \boldsymbol{\beta}'_{\boldsymbol{j},\mathsf{F:S}}\mathsf{F}_{\boldsymbol{i}} : \mathsf{S}_{\boldsymbol{i}} + \boldsymbol{\beta}'_{\boldsymbol{j},\mathsf{F:A}}\mathsf{F}_{\boldsymbol{i}} : \mathsf{A}_i + \epsilon$$

**'CP, All + Age:Inf. + RaceEthn:Inf.' - fixed effects for demographics, infection status & assay platform; interaction terms between infection status & both race/ethnicity and age**

$$\mathsf{M}_{i,j} \quad = \quad \alpha_j + \beta_{j,\mathsf{A}}\mathsf{A}_i + \boldsymbol{\beta}'_{\boldsymbol{j},\mathsf{R}}\mathsf{R}_{\boldsymbol{i}} + \boldsymbol{\beta}'_{\boldsymbol{j},\mathsf{S}}\mathsf{S}_{\boldsymbol{i}} + \boldsymbol{\beta}'_{\boldsymbol{j},\mathsf{P}}\mathsf{P}_{\boldsymbol{i}} + \boldsymbol{\beta}'_{\boldsymbol{j},\mathsf{F}}\mathsf{F}_{\boldsymbol{i}} + \boldsymbol{\beta}'_{\boldsymbol{j},\mathsf{F:R}}\mathsf{F}_{\boldsymbol{i}} : \mathsf{R}_{\boldsymbol{i}} + \boldsymbol{\beta}'_{\boldsymbol{j},\mathsf{F:A}}\mathsf{F}_{\boldsymbol{i}} : \mathsf{A}_i + \epsilon$$

**'CP, All + Sex:Inf. + RaceEthn:Inf.' - fixed effects for demographics, infection status & assay platform; interaction terms between infection status & both race/ethnicity and sex**

$$\mathsf{M}_{i,j} \quad = \quad \alpha_j + \beta_{j,\mathsf{A}}\mathsf{A}_i + \boldsymbol{\beta}'_{\boldsymbol{j},\mathsf{R}}\mathsf{R}_{\boldsymbol{i}} + \boldsymbol{\beta}'_{\boldsymbol{j},\mathsf{S}}\mathsf{S}_{\boldsymbol{i}} + \boldsymbol{\beta}'_{\boldsymbol{j},\mathsf{P}}\mathsf{P}_{\boldsymbol{i}} + \boldsymbol{\beta}'_{\boldsymbol{j},\mathsf{F}}\mathsf{F}_{\boldsymbol{i}} + \boldsymbol{\beta}'_{\boldsymbol{j},\mathsf{F:R}}\mathsf{F}_{\boldsymbol{i}} : \mathsf{R}_{\boldsymbol{i}} + \boldsymbol{\beta}'_{\boldsymbol{j},\mathsf{F:S}}\mathsf{F}_{\boldsymbol{i}} : \mathsf{S}_{\boldsymbol{i}} + \epsilon$$

**'CP, All + Age:Inf. + Sex:Inf. + RaceEthn:Inf.' - fixed effects for demographics, infection status & assay platform; interaction terms between infection status & race/ethnicity, age and sex**

$$\mathsf{M}_{i,j} \quad = \quad \alpha_j + \beta_{j,\mathsf{A}}\mathsf{A}_i + \boldsymbol{\beta}'_{\boldsymbol{j},\mathsf{R}}\mathsf{R}_{\boldsymbol{i}} + \boldsymbol{\beta}'_{\boldsymbol{j},\mathsf{S}}\mathsf{S}_{\boldsymbol{i}} + \boldsymbol{\beta}'_{\boldsymbol{j},\mathsf{P}}\mathsf{P}_{\boldsymbol{i}} + \boldsymbol{\beta}'_{\boldsymbol{j},\mathsf{F}}\mathsf{F}_{\boldsymbol{i}} + \boldsymbol{\beta}'_{\boldsymbol{j},\mathsf{F:R}}\mathsf{F}_{\boldsymbol{i}} : \mathsf{R}_{\boldsymbol{i}} + \boldsymbol{\beta}'_{\boldsymbol{j},\mathsf{F:A}}\mathsf{F}_{\boldsymbol{i}} : \mathsf{A}_{\boldsymbol{i}} + \boldsymbol{\beta}'_{\boldsymbol{j},\mathsf{F:S}}\mathsf{F}_{\boldsymbol{i}} : \mathsf{S}_{\boldsymbol{i}} + \epsilon$$

**'PP, All' - fixed effects for demographics, infection status & platform; study effects for age and sex**

$$\mathsf{M}_{i,j,k} \quad = \quad \alpha_{j,k} + \beta_{j,k,\mathsf{A}}\mathsf{A}_i + \boldsymbol{\beta}'_{\boldsymbol{j},\mathsf{R}}\mathsf{R}_i + \boldsymbol{\beta}'_{\boldsymbol{j},\boldsymbol{k},\mathsf{S}}\mathsf{S}_{\boldsymbol{i}} + \boldsymbol{\beta}'_{\boldsymbol{j},\mathsf{P}}\mathsf{P}_{\boldsymbol{i}} + \boldsymbol{\beta}'_{\boldsymbol{j},\mathsf{F}}\mathsf{F}_{\boldsymbol{i}} + \epsilon$$

**'PP, All + Sex:Inf.' - fixed effects for demographics, infection status & assay platform; interaction terms between infection status & sex; study effects for age and sex**

$$M_{i,j} \quad = \quad \alpha_{j,k} + \beta_{j,k,\mathsf{A}}\mathsf{A}_i + \boldsymbol{\beta}'_{\boldsymbol{j},\mathsf{R}}\mathsf{R}_{\boldsymbol{i}} + \boldsymbol{\beta}'_{\boldsymbol{j},\boldsymbol{k},\mathsf{S}}\mathsf{S}_{\boldsymbol{i}} + \boldsymbol{\beta}'_{\boldsymbol{j},\mathsf{P}}\mathsf{P}_{\boldsymbol{i}} + \boldsymbol{\beta}'_{\boldsymbol{j},\mathsf{F}}\mathsf{F}_{\boldsymbol{i}} + \boldsymbol{\beta}'_{\boldsymbol{j},\mathsf{F:S}}\mathsf{F}_{\boldsymbol{i}} : \mathsf{S}_{\boldsymbol{i}} + \epsilon$$

**'PP, All + Age:Inf.' - fixed effects for demographics, infection status & assay platform; interaction terms between infection status & age; study effects for age and sex**

$$M_{i,j} \quad = \quad \alpha_{j,k} + \beta_{j,k,\mathsf{A}}\mathsf{A}_i + \boldsymbol{\beta}'_{\boldsymbol{j},\mathsf{R}}\mathsf{R}_{\boldsymbol{i}} + \boldsymbol{\beta}'_{\boldsymbol{j},\boldsymbol{k},\mathsf{S}}\mathsf{S}_{\boldsymbol{i}} + \boldsymbol{\beta}'_{\boldsymbol{j},\mathsf{P}}\mathsf{P}_{\boldsymbol{i}} + \boldsymbol{\beta}'_{\boldsymbol{j},\mathsf{F}}\mathsf{F}_{\boldsymbol{i}} + \boldsymbol{\beta}'_{\boldsymbol{j},\mathsf{F:A}}\mathsf{F}_{\boldsymbol{i}} : \mathsf{A}_i + \epsilon$$

**'PP, All + RaceEthn:Inf.' - fixed effects for demographics, infection status & assay platform; interaction terms between infection status & race/ethnicity; study effects for age and sex**

$$M_{i,j} \quad = \quad \alpha_{j,k} + \beta_{j,k,\mathsf{A}}\mathsf{A}_i + \boldsymbol{\beta}'_{\boldsymbol{j},\mathsf{R}}\mathsf{R}_{\boldsymbol{i}} + \boldsymbol{\beta}'_{\boldsymbol{j},\boldsymbol{k},\mathsf{S}}\mathsf{S}_{\boldsymbol{i}} + \boldsymbol{\beta}'_{\boldsymbol{j},\mathsf{P}}\mathsf{P}_{\boldsymbol{i}} + \boldsymbol{\beta}'_{\boldsymbol{j},\mathsf{F}}\mathsf{F}_{\boldsymbol{i}} + \boldsymbol{\beta}'_{\boldsymbol{j},\mathsf{F:R}}\mathsf{F}_{\boldsymbol{i}} : \mathsf{R}_{\boldsymbol{i}} + \epsilon$$

**'PP, All + Sex:Inf. + Age:Inf.' - fixed effects for demographics, infection status & assay platform; interaction terms between infection status & age and sex; study effects for age and sex**

$$M_{i,j} \quad = \quad \alpha_{j,k} + \beta_{j,k,\mathsf{A}}\mathsf{A}_i + \boldsymbol{\beta}'_{\boldsymbol{j},\mathsf{R}}\mathsf{R}_i + \boldsymbol{\beta}'_{\boldsymbol{j},\boldsymbol{k},\mathsf{S}}\mathsf{S}_{\boldsymbol{i}} + \boldsymbol{\beta}'_{\boldsymbol{j},\mathsf{P}}\mathsf{P}_{\boldsymbol{i}} + \boldsymbol{\beta}'_{\boldsymbol{j},\mathsf{F}}\mathsf{F}_{\boldsymbol{i}} + \boldsymbol{\beta}'_{\boldsymbol{j},\mathsf{F:A}}\mathsf{F}_{\boldsymbol{i}} : \mathsf{A}_i + \boldsymbol{\beta}'_{\boldsymbol{j},\mathsf{F:S}}\mathsf{F}_{\boldsymbol{i}} : \mathsf{S}_{\boldsymbol{i}} + \epsilon$$

**"PP, All + Age:Inf. + RaceEthn:Inf.' - fixed effects for demographics, infection status & assay platform; interaction terms between infection status & race/ethnicity and age; study effects for age and sex**

$$M_{i,j} = \alpha_{j,k} + \beta_{j,k,\mathsf{A}}\mathsf{A}_i + \boldsymbol{\beta}'_{j,\mathsf{R}}\mathsf{R}_i + \boldsymbol{\beta}'_{j,k,\mathsf{S}}\mathsf{S}_i + \boldsymbol{\beta}'_{j,\mathsf{P}}\mathsf{P}_i + \boldsymbol{\beta}'_{j,\mathsf{F}}\mathsf{F}_i + \boldsymbol{\beta}'_{j,\mathsf{F:R}}\mathsf{F}_i : \mathsf{R}_i + \boldsymbol{\beta}'_{j,\mathsf{F:A}}\mathsf{F}_i : \mathsf{A}_i + \epsilon$$

**'PP, All + Sex:Inf. + RaceEthn:Inf.' - fixed effects for demographics, infection status & assay platform; interaction terms between infection status & race/ethnicity and sex; study effects for age and sex**

$$M_{i,j} = \alpha_{j,k} + \beta_{j,k,\mathsf{A}}\mathsf{A}_i + \boldsymbol{\beta}'_{j,\mathsf{R}}\mathsf{R}_i + \boldsymbol{\beta}'_{j,k,\mathsf{S}}\mathsf{S}_i + \boldsymbol{\beta}'_{j,\mathsf{P}}\mathsf{P}_i + \boldsymbol{\beta}'_{j,\mathsf{F}}\mathsf{F}_i + \boldsymbol{\beta}'_{j,\mathsf{F:R}}\mathsf{F}_i : \mathsf{R}_i + \boldsymbol{\beta}'_{j,\mathsf{F:S}}\mathsf{F}_i : \mathsf{S}_i + \epsilon$$

**'PP, All + Age:Inf. + Sex:Inf. + RaceEthn:Inf.' - fixed effects for demographics, infection status & assay platform; interaction terms between infection status & race/ethnicity, sex and age; study effects for age and sex**

$$M_{i,j} = \alpha_{j,k} + \beta_{j,k,\mathsf{A}}\mathsf{A}_i + \boldsymbol{\beta}'_{j,\mathsf{R}}\mathsf{R}_i + \boldsymbol{\beta}'_{j,k,\mathsf{S}}\mathsf{S}_i + \boldsymbol{\beta}'_{j,\mathsf{P}}\mathsf{P}_i + \boldsymbol{\beta}'_{j,\mathsf{F}}\mathsf{F}_i + \boldsymbol{\beta}'_{j,\mathsf{F:R}}\mathsf{F}_i : \mathsf{R}_i + \boldsymbol{\beta}'_{j,\mathsf{F:A}}\mathsf{F}_i : \mathsf{A}_i + \boldsymbol{\beta}'_{j,\mathsf{F:S}}\mathsf{F}_i : \mathsf{S}_i + \epsilon$$

#### Prior specification & prior predictive model checking

In carrying out Bayesian inference, we first specify prior distributions for the multi-level model parameters. These parameters consist of the population-level fixed effects for each marker $j$ (coefficients, $\beta_{j,\cdot}$), standard deviations of both the group-level effects (intercept - $\sigma_{j,\alpha}$ and coefficients - $\sigma_{j,\mathsf{Age}}$, $\sigma_{j,\mathsf{Sex}}$) as well as of the additive Gaussian noise term, $\epsilon$ ($\sigma_j$), and the population-level intercept ($\alpha_j$). These prior distributions appear below:

$$\begin{aligned} \beta_{j,\cdot} &\sim \mathsf{Student\text{-}t}(3, 0, 1.0) \\ \sigma_{j,\alpha},\ \sigma_{j,\mathsf{Age}},\ \sigma_{j,\mathsf{Sex}},\ \sigma_j &\sim \mathsf{half\text{-}Student\text{-}t}(3, 0, 1.5) \\ \alpha_j &\sim \mathsf{Student\text{-}t}(3, \hat{\mu}_j, 1.5) \end{aligned}$$

.

The Student-t distributions for the population-level fixed effects reflected our weak prior beliefs that biological and technical covariates would have little to no effect on marker expression while still allowing for non-zero (and potentially large) effects for some covariates. For the population-level intercept term, $\alpha_j$, we extracted and used the brms package default prior location for each marker ($\hat{\mu}_j$ above) while setting the degrees of freedom and scale to 3 and 1.5, respectively. Prior predictive simulations with the model named 'CP, All' indicated that the ranges of marker expression observed in our data were well-covered by these settings.

#### Multi-level model fitting and convergence assessment

Fitting of the multi-level models was carried out in a Bayesian manner. We used the NUTS sampler [7] to generate samples from each parameter's posterior distribution with an adaptive delta parameter value of 0.999 and a maximum tree depth of 15. We generated 4k samples from each of 3 parallel chains with half of the samples from each chain used for burn-in. None of the posterior samples for any model parameter across all 29×20 marker-model combinations attained a potential scale reduction factor metric ($\hat{R}$; [5]) value greater than 1.01.

#### Implementation details

All model-fitting, analysis and plotting code was implemented in the R programming language [10]. In order to fit and compare the Bayesian multi-level models, we used the brms [3, 2] package.

#### Model weighting by stacking of LOO-PSIS estimates of predictive performance

To compare the different multi-level models in terms of their ability to predict marker expression, we compute model stacking weights [15] based on Pareto-smoothed importance sampling (PSIS) estimates of each model's leave-one-sample-out cross-validation (LOOCV) performance [14]. In brief, the PSIS estimates approximate each model's LOOCV performance based on the posterior using the full likelihood. These estimates are reliable provided that the Pareto shape parameter $k$ is less than 0.5; though estimates were shown to have good performance at values of $k$ as high as 0.7 [14]. For all models, we did not observe $k$ values greater than 0.7, indicating that the PSIS-based estimates of LOOCV performance could be used. We then computed model stacking weights [15], assessing the relative predictive performance of the 20 multi-level models for the expression of each biomarker. The model stacking weights correspond to the combination of the multi-level models that maximizes leave-one-out predictive density for a given marker's expression. As such, the weight assigned to each multi-level model (summing to 1) indicates that model's relative contribution (compared with the other models) to the leave-one-out predictive density. Results of this analysis appear in Supplementary Figure 4.

### 3 Diagnostic classifier performance comparison

#### Comparison of performance across IMX-BVN-x classifier generations

Our IMX-BVN diagnostic classifier has undergone multiple rounds of development, each of which involved the use of different features and datasets for training. To assess how those changes affected within- and across-group performance, we evaluated the first (IMX-BVN-1 [8]), second (IMX-BVN-2 [1, 11] and third (current, IMX-BVN-3) generations of the classifier in our NanoString-profiled validation set (N=846; patients with 'Other' and 'Unknown' RaceEthnicity designations were removed due to a lack of bacterial infection and viral infection patients, respectively). We briefly detail how these classifiers differed: IMX-BVN-1 - a multi-layer perceptron trained on the geometric means of six modules of acute-infection-associated gene expression derived from 1069 adult patient samples across 18 studies; IMX-BVN-2 - a logistic regression classifier trained on a combination of 41 features (gene expression from 29 markers, geometric means of six expression modules used for IMX-BVN-1, arithmetic means of six expression modules used for IMX-BVN-1) derived from 3413 adult and pediatric patient samples across 43 studies; IMX-BVN-3 - an ensemble of multi-layer perceptrons trained on gene expression features from 29 markers (different from those used for IMX-BVN-1 and IMX-BVN-2) derived from 3159 adult and pediatric patient samples across 42 studies. We note that one study of NanoString-profiled bacterial and viral infections from IMX-BVN-2 development was removed for IMX-BVN-3 training.

In terms of within-group performance (Suppl. Figure 34A; Suppl. Table 5), IMX-BVN-3 is better or as good as IMX-BVN-2 across all patient categories. Composite across-group performance (Suppl. Figure 34B; Suppl. Table 6) does not indicate any substantial differences in performance between the two generations of the BVN classifier. We note that the shifted distribution of composite across-group performance for the Black or African descent - Asian and White - Asian comparisons is likely due to the over-representation of viral infections in the relatively smaller Asian patients subgroup. In terms of across-group performance for individual pairwise classification tasks (Suppl. Figures 34C and D; Suppl. Tables 7, 8, and 9), we find that IMX-BVN-3 shows generally less (or no more) disparity in performance compared with IMX-BVN-2: the IMX-BVN-3 performance densities are closer to or straddling 0. Again, we note that we disregard across-group measures of performance that include the non-infected scores as they will not be used in the diagnostic report in our final product.

#### Comparison of classifier performance between IMX-BVN-3 and SeptiCyte Lab$^{TM}$

To further evaluate within- and across-group performance, we compared the IMX-BVN-3 classifier with a 4-marker gene expression signature inspired by ImmuneExpress' SeptiScore$^{TM}$ [9]. We refer to this score derived from the 4 SeptiScore markers as 'SC4'. SC4 is a linear combination of the log-normalized expression values of the markers PLAC8, PLA2G7, LAMP1 and CEACAM4. We compute the SC4 score with the following equation:

$$SC4 = PLAC8 - PLA2G7 + LAMP1 - CEACAM4 \tag{1}$$

. This 4-marker signature was developed for discrimination of sepsis patients from infection-negative patients with systemic inflammation [9]. As such, we relabel all bacterial and viral infected patients in our validation cohort as '1' and all non-infected patients as '0'. Furthermore, to facilitate performance comparison of SC4 and IMX-BVN-3, we take the sum of IMX-BVN-3's predicted probability scores for the bacterial and viral infection classes as a single 'infected' score. We then evaluate either the SC4 score or the IMX-BVN-3 'infected' score with the corresponding within- or across-group performance metric (Suppl. Figure 35, Supplementary Tables 12 and 13). We were able to compute both SC4 and IMX-BVN-3 'infected' scores for 689 patients from our NanoString validation set.

In terms of within-group performance (Supplementary Table 12), IMX-BVN-3 is significantly higher for female patients (IMX-BVN-3 AUC - posterior median: 0.805, 95% posterior CI: (0.752, 0.850); SC4 AUC - posterior median: 0.691, 95% posterior CI: (0.632, 0.745)). Both IMX-BVN-3-based and SC4-based scores show nearly indistinguishable within-group performance in male patients, young patients and Black or African descent or Hispanic patients. In terms of across-group performance (Supplementary Table 13), IMX-BVN-3 demonstrates significantly less performance disparity than SC4 between White and Black or African descent patients (IMX-BVN-3 $\Delta\mathsf{xAUC}_{S,YA}$ - posterior median: -0.065, 95% posterior CI: (-0.194, 0.077); SC4 $\Delta\mathsf{xAUC}_{S,YA}$ - posterior median: -0.386, 95% posterior CI: (-0.492, -0.271)). Specifically, SC4 scores result in relatively poorer separation of White infected patients from Black or African descent non-infected patients. This finding is consistent with previous analyses of SC4 performance that showed higher infected scores for Black or African descent patients with systemic inflammatory response syndrome but no confirmed infection [4]. Conversely, SC4 shows significantly less performance disparity than IMX-BVN-3 between male and female patients (IMX-BVN-3 $\Delta\mathsf{xAUC}_{F,M}$ - posterior median: -0.085, 95% posterior CI: (-0.165, -0.012); SC4 $\Delta\mathsf{xAUC}_{F,M}$ - posterior median: 0.003, 95% posterior CI: (-0.080, 0.081)). We caution over-interpretation of the observed disparities between either White or Hispanic patients and Asian patients due to the limited number of Asian patients in this analysis. Consistent with our findings with PCT, this analysis shows the broader potential for bias in diagnostic classifiers of acute infection.

In the previous analysis we modified the IMX-BVN-3 predicted probability scores to match the intended use of the SC4 panel (infected-vs-non-infected classification). We also evaluate the within-group performance of the IMX-BVN-3 bacterial predicted probabilities and the SC4 scores to separate bacterial infection from non-bacterial inflammation, a task aligned

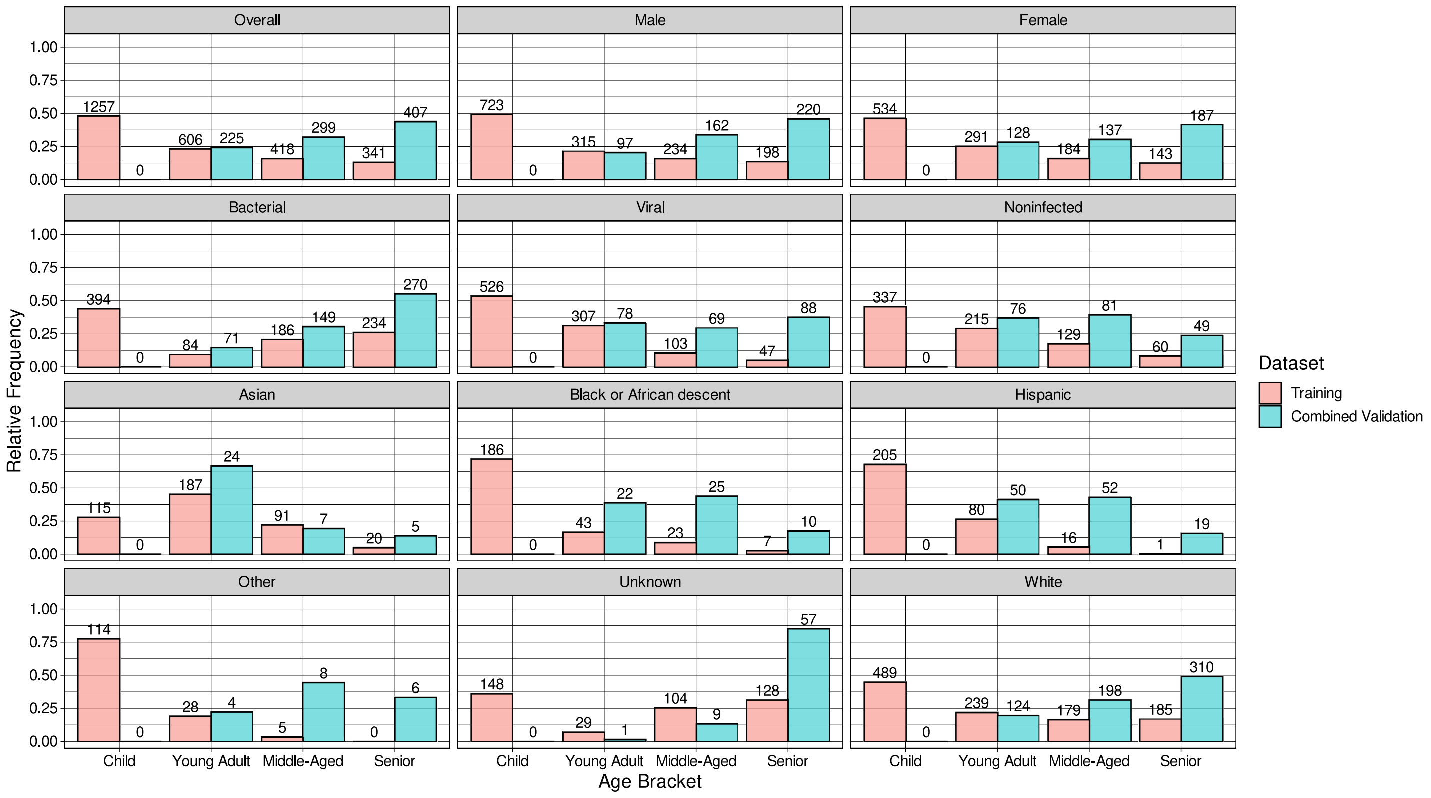


Supplementary Figure 1: **Breakdown of demographic composition of datasets of acute infection patients by age**. Hispanic and Black or African descent patient subgroups both showed skews in their age distribution in our training datasets with children and young adults being relatively more represented. We also observe relative under-representation of young adults among our training samples with bacterial infections and of older (middle-aged or senior) patients among our training samples with viral infections. The height (number at the top) of each bar specifies the relative frequency (total number) of the given sample type.

with IMX-BVN-3's intended use. As shown in Suppl. Figure 36, IMX-BVN-3 performance is significantly higher than that of SC4 in nearly all within-group comparisons, indicating that the IMX-BVN-3 scores are competitive in both classification scenarios.

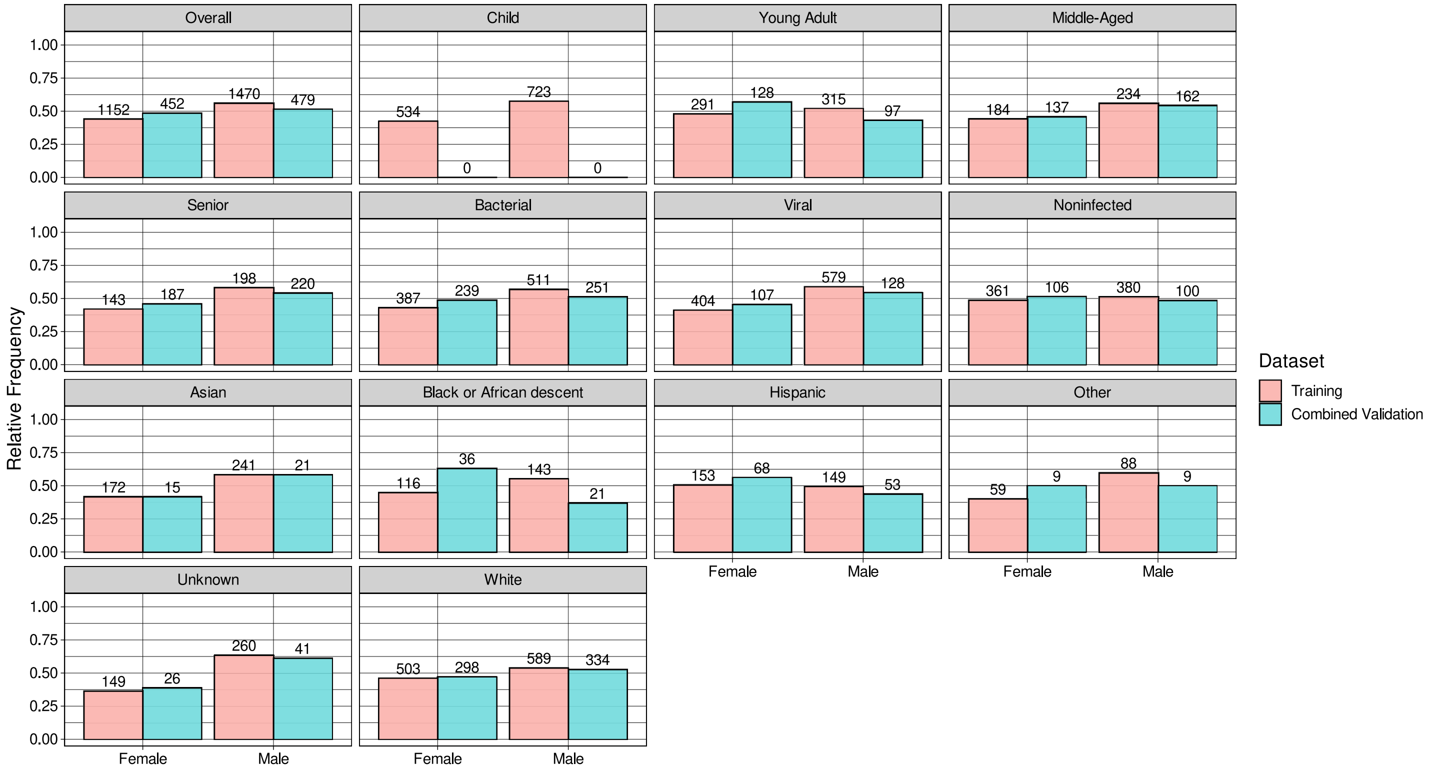


Supplementary Figure 2: **Breakdown of demographic composition of datasets of acute infection patients by sex**. The composition of our datasets is largely similar with respect to sex across nearly all patient subgroups. Although, we do note that there is a slight skew towards male patients in the training data that is reflected in some of the patient subgroups (e.g. older patients, infected patients, Asian patients). The height (number at the top) of each bar specifies the relative frequency (total number) of the given sample type.

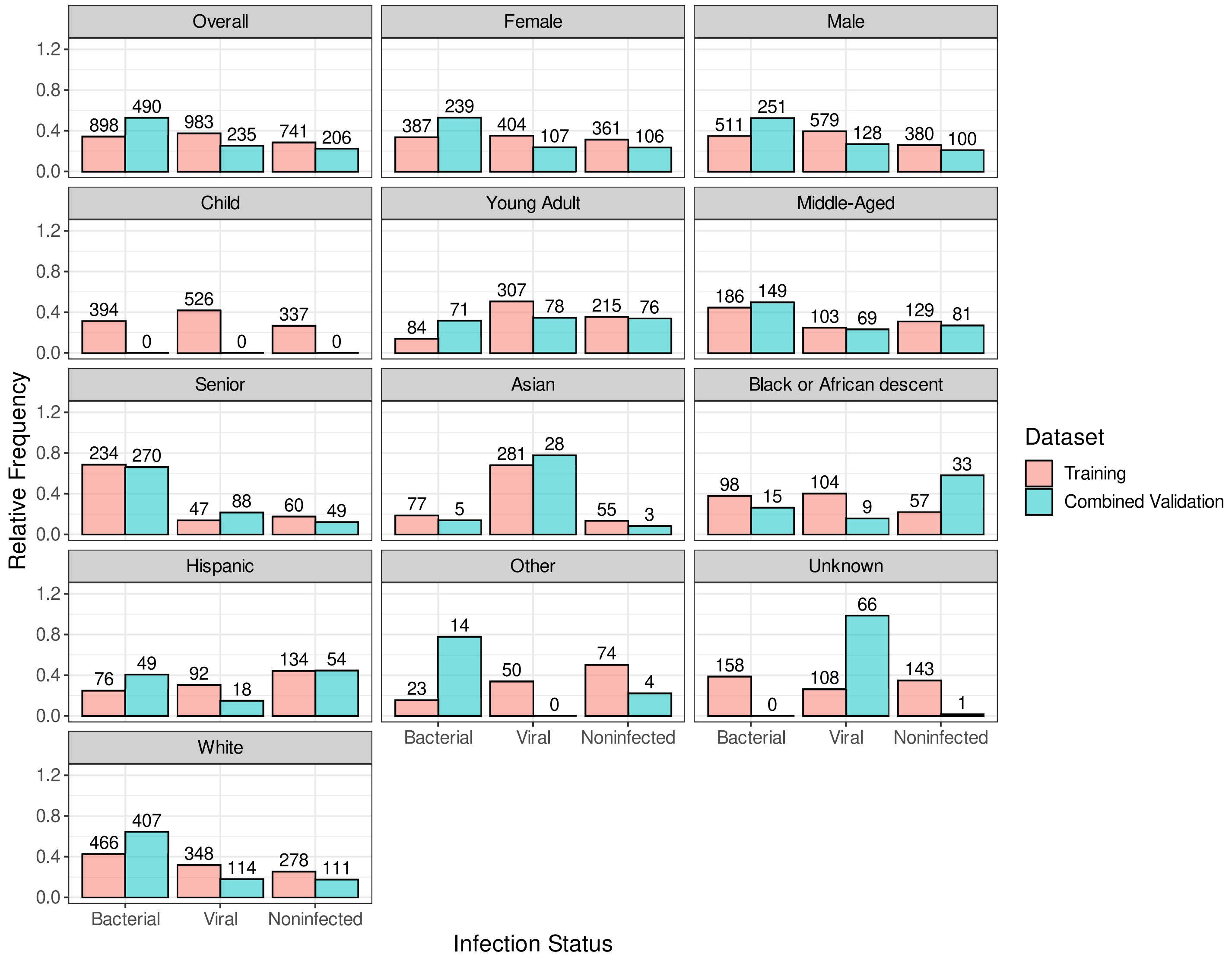


Supplementary Figure 3: **Breakdown of demographic composition of datasets of acute infection patients by infection status**. The infection status of patients was largely consistent across our training and validation datasets with some exceptions: our validation set contained relatively fewer viral patients than our training set. We also note that bacterial infected patients in our datasets skewed older while Asian patients tended to have viral infections. The height (number at the top) of each bar specifies the relative frequency (total number) of the given sample type.

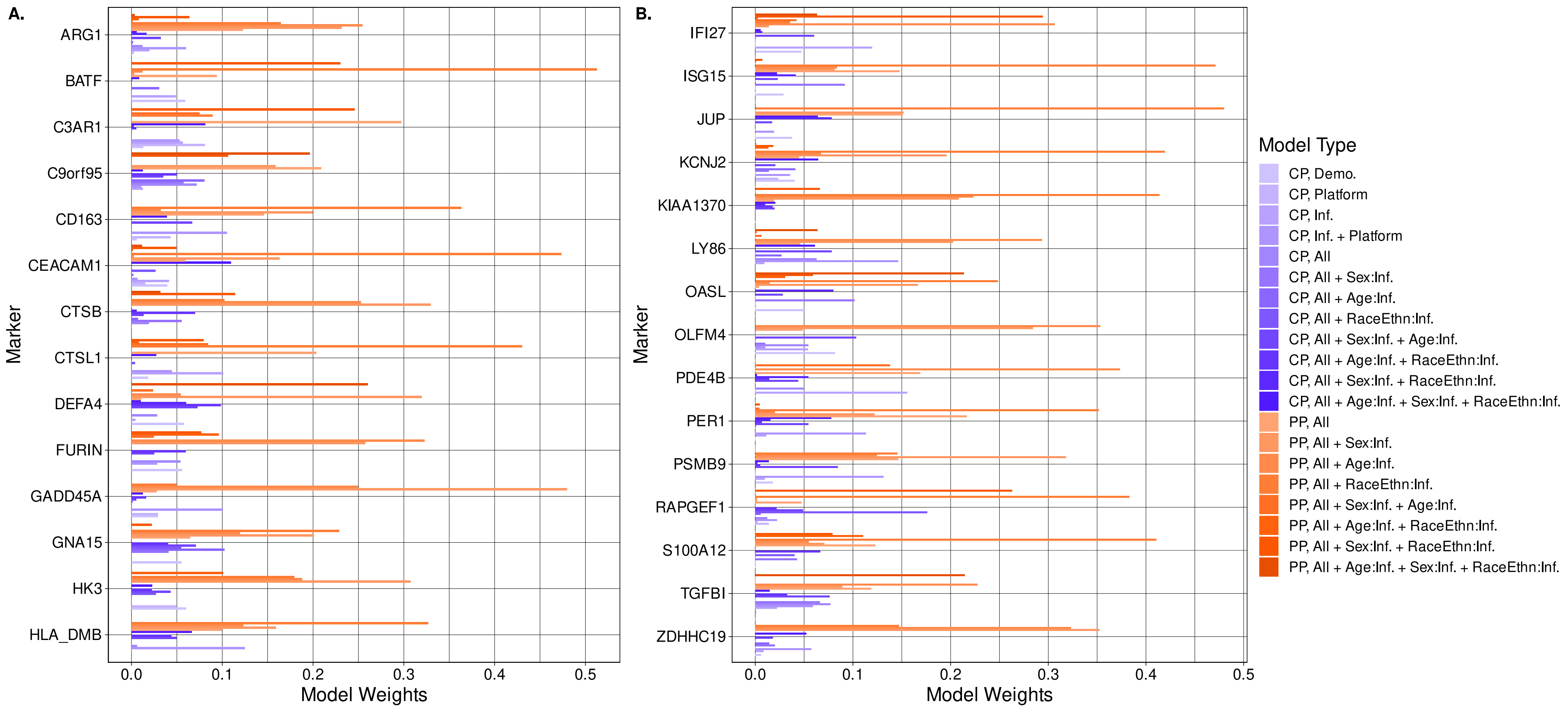


Supplementary Figure 4: **Model stacking weights from Bayesian multi-level regression modeling**. Weights indicate the relative predictive performance of different regression models of marker expression that include fixed (CP) and random (PP) effects (**A.** and **B.**) as well as interactions (e.g. sex with infection status, 'Sex:Inf.'). For every marker in the IMX-BVN-3 diagnostic panel, models with random effects for age and sex (bars colored in orange) attain the highest stacking weights indicating variability in these effects on marker expression across studies. 'Demo.' - demographic covariates race/ethnicity, age, and sex. 'Platform' - assay platform, 'Inf.' - infection status, 'All' - race/ethnicity, age, sex, assay platform, and infection status.

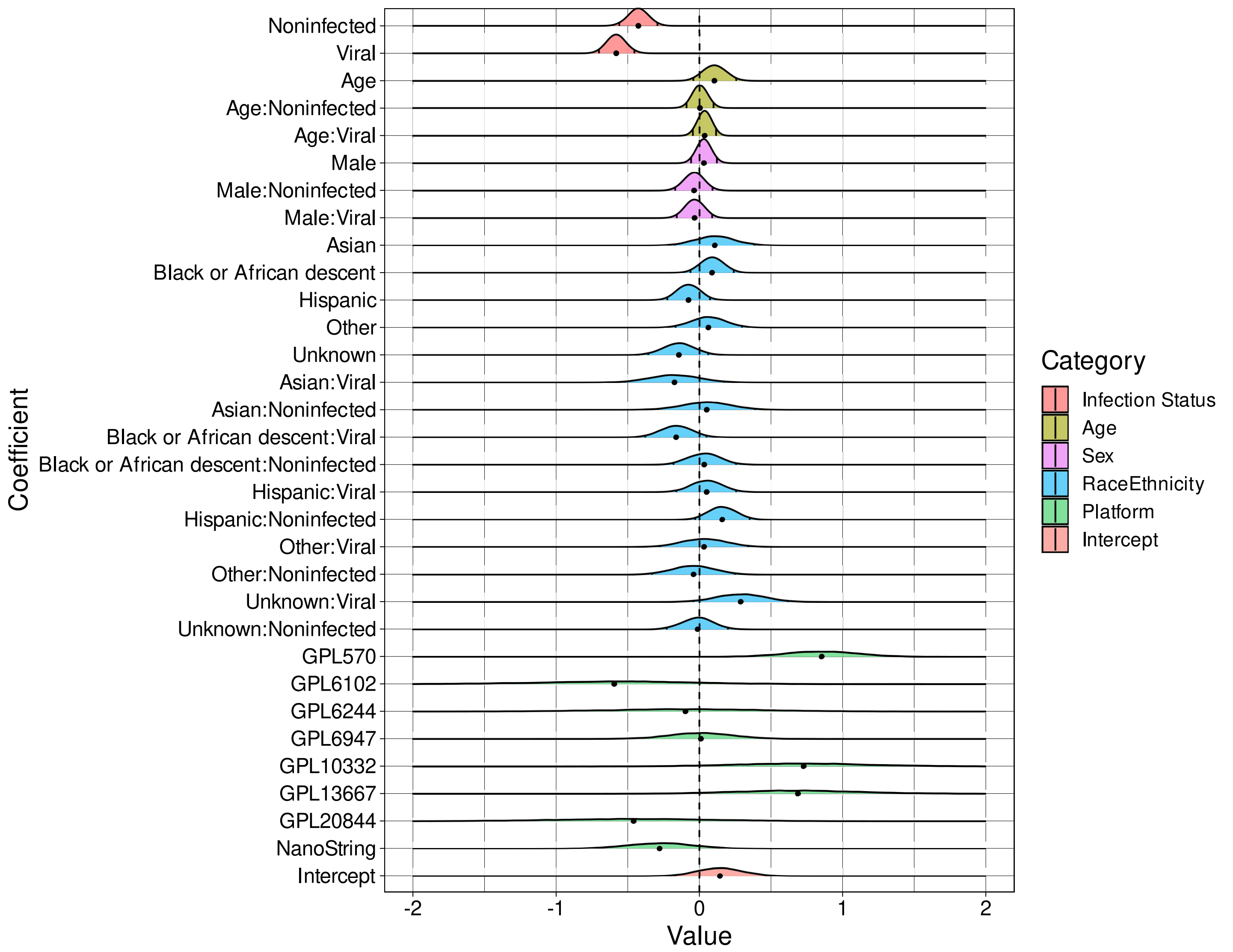


Supplementary Figure 5: **Effects inference from Bayesian multi-level model of ARG1 expression**. Shown are density plots of samples of the posterior distributions for each of the listed effects. The posterior median is indicated by the black dot at the base of the density while tick marks within the density demarcate the 2.5th and 97.5th posterior quantiles. We note significant association of ARG1 expression with infection status (density shifted from 0) though not with other patient covariates. We also detect associations between ARG1 expression and whether the sample was profiled on the GPL570 platform.

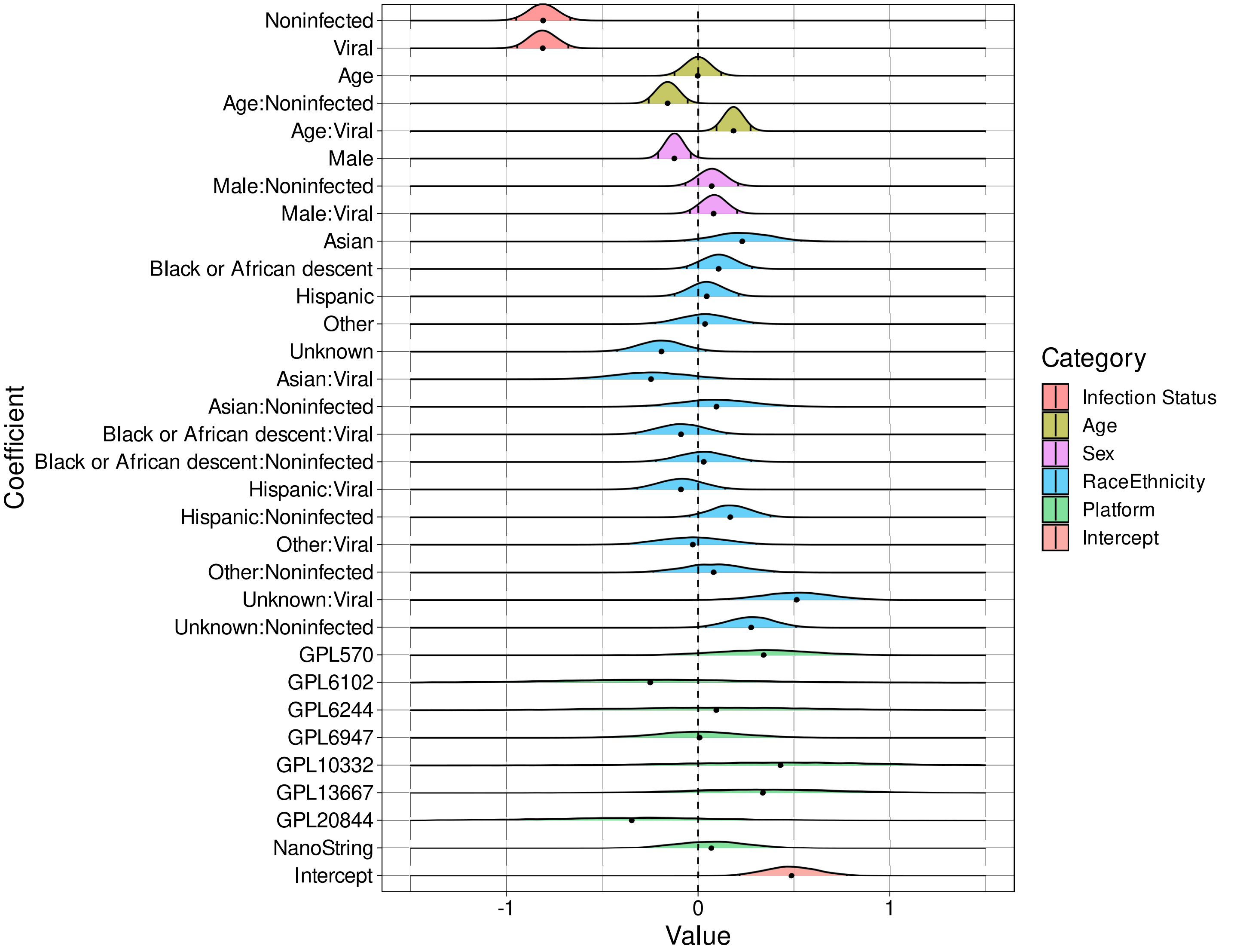


Supplementary Figure 6: **Effects inference from Bayesian multi-level model of BATF expression**. Shown are density plots of samples of the posterior distributions for each of the listed effects. The posterior median is indicated by the black dot at the base of the density while tick marks within the density demarcate the 2.5th and 97.5th posterior quantiles. We note significant association of BATF expression with infection status (density shifted from 0). We observe significant association between BATF expression and patient sex, and we note association between marker expression and the interaction of age and race/ethnicity with infection status. We do not detect associations between BATF expression and assay platform type.

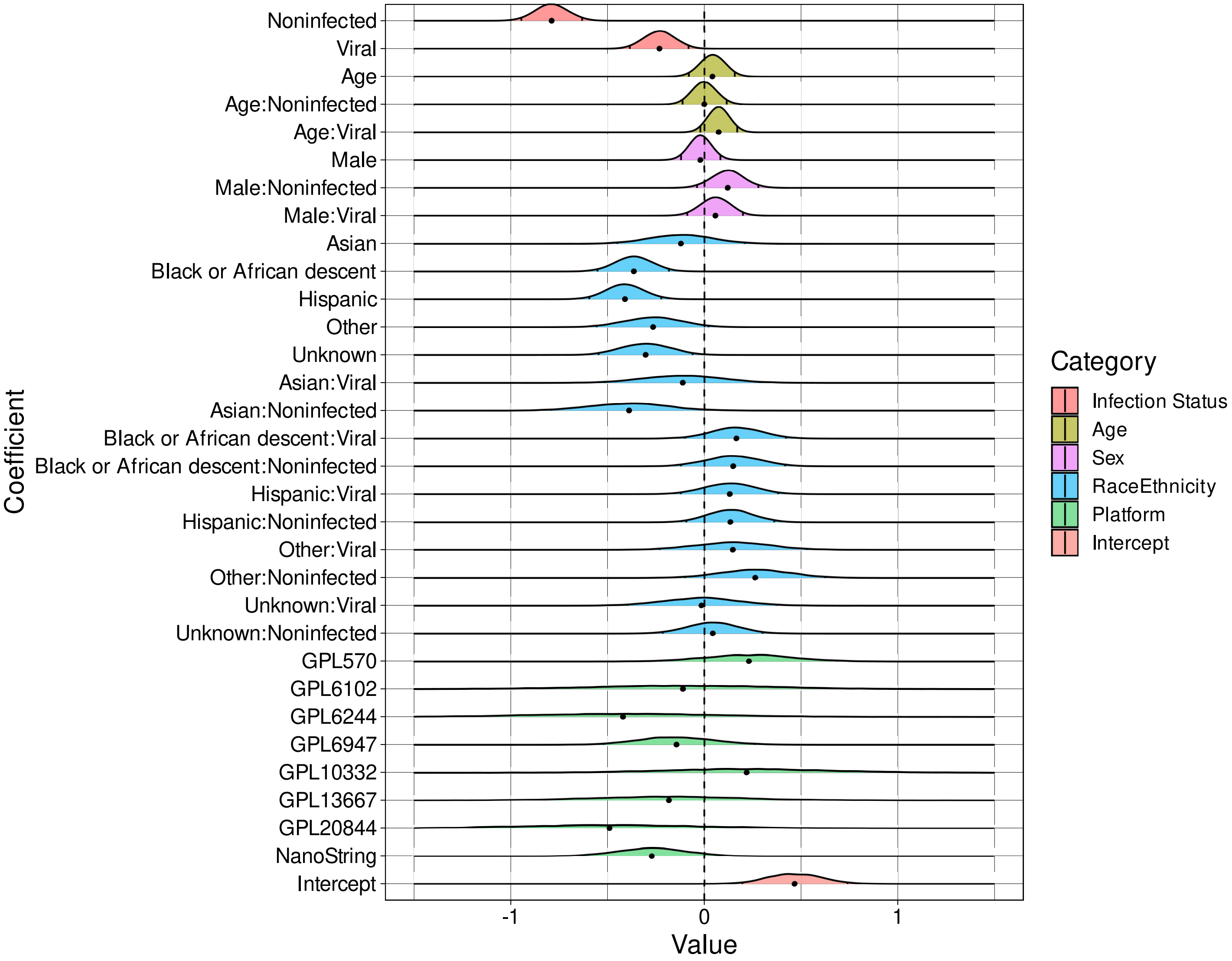


Supplementary Figure 7: **Effects inference from Bayesian multi-level model of C3AR1 expression**. Shown are density plots of samples of the posterior distributions for each of the listed effects. The posterior median is indicated by the black dot at the base of the density while tick marks within the density demarcate the 2.5th and 97.5th posterior quantiles. We note significant association of C3AR1 expression with infection status (density shifted from 0). We observe significant association between C3AR1 expression and patient race/ethnicity. We do not detect associations between C3AR1 expression and assay platform type.

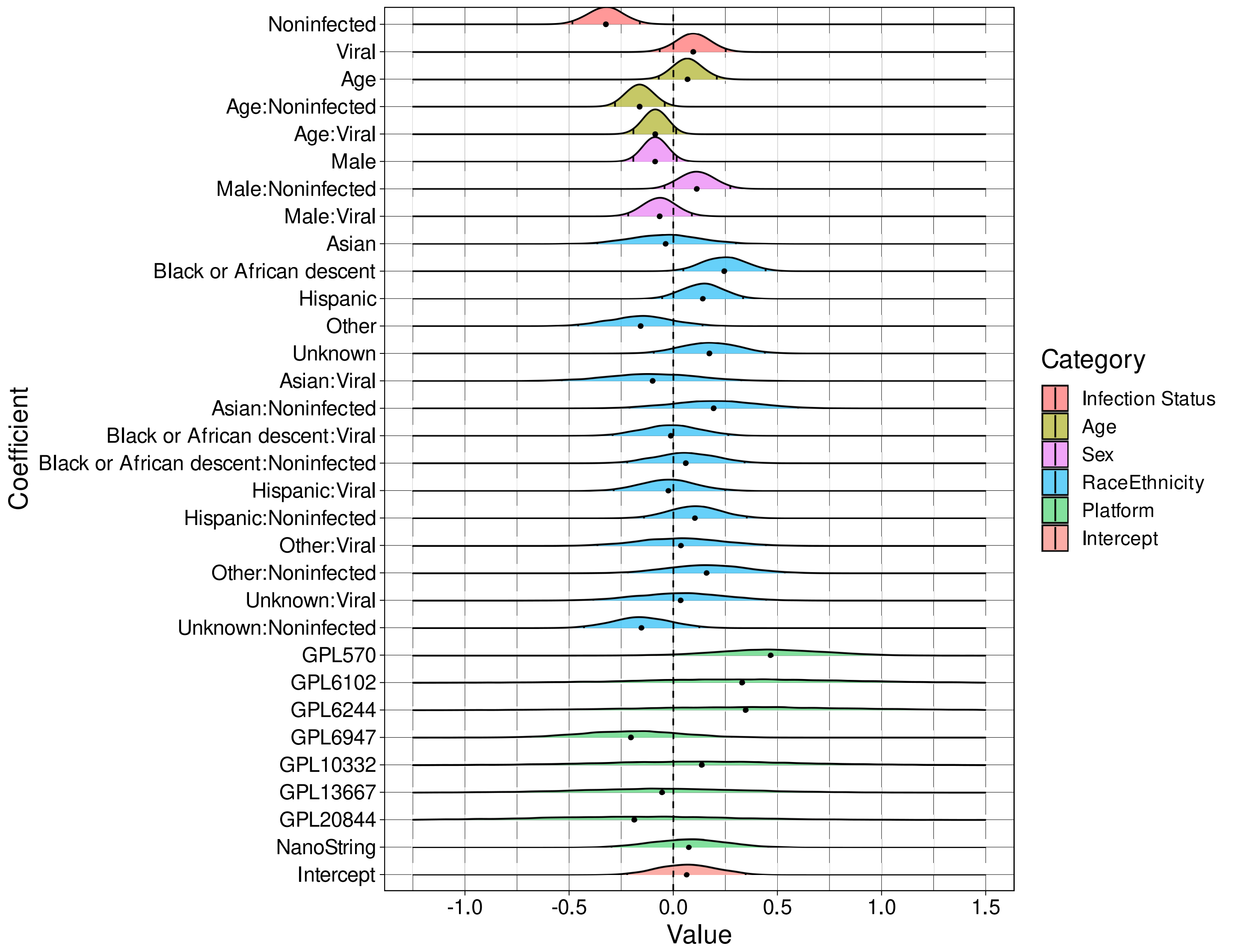


Supplementary Figure 8: **Effects inference from Bayesian multi-level model of C9orf95 expression**. Shown are density plots of samples of the posterior distributions for each of the listed effects. The posterior median is indicated by the black dot at the base of the density while tick marks within the density demarcate the 2.5th and 97.5th posterior quantiles. We note significant association of C9orf95 expression with infection status (density shifted from 0). We also observe association between C9orf95 expression and patient race/ethnicity (i.e. whether the patient is Black or of African descent) as well as between marker expression and the interaction between age and non-infected status. We do not detect associations between marker expression and assay platform type.

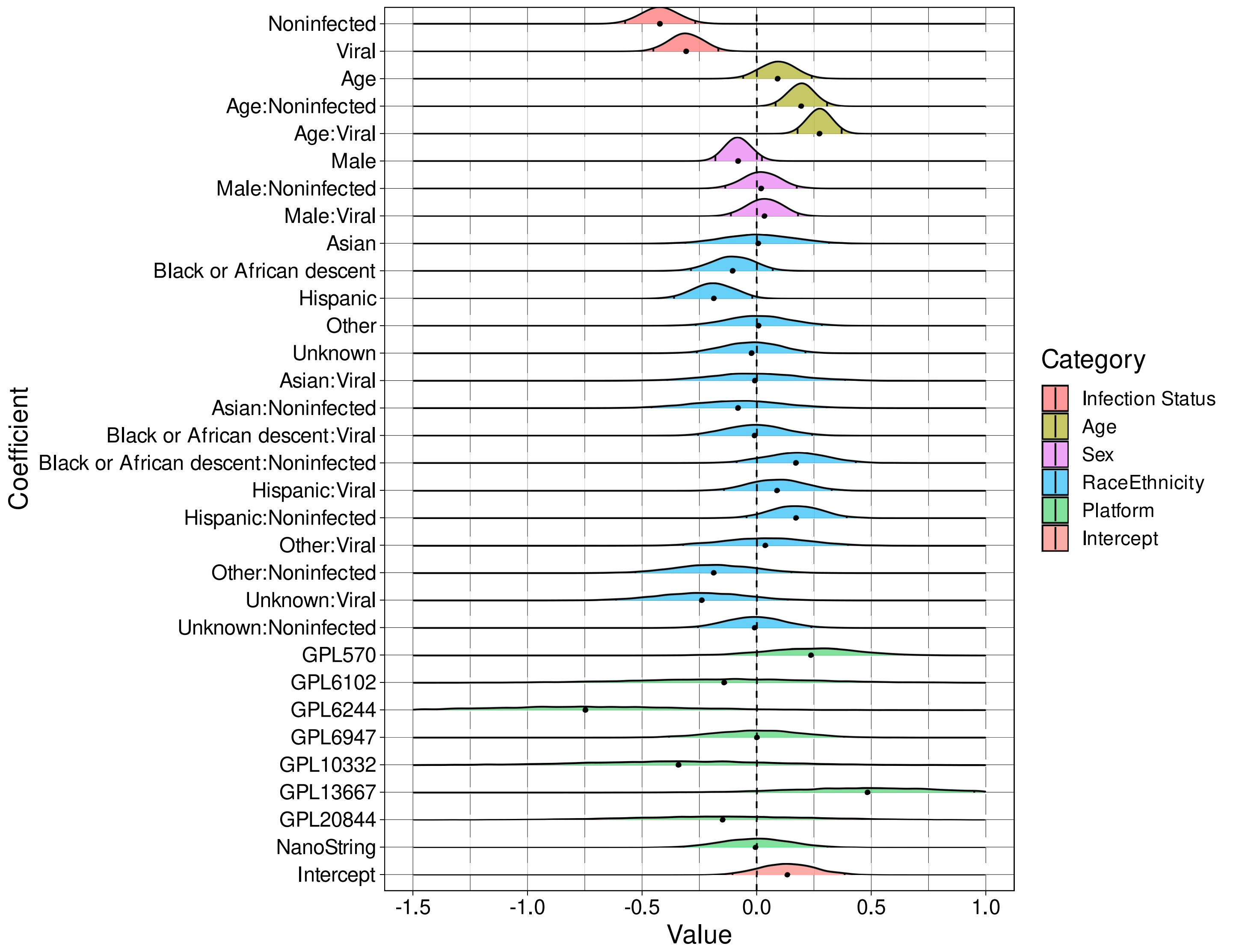


Supplementary Figure 9: **Effects inference from Bayesian multi-level model of CD163 expression**. Shown are density plots of samples of the posterior distributions for each of the listed effects. The posterior median is indicated by the black dot at the base of the density while tick marks within the density demarcate the 2.5th and 97.5th posterior quantiles. We note significant association of CD163 expression with infection status (density shifted from 0). We also observe association between CD163 expression and patient race/ethnicity (i.e. whether the patient is Hispanic) as well as between marker expression and the interaction between age and infection status. We do not detect associations between marker expression and assay platform type.

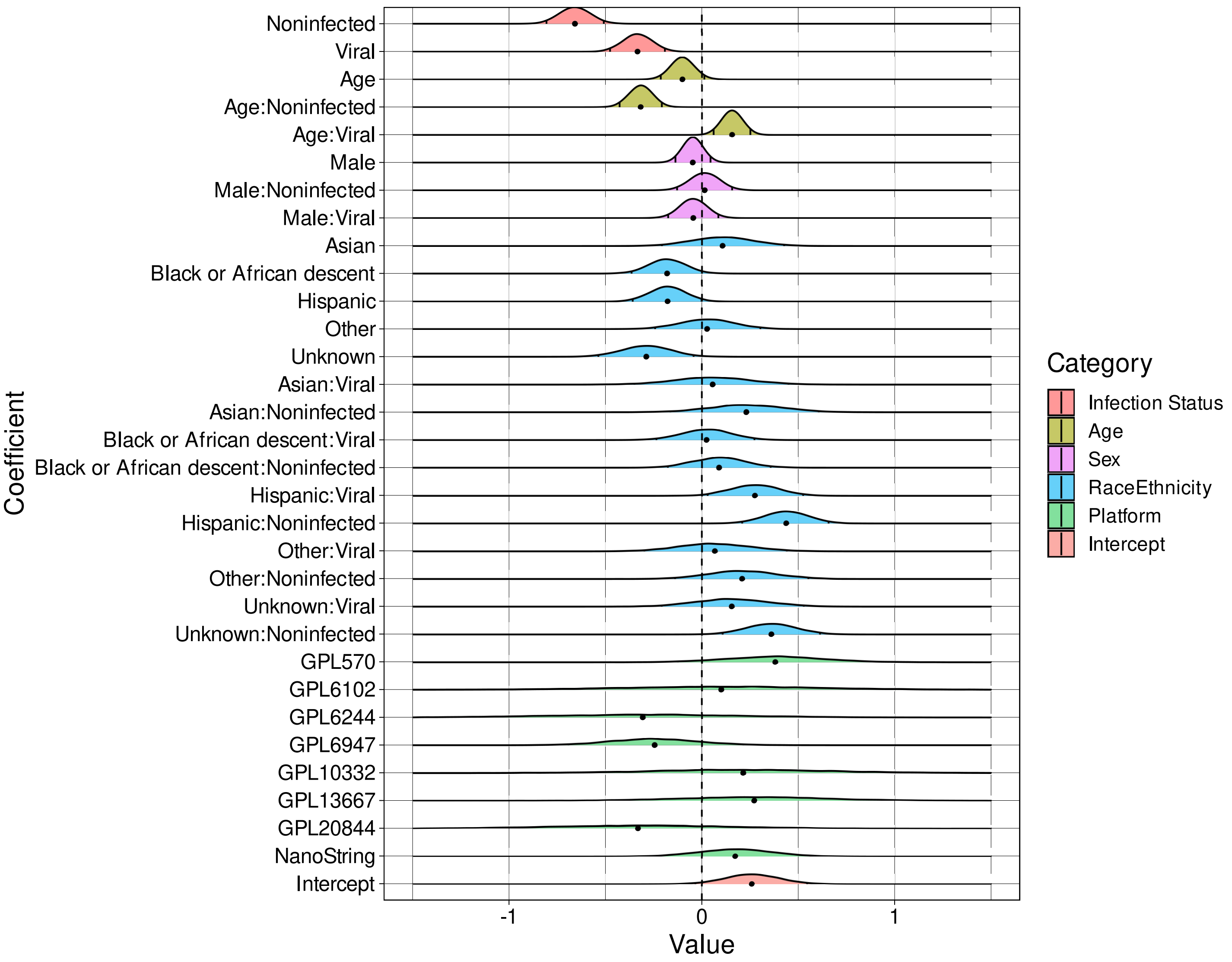


Supplementary Figure 10: **Effects inference from Bayesian multi-level model of CEACAM1 expression**. Shown are density plots of samples of the posterior distributions for each of the listed effects. The posterior median is indicated by the black dot at the base of the density while tick marks within the density demarcate the 2.5th and 97.5th posterior quantiles. We note significant association of CEACAM1 expression with infection status (density shifted from 0). We observe association between CEACAM1 expression and patient race/ethnicity. We also see significant association between marker expression and the interaction between age or race/ethnicity and infection status. We do not detect associations between marker expression and assay platform type.

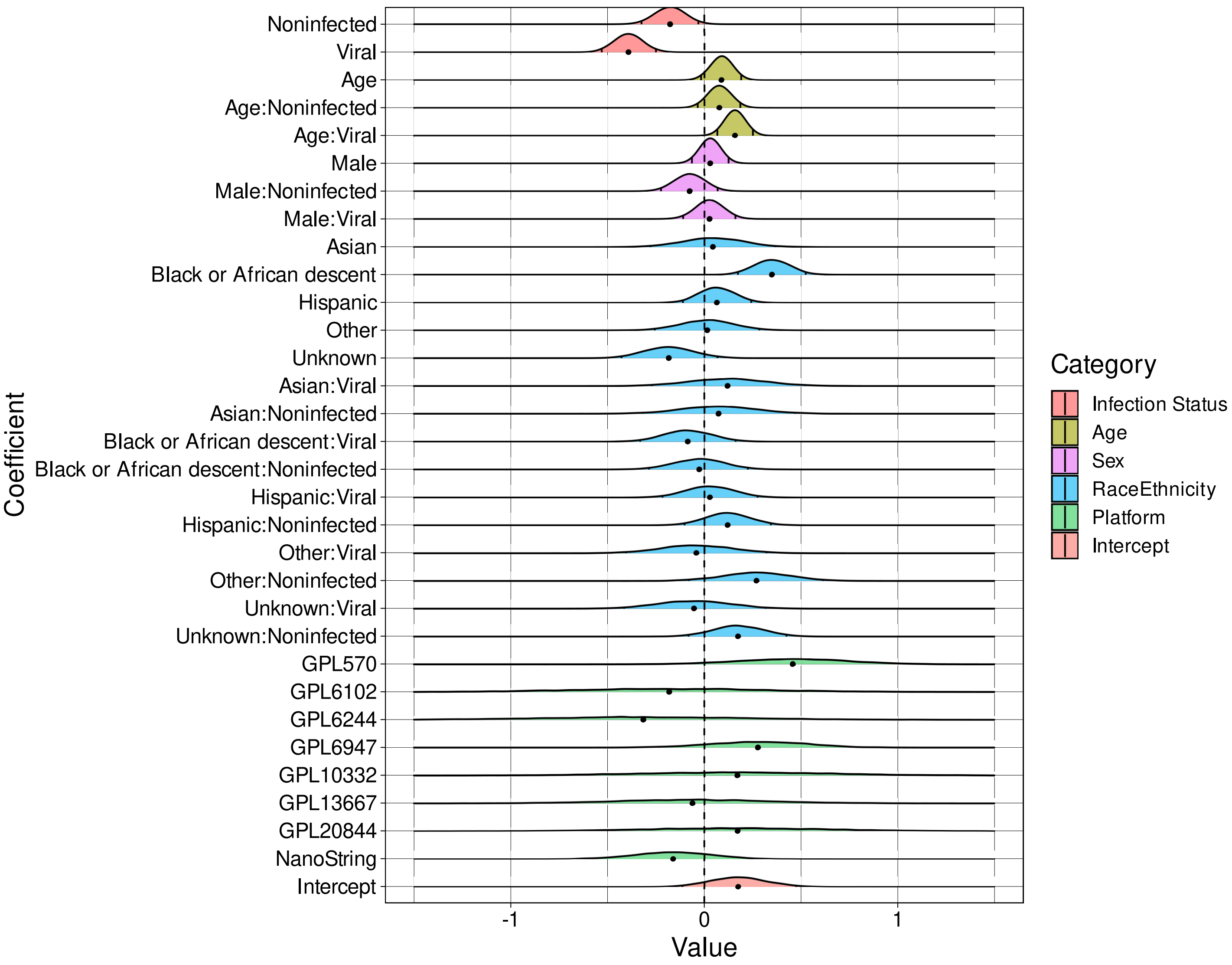


Supplementary Figure 11: **Effects inference from Bayesian multi-level model of CTSB expression**. Shown are density plots of samples of the posterior distributions for each of the listed effects. The posterior median is indicated by the black dot at the base of the density while tick marks within the density demarcate the 2.5th and 97.5th posterior quantiles. We note significant association of CTSB expression with infection status (density shifted from 0). We also detect association between CTSB expression and patient race/ethnicity (i.e. whether patient is Black or of African descent). We note association between marker expression and the interaction between age and viral infection status. We do not detect associations between marker expression and assay platform type.

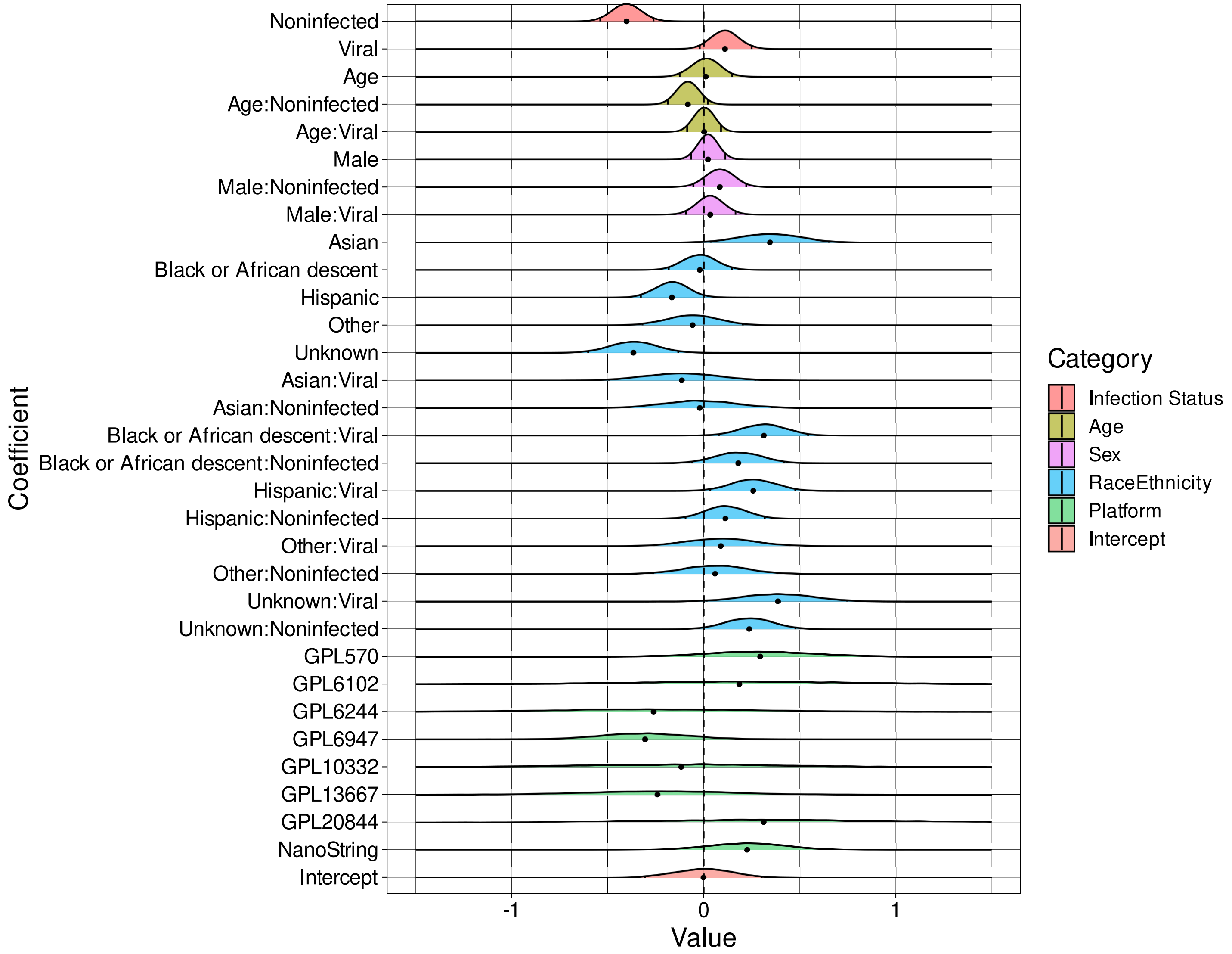


Supplementary Figure 12: **Effects inference from Bayesian multi-level model of CTSL1 expression**. Shown are density plots of samples of the posterior distributions for each of the listed effects. The posterior median is indicated by the black dot at the base of the density while tick marks within the density demarcate the 2.5th and 97.5th posterior quantiles. We note significant association of CTSL1 expression with infection status (density shifted from 0). We also observe association between CTSL1 expression and main as well as interaction effects with patient race/ethnicity. We do not detect associations between marker expression and assay platform type.

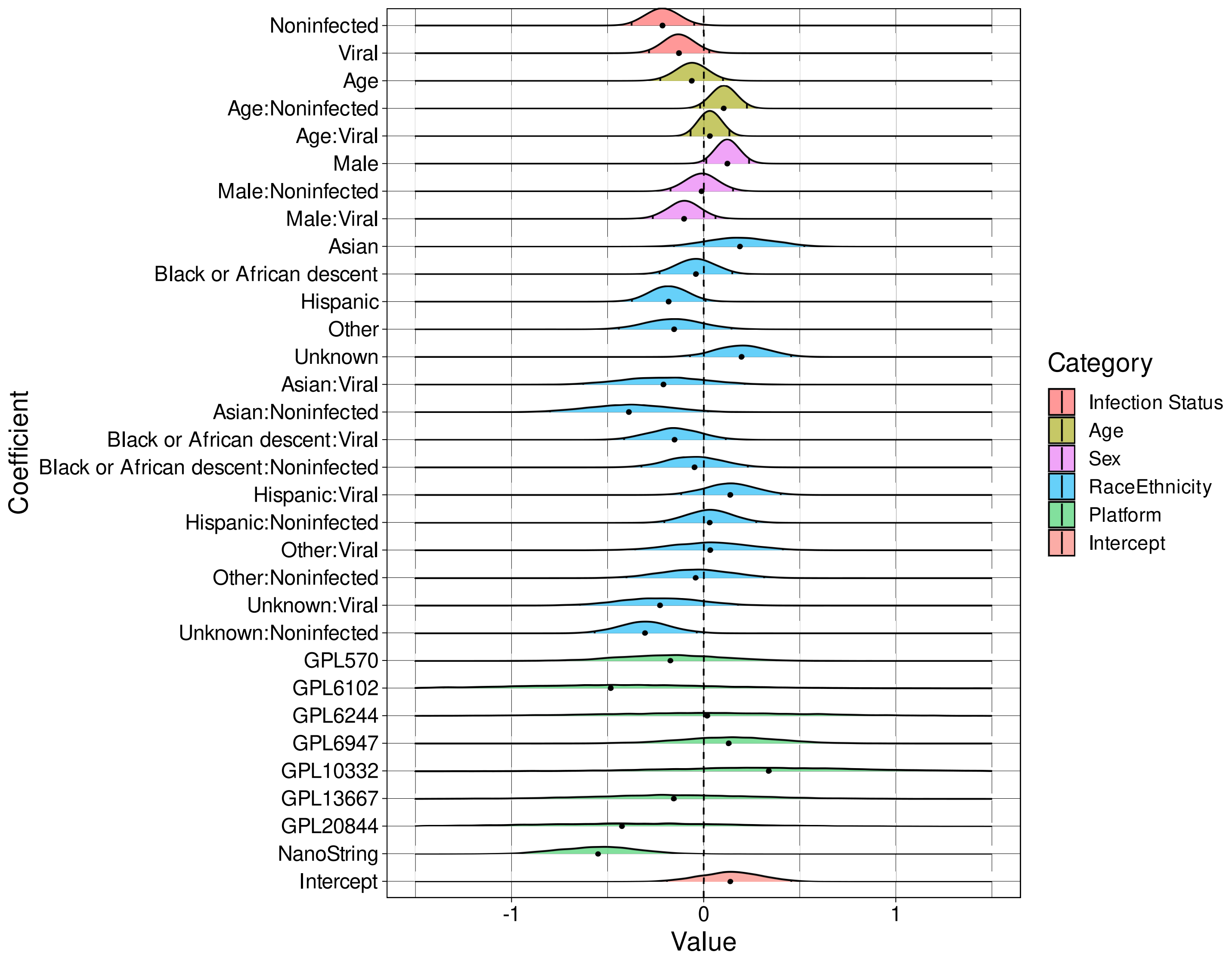


Supplementary Figure 13: **Effects inference from Bayesian multi-level model of DEFA4 expression**. Shown are density plots of samples of the posterior distributions for each of the listed effects. The posterior median is indicated by the black dot at the base of the density while tick marks within the density demarcate the 2.5th and 97.5th posterior quantiles. We detect significant (albeit mild) association of DEFA4 expression with infection status (density shifted from 0). We observe association between DEFA4 expression and sex (whether the patient is male). We also detect a significant association between marker expression and whether the sample was profiled with the NanoString platform.

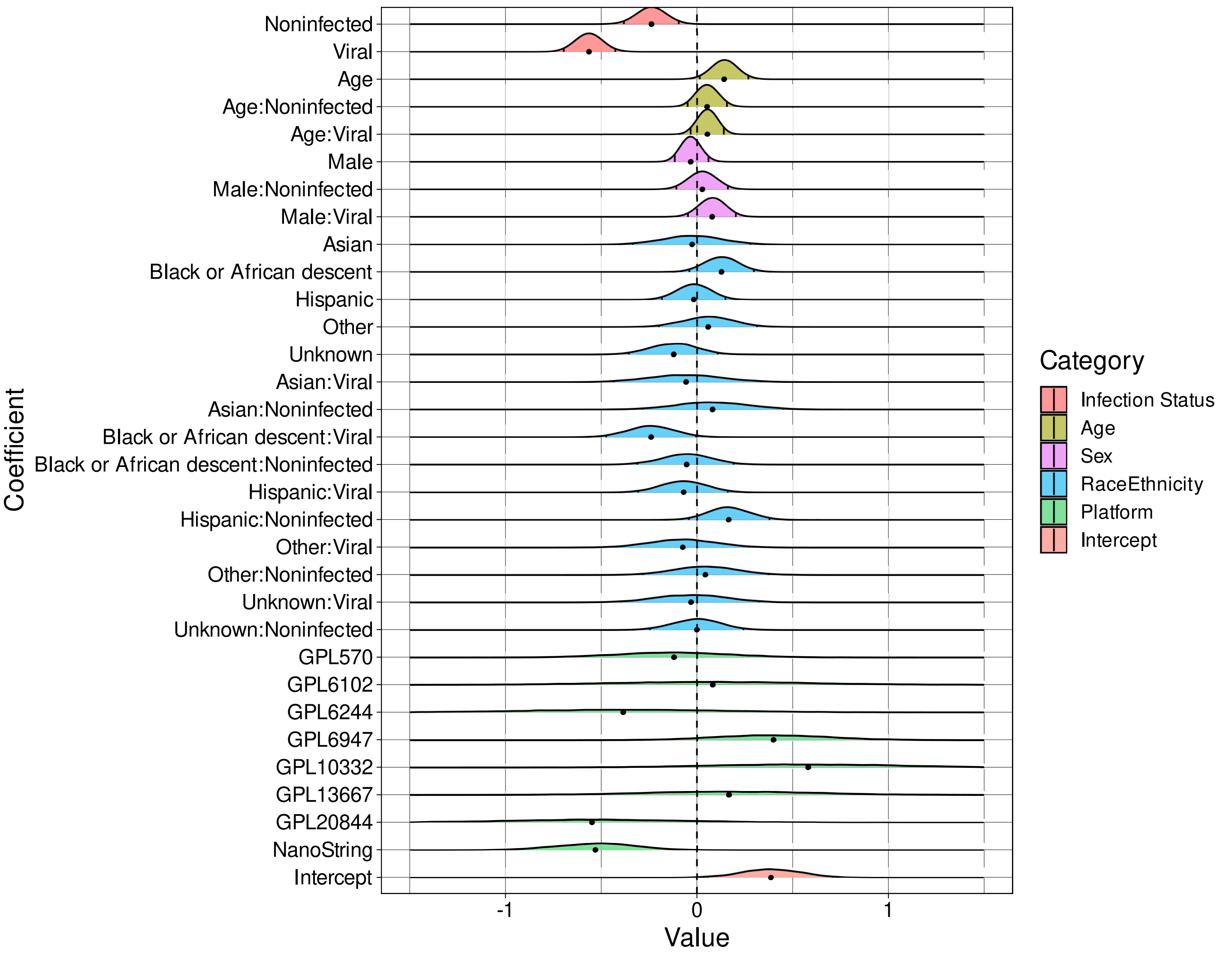


Supplementary Figure 14: **Effects inference from Bayesian multi-level model of FURIN expression**. Shown are density plots of samples of the posterior distributions for each of the listed effects. The posterior median is indicated by the black dot at the base of the density while tick marks within the density demarcate the 2.5th and 97.5th posterior quantiles. We detect significant association of FURIN expression with infection status (density shifted from 0). We observe association between FURIN expression and age as well as the interaction between viral infection status and whether the patient is Black or African descent. We also detect a significant association between marker expression and whether the sample was profiled with the NanoString platform.

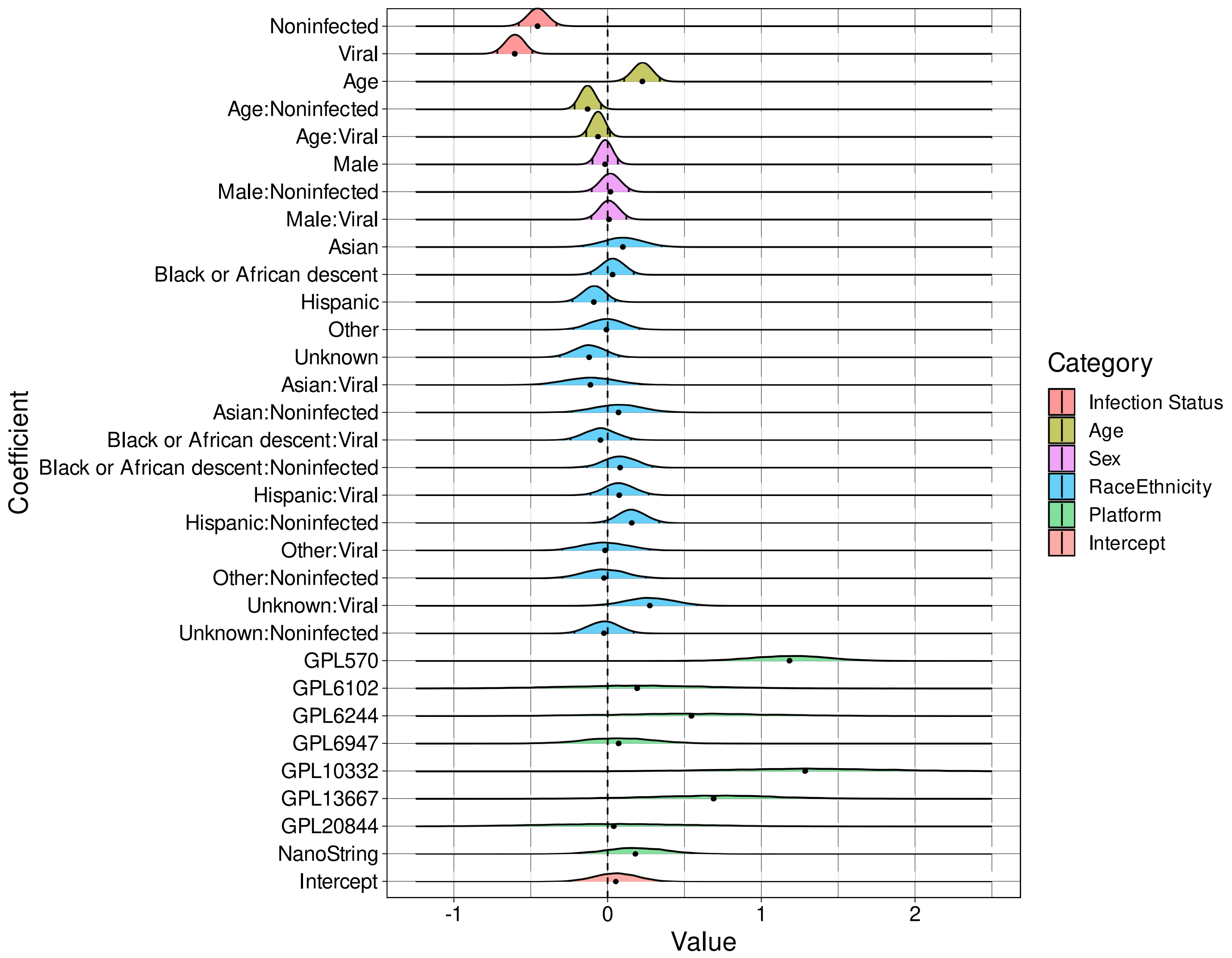


Supplementary Figure 15: **Effects inference from Bayesian multi-level model of GADD45A expression**. Shown are density plots of samples of the posterior distributions for each of the listed effects. The posterior median is indicated by the black dot at the base of the density while tick marks within the density demarcate the 2.5th and 97.5th posterior quantiles. We detect significant association of GADD45A expression with infection status (density shifted from 0). We also find association between GADD45A expression and age (both main and interaction effects). We also detect a significant association between marker expression and whether the sample was profiled with different microarray platforms (e.g. GPL570, GPL10332).

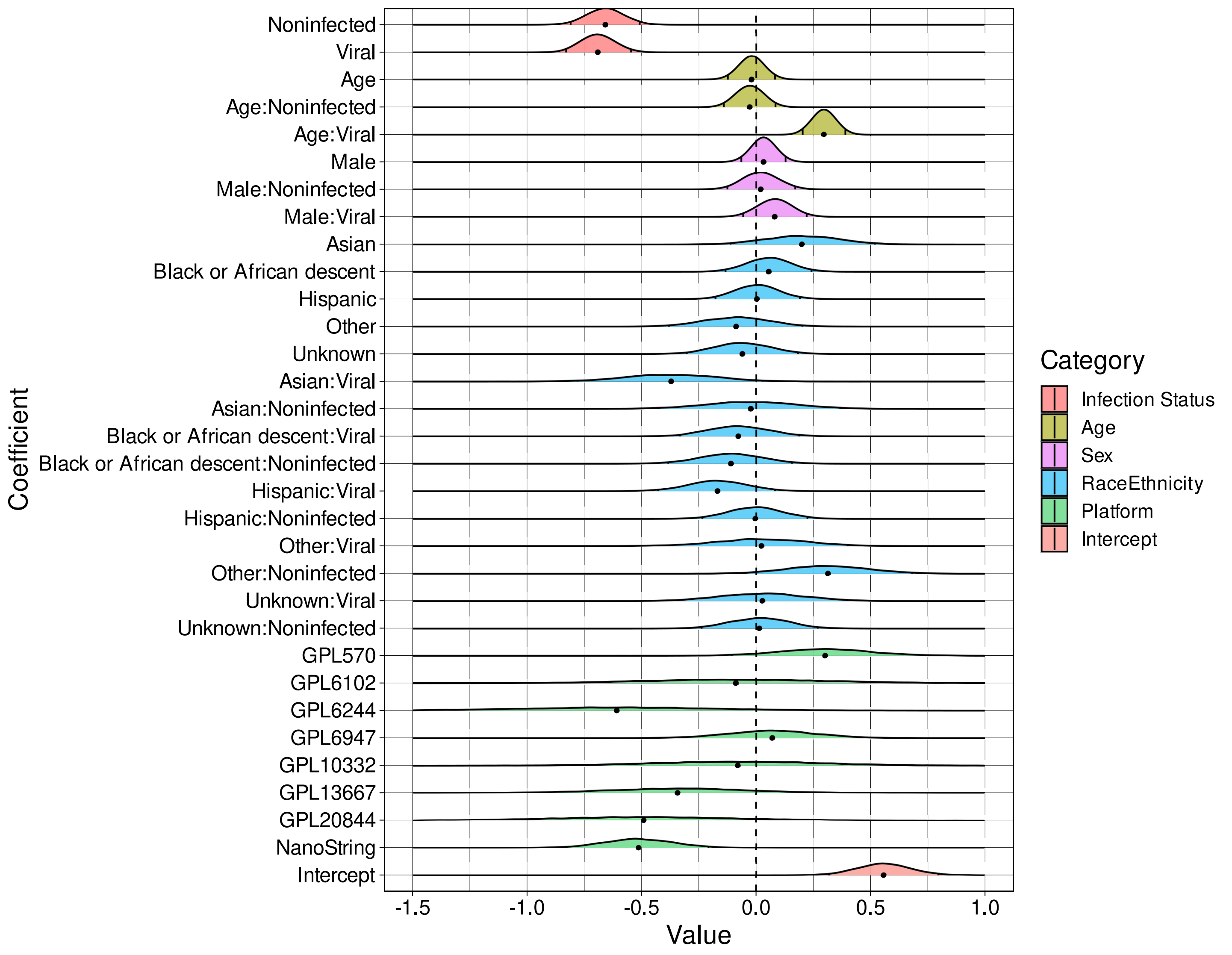


Supplementary Figure 16: **Effects inference from Bayesian multi-level model of GNA15 expression**. Shown are density plots of samples of the posterior distributions for each of the listed effects. The posterior median is indicated by the black dot at the base of the density while tick marks within the density demarcate the 2.5th and 97.5th posterior quantiles. We detect significant association of GNA15 expression with infection status (density shifted from 0). We also find association between GNA15 expression and the interaction between age and viral infection status. We also detect a significant association between marker expression and whether the sample was profiled with the NanoString platform.

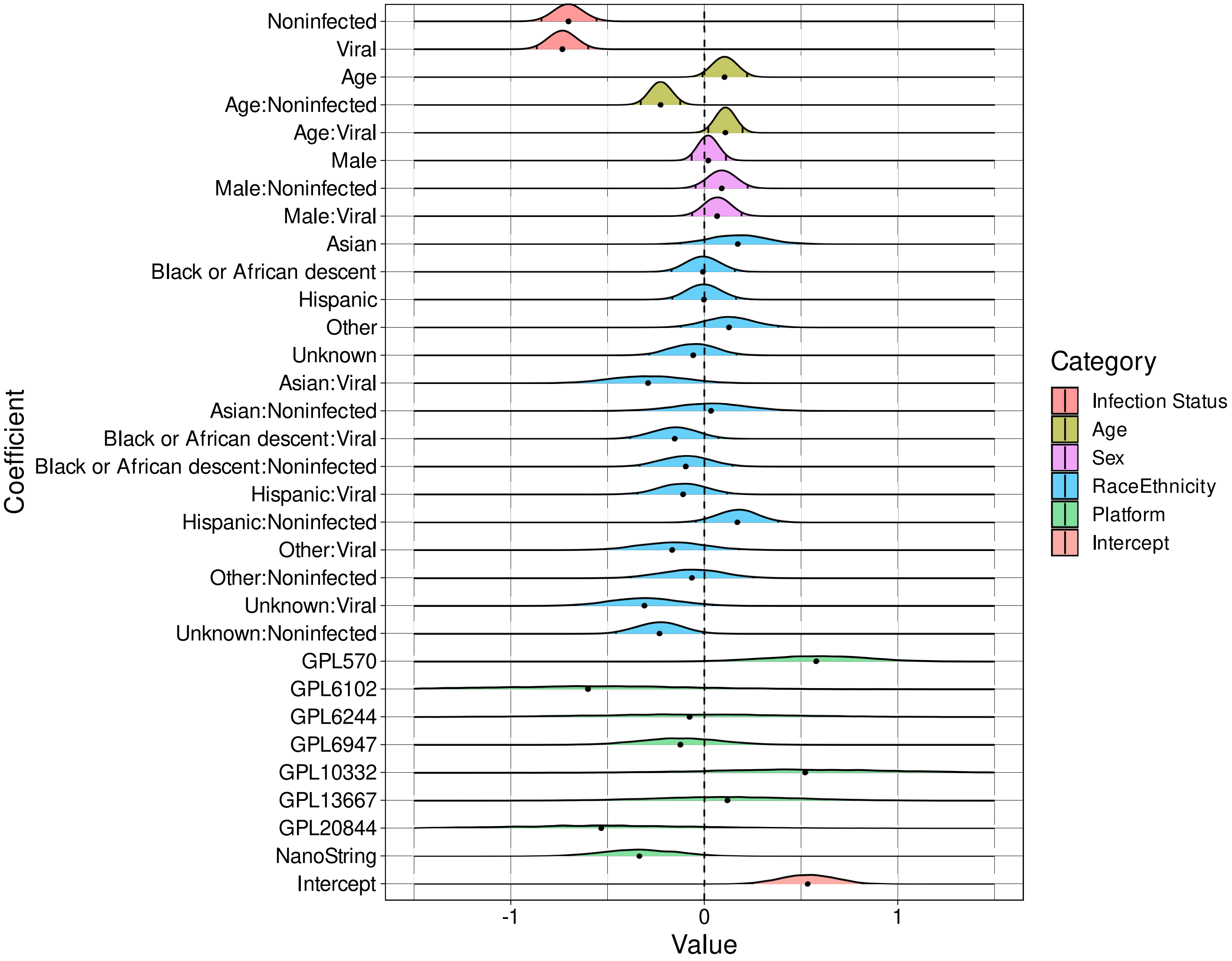


Supplementary Figure 17: **Effects inference from Bayesian multi-level model of HK3 expression**. Shown are density plots of samples of the posterior distributions for each of the listed effects. The posterior median is indicated by the black dot at the base of the density while tick marks within the density demarcate the 2.5th and 97.5th posterior quantiles. We detect significant association of HK3 expression with infection status (density shifted from 0). We also find association between HK3 expression and the interaction between age and infection status. We also detect a significant association between marker expression and whether the sample was profiled on the GPL570 microarray platform.

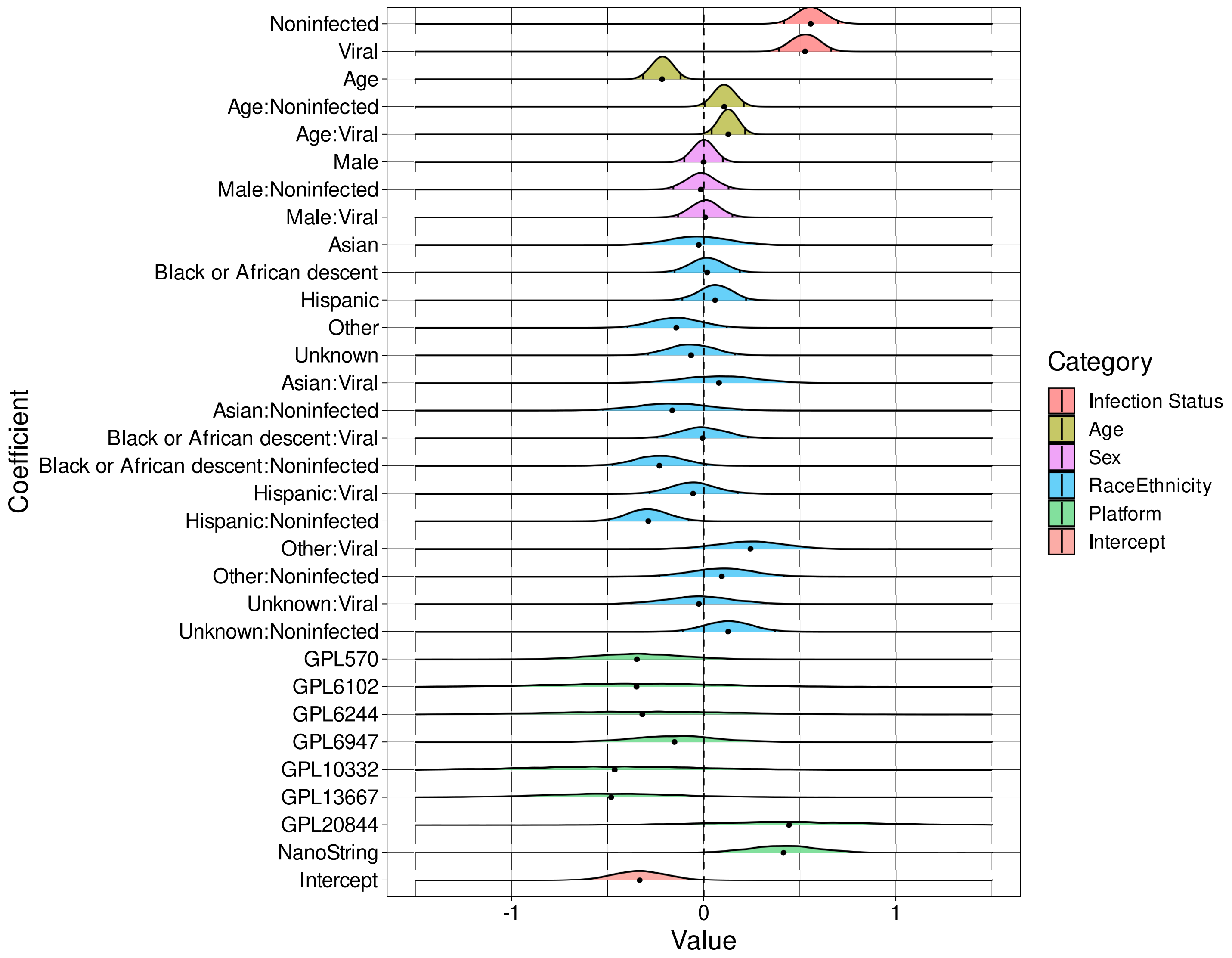


Supplementary Figure 18: **Effects inference from Bayesian multi-level model of HLA-DMB expression**. Shown are density plots of samples of the posterior distributions for each of the listed effects. The posterior median is indicated by the black dot at the base of the density while tick marks within the density demarcate the 2.5th and 97.5th posterior quantiles. We detect significant association of HLA-DMB expression with infection status (density shifted from 0). We also observe association between HLA-DMB expression and age (main and interaction effects) as well as association with the interaction between whether the patient is Hispanic and non-infected. We also note a significant association between marker expression and whether the sample was profiled on the NanoString platform.

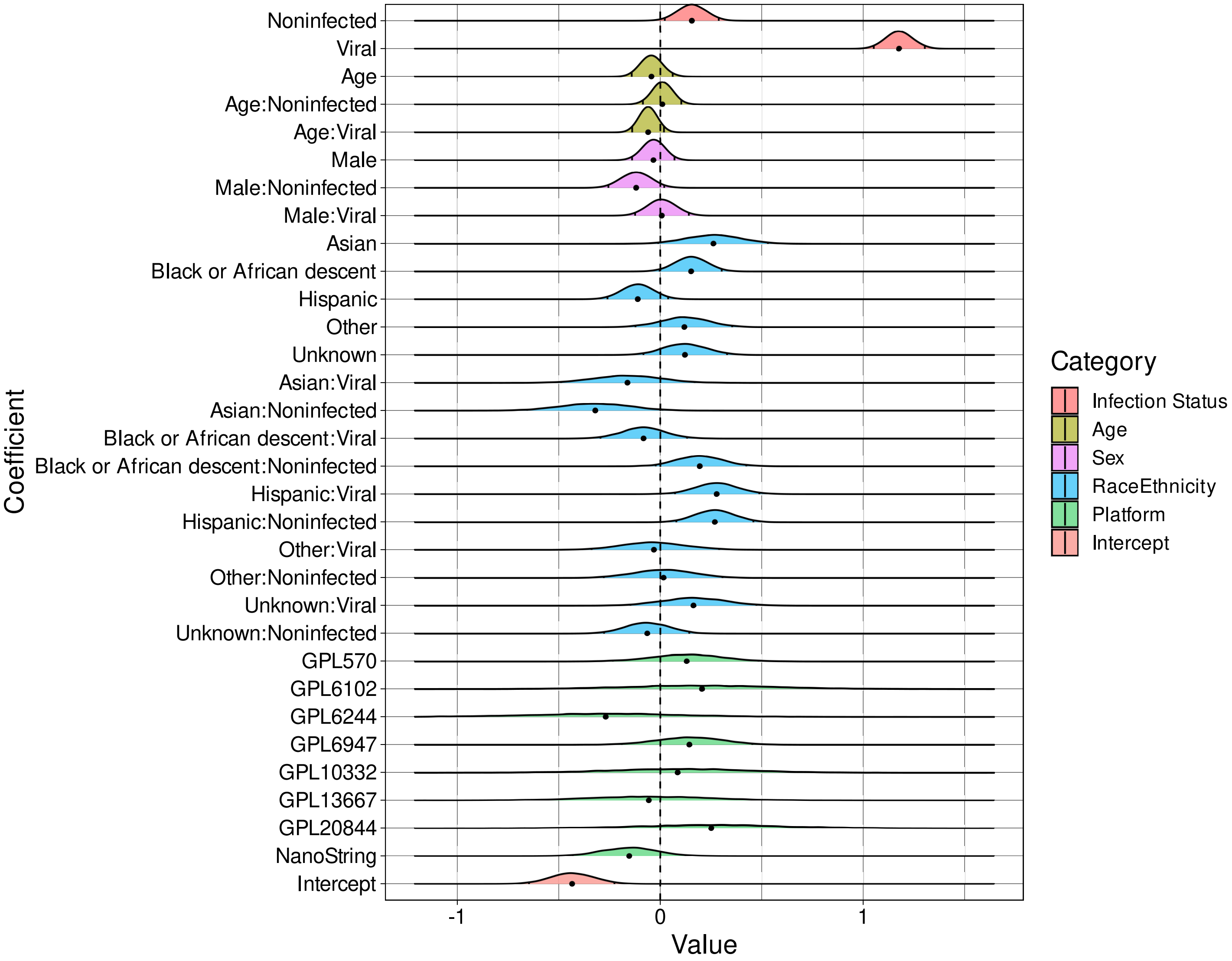


Supplementary Figure 19: **Effects inference from Bayesian multi-level model of IFI27 expression**. Shown are density plots of samples of the posterior distributions for each of the listed effects. The posterior median is indicated by the black dot at the base of the density while tick marks within the density demarcate the 2.5th and 97.5th posterior quantiles. We detect significant association of IFI27 expression with infection status (density shifted from 0). In addition, we note association with the interaction of Hispanic race/ethnicity and infection status. We do not observe associations between marker expression and technical platform.

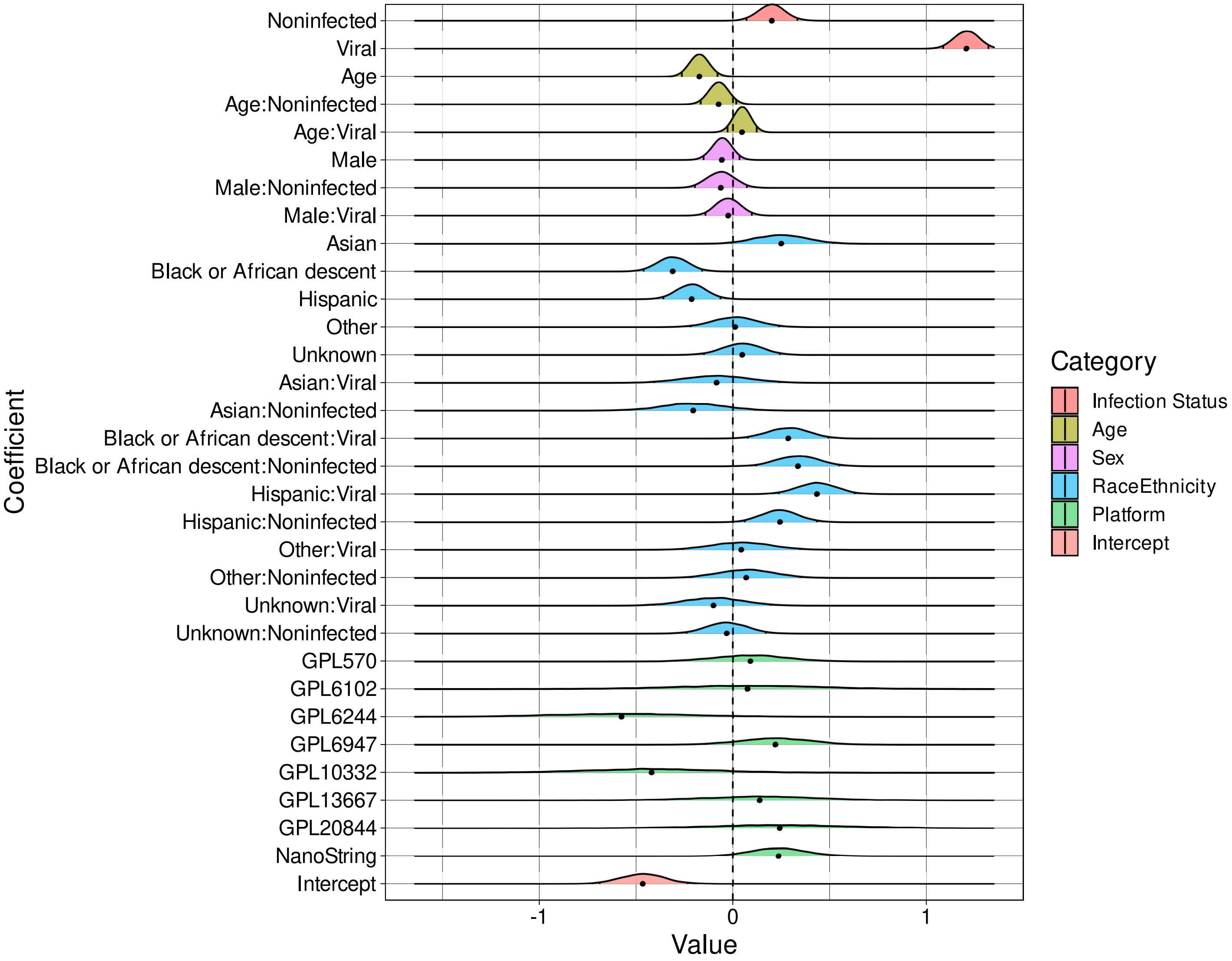


Supplementary Figure 20: **Effects inference from Bayesian multi-level model of ISG15 expression**. Shown are density plots of samples of the posterior distributions for each of the listed effects. The posterior median is indicated by the black dot at the base of the density while tick marks within the density demarcate the 2.5th and 97.5th posterior quantiles. We detect significant association of ISG15 expression with infection status (density shifted from 0) and age. We also note association with race/ethnicity (main and interaction effects). We do not observe associations between marker expression and technical platform.

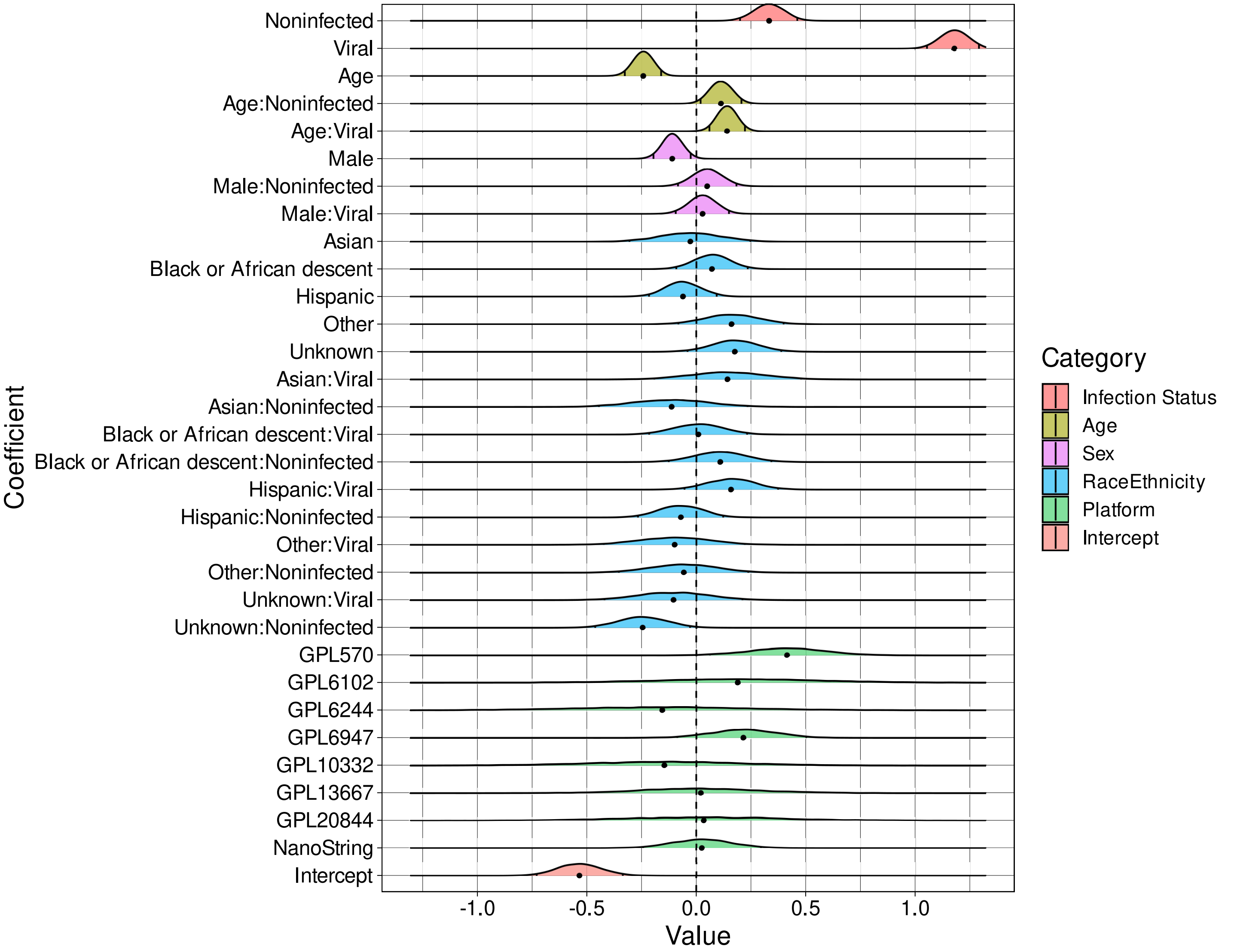


Supplementary Figure 21: **Effects inference from Bayesian multi-level model of JUP expression**. Shown are density plots of samples of the posterior distributions for each of the listed effects. The posterior median is indicated by the black dot at the base of the density while tick marks within the density demarcate the 2.5th and 97.5th posterior quantiles. We detect significant association of JUP expression with infection status (density shifted from 0) and sex. We also note association with age (main and interaction effects). We do observe associations between marker expression and whether the sample was profiled on the GPL570 microarray platform.

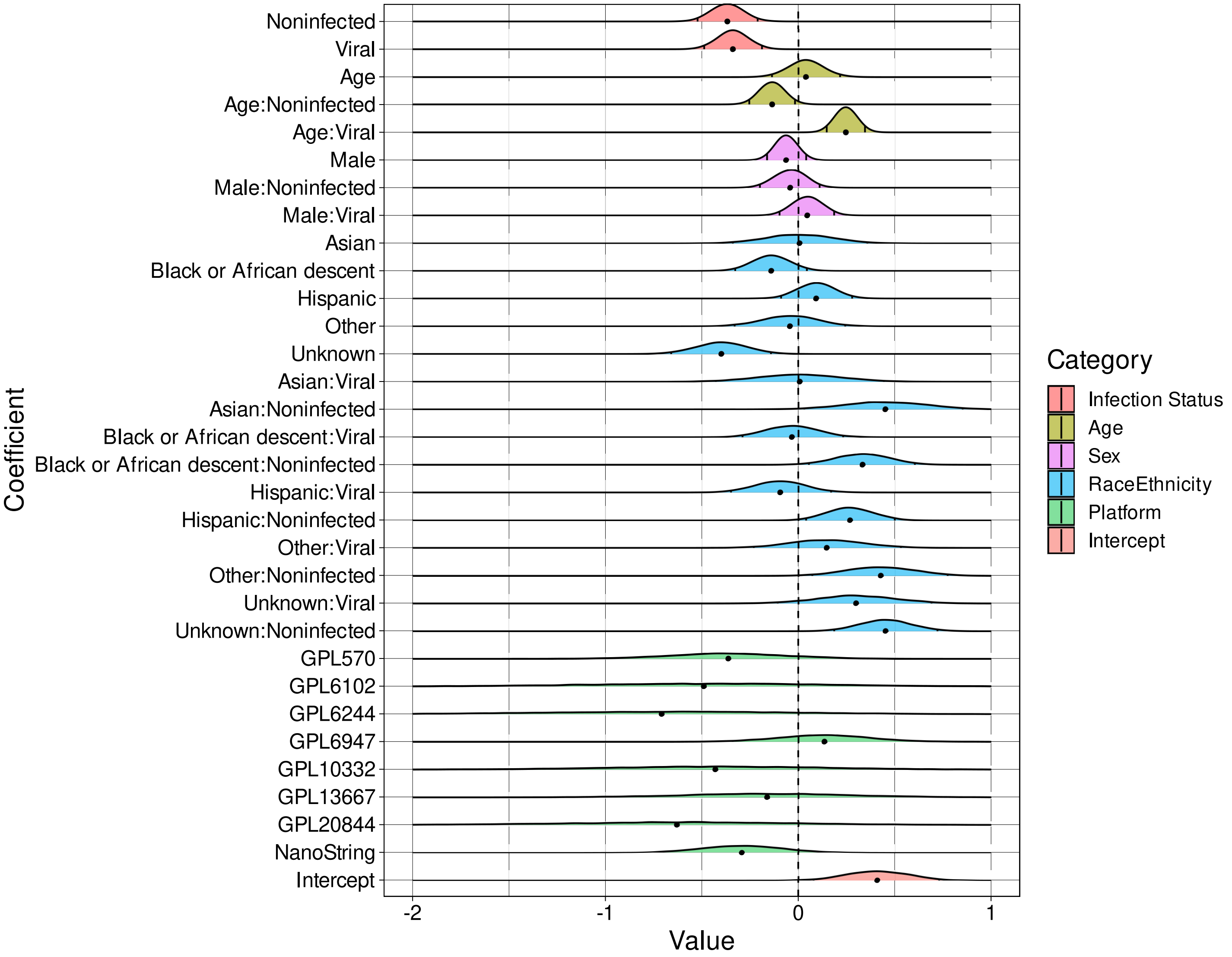


Supplementary Figure 22: **Effects inference from Bayesian multi-level model of KCNJ2 expression**. Shown are density plots of samples of the posterior distributions for each of the listed effects. The posterior median is indicated by the black dot at the base of the density while tick marks within the density demarcate the 2.5th and 97.5th posterior quantiles. We detect significant association of KCNJ2 expression with infection status (density shifted from 0) and the interactions of both age and race/ethnicity with infection status. We do not observe associations between marker expression and technical platform.

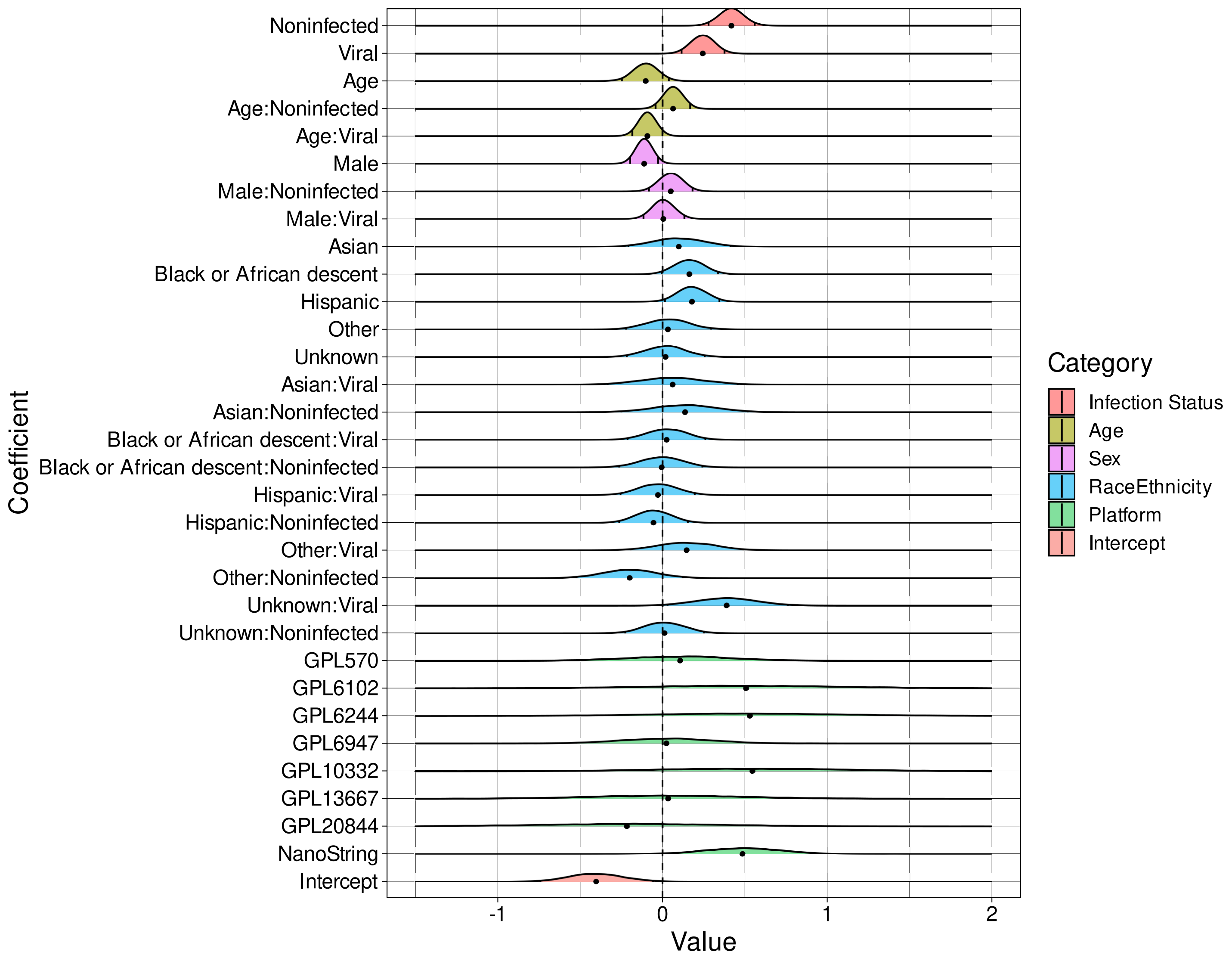


Supplementary Figure 23: **Effects inference from Bayesian multi-level model of KIAA1370 expression**. Shown are density plots of samples of the posterior distributions for each of the listed effects. The posterior median is indicated by the black dot at the base of the density while tick marks within the density demarcate the 2.5th and 97.5th posterior quantiles. We detect significant association of KIAA1370 expression with infection status (density shifted from 0) as well as with sex and race/ethnicity. We also observe significant association between marker expression and the interaction of age with viral infection. We do not observe associations between marker expression and technical platform.

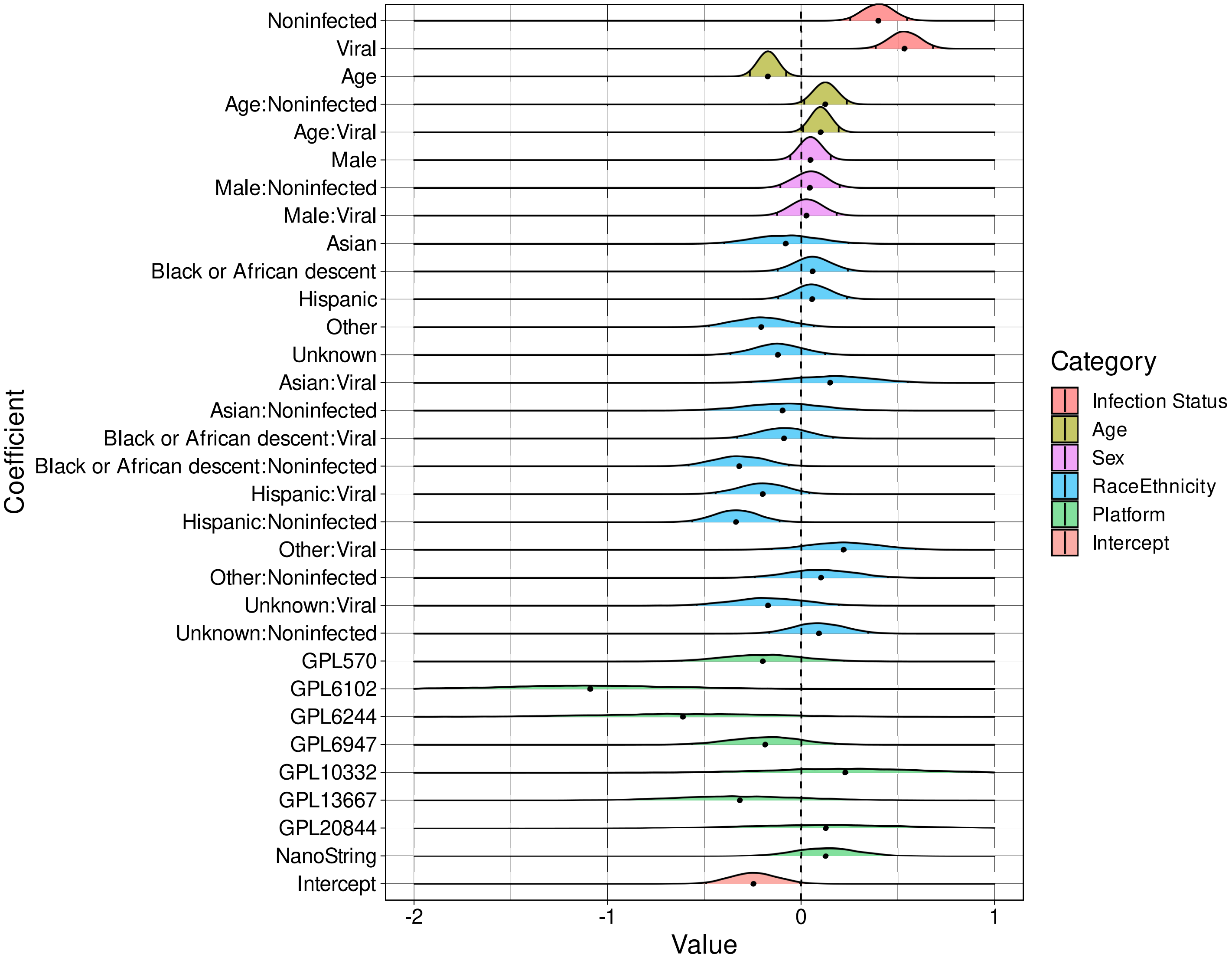


Supplementary Figure 24: **Effects inference from Bayesian multi-level model of LY86 expression**. Shown are density plots of samples of the posterior distributions for each of the listed effects. The posterior median is indicated by the black dot at the base of the density while tick marks within the density demarcate the 2.5th and 97.5th posterior quantiles. We detect significant association of LY86 expression with infection status (density shifted from 0) as well as with interactions between race/ethnicity and infection status. We also detect association with age (main and interaction effects). We do observe associations between marker expression and whether the sample was profiled on the GPL6102 microarray platform.

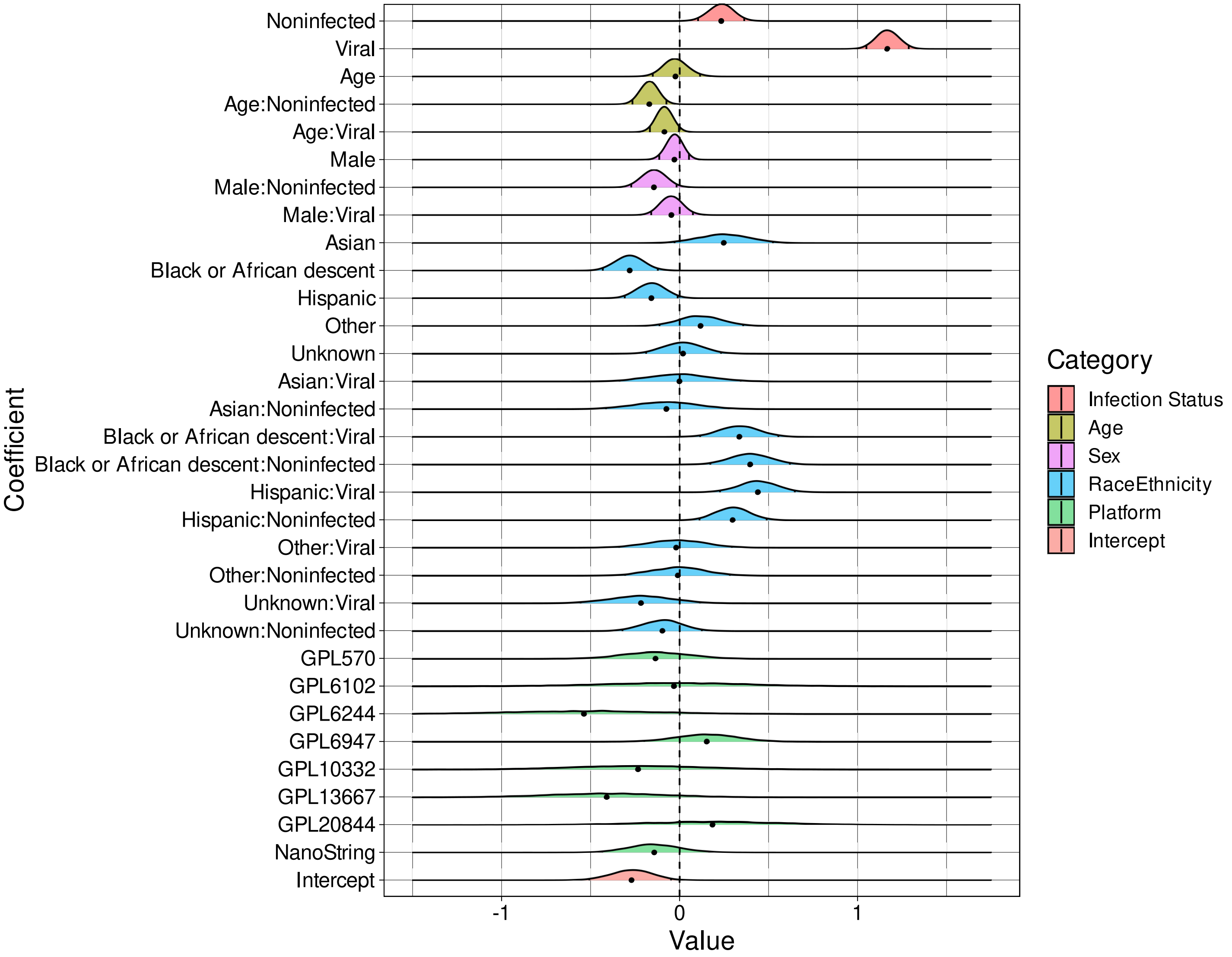


Supplementary Figure 25: **Effects inference from Bayesian multi-level model of OASL expression**. Shown are density plots of samples of the posterior distributions for each of the listed effects. The posterior median is indicated by the black dot at the base of the density while tick marks within the density demarcate the 2.5th and 97.5th posterior quantiles. We detect significant association of OASL expression with infection status (density shifted from 0) as well as with interactions between age or sex and infection status. We also detect association with race/ethnicity (main and interaction effects). We do not observe associations between marker expression and technical platform.

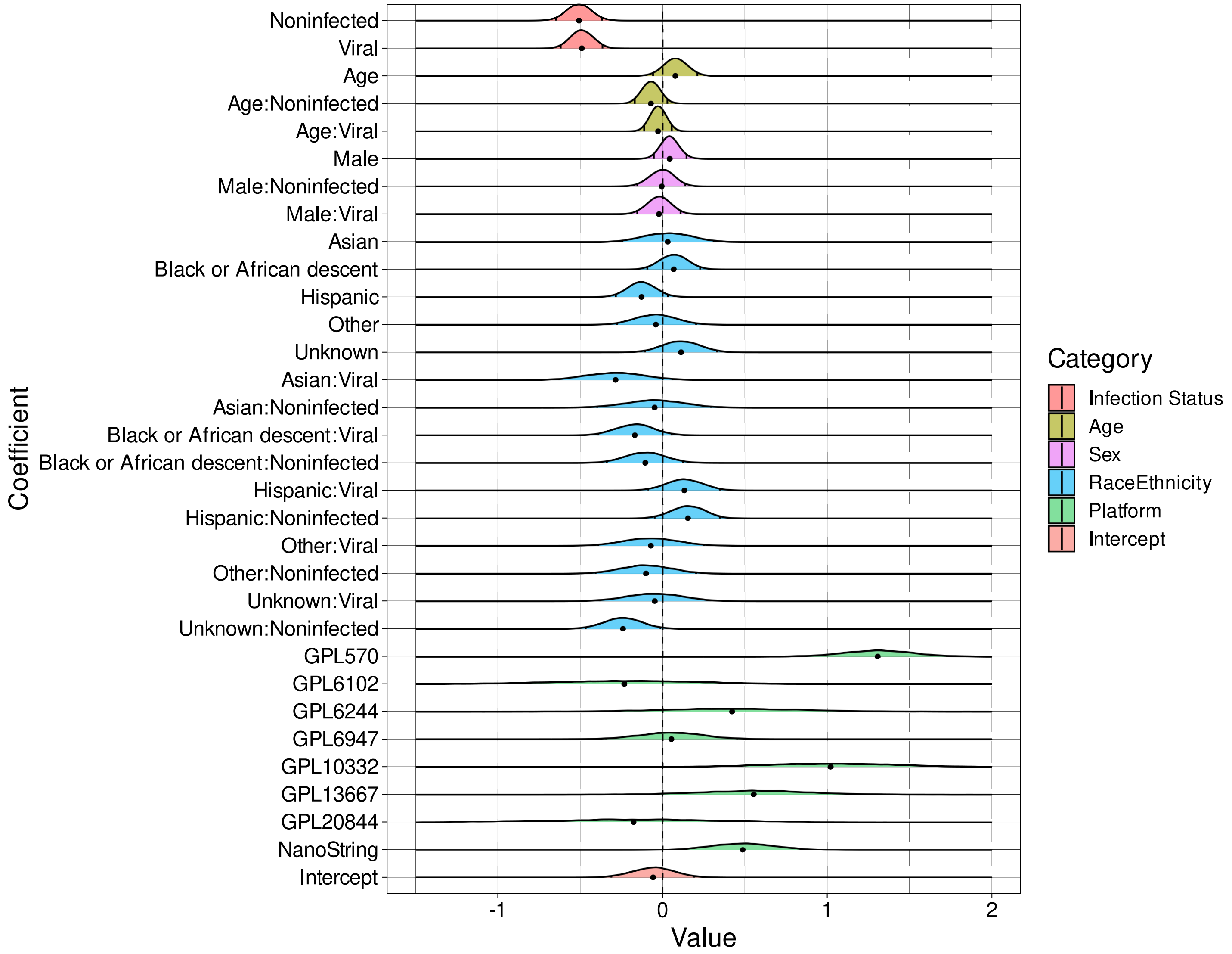


Supplementary Figure 26: **Effects inference from Bayesian multi-level model of OLFM4 expression**. Shown are density plots of samples of the posterior distributions for each of the listed effects. The posterior median is indicated by the black dot at the base of the density while tick marks within the density demarcate the 2.5th and 97.5th posterior quantiles. We detect significant association of OLFM4 expression with infection status (density shifted from 0). We also observe associations between marker expression and whether the sample was profiled on multiple platforms (GPL570, NanoString, GPL10332).

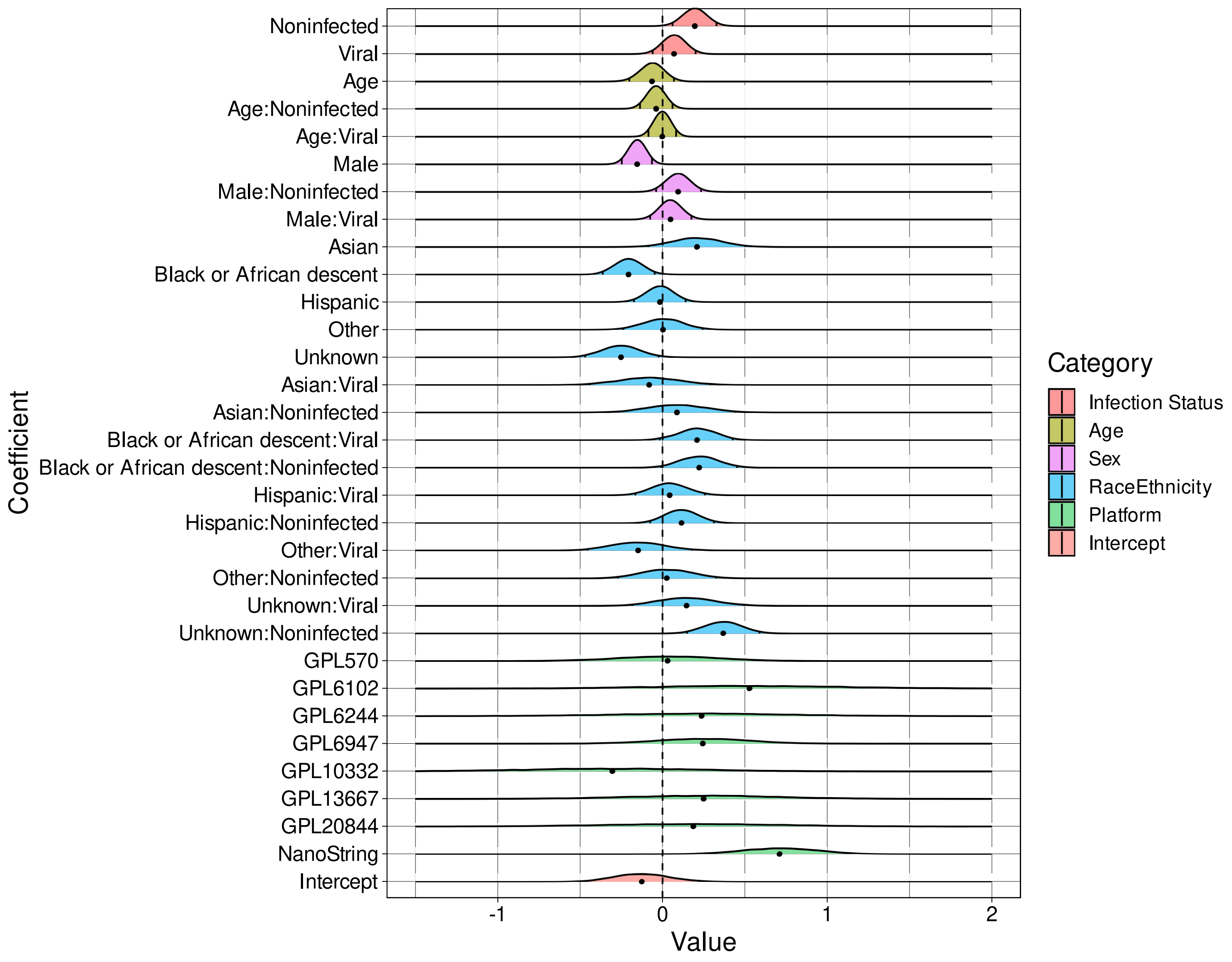


Supplementary Figure 27: **Effects inference from Bayesian multi-level model of PDE4B expression**. Shown are density plots of samples of the posterior distributions for each of the listed effects. The posterior median is indicated by the black dot at the base of the density while tick marks within the density demarcate the 2.5th and 97.5th posterior quantiles. We detect significant (albeit mild) association of PDE4B expression with infection status (density shifted from 0) as well as with sex and race/ethnicity. We also observe associations between marker expression and whether the sample was profiled on NanoString.

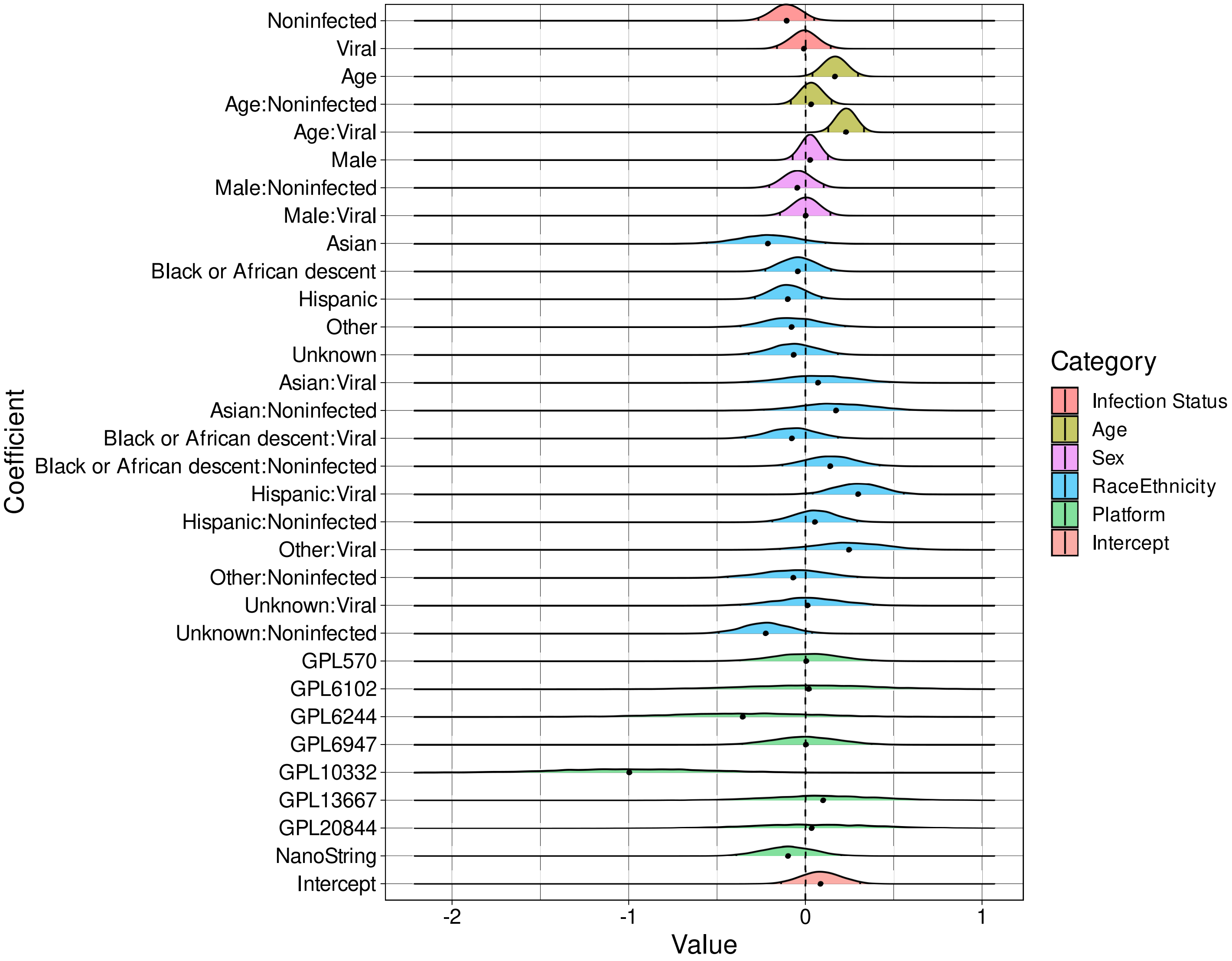


Supplementary Figure 28: **Effects inference from Bayesian multi-level model of PER1 expression**. Shown are density plots of samples of the posterior distributions for each of the listed effects. The posterior median is indicated by the black dot at the base of the density while tick marks within the density demarcate the 2.5th and 97.5th posterior quantiles. We do not detect significant association of PER1 expression with infection status. We do observe associations with age and between interactions of age or whether the patient is Hispanic with viral infection status. We also observe associations between marker expression and whether the sample was profiled on the GPL10332 microarray platform.

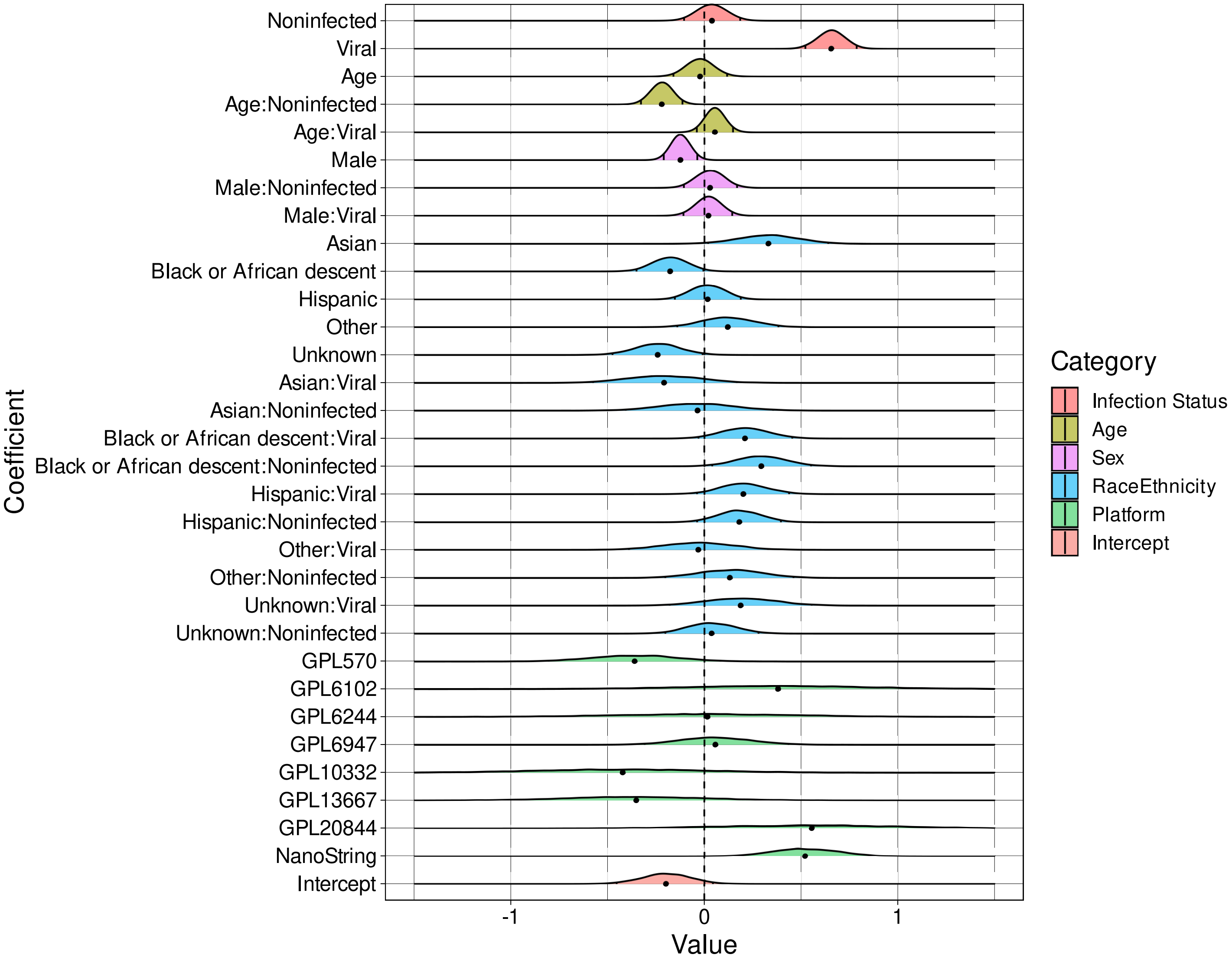


Supplementary Figure 29: **Effects inference from Bayesian multi-level model of PSMB9 expression**. Shown are density plots of samples of the posterior distributions for each of the listed effects. The posterior median is indicated by the black dot at the base of the density while tick marks within the density demarcate the 2.5th and 97.5th posterior quantiles. We do detect significant association of PSMB9 expression with infection status as well with sex, interactions between age and non-infected status and race/ethnicity (main and interaction effects). We also observe associations between marker expression and whether the sample was profiled on the NanoString platform.

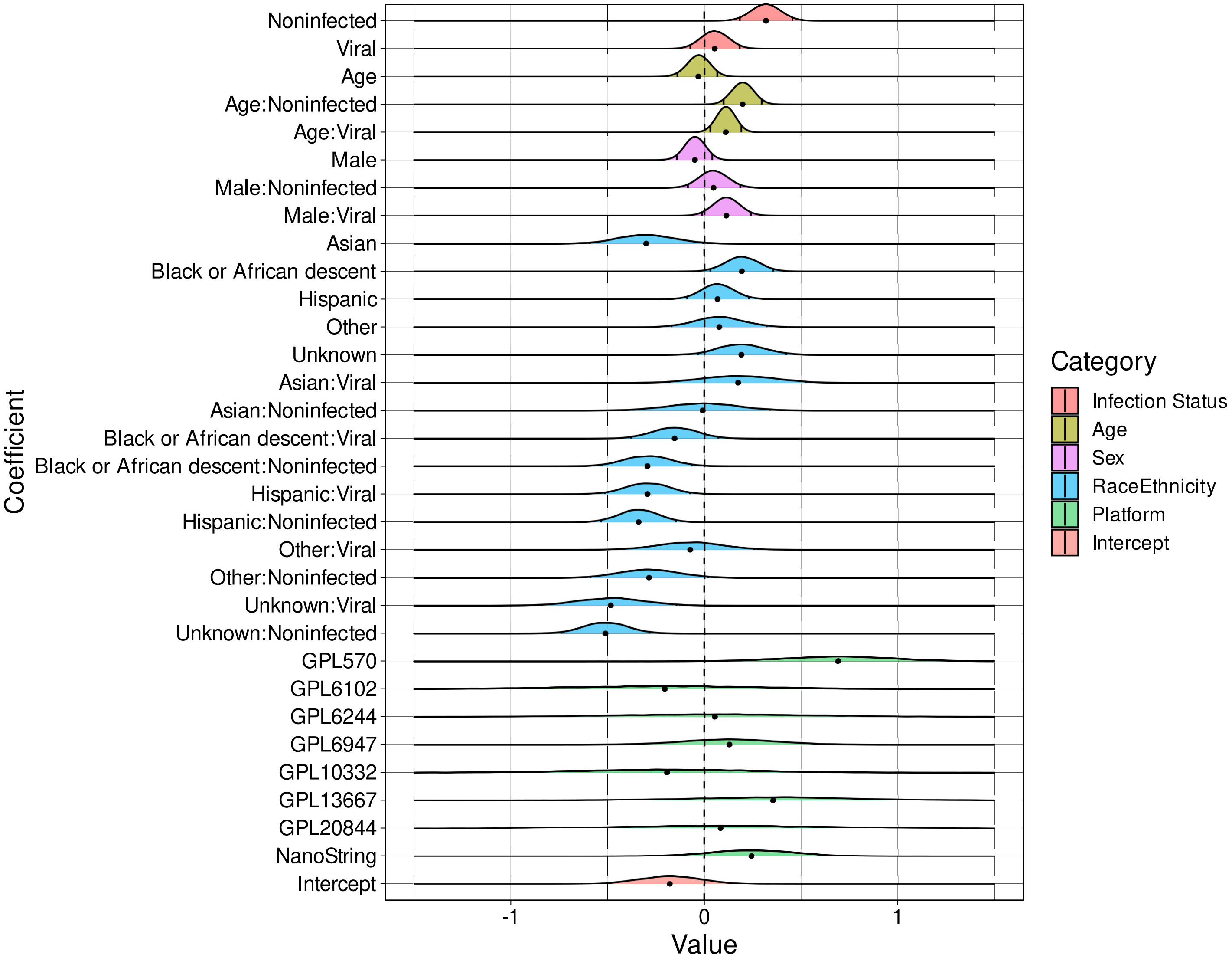


Supplementary Figure 30: **Effects inference from Bayesian multi-level model of RAPGEF1 expression**. Shown are density plots of samples of the posterior distributions for each of the listed effects. The posterior median is indicated by the black dot at the base of the density while tick marks within the density demarcate the 2.5th and 97.5th posterior quantiles. We do detect significant association (albeit mild) of RAPGEF1 expression with infection status as well as with race/ethnicity. We also observe associations between the interactions of age and race/ethnicity with infection status. We also observe associations between marker expression and whether the sample was profiled on the GPL570 microarray platform.

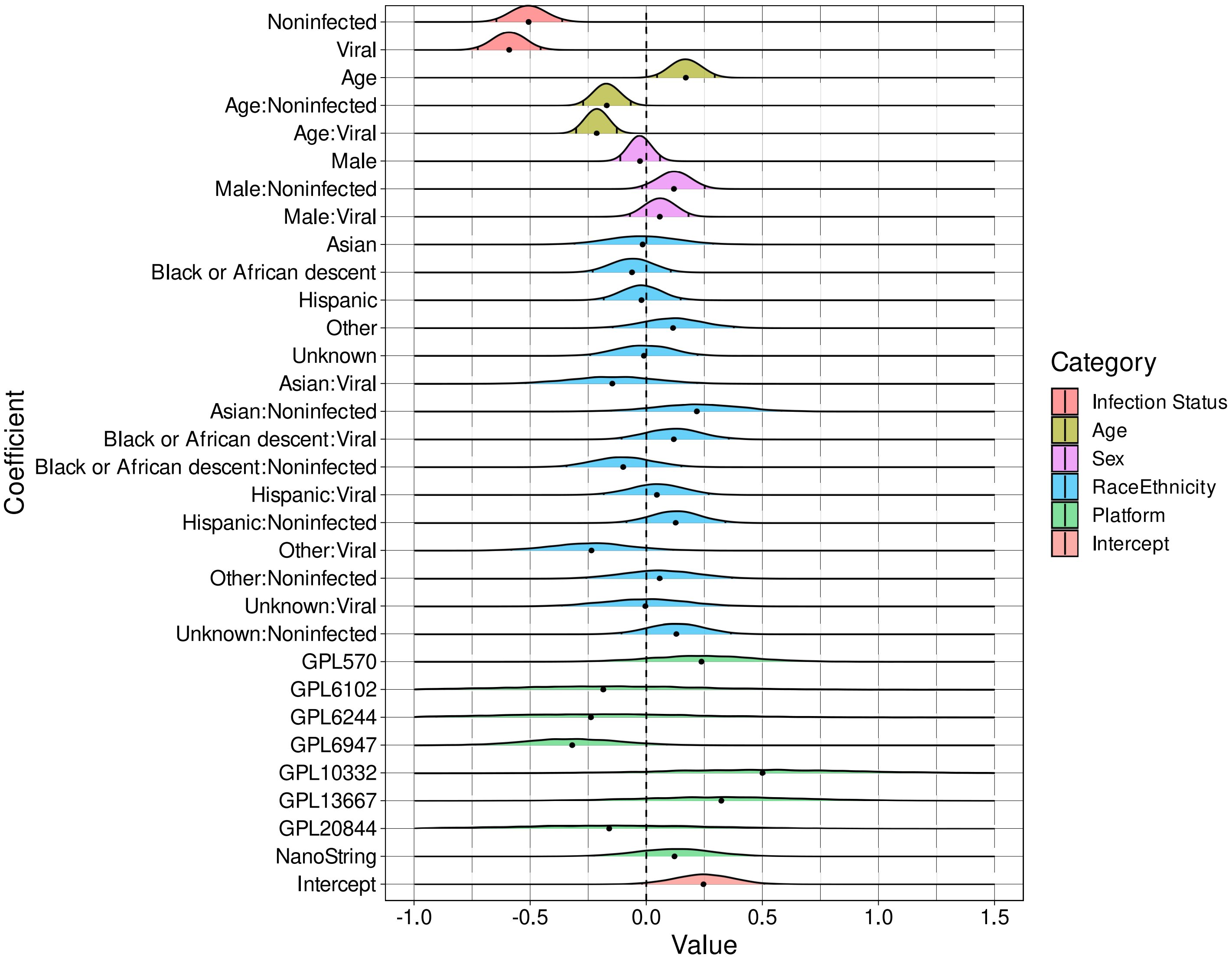


Supplementary Figure 31: **Effects inference from Bayesian multi-level model of S100A12 expression**. Shown are density plots of samples of the posterior distributions for each of the listed effects. The posterior median is indicated by the black dot at the base of the density while tick marks within the density demarcate the 2.5th and 97.5th posterior quantiles. We do detect significant association of S100A12 expression with infection status as well as with age (main and interaction effects). We do not observe associations between marker expression and technical platform.

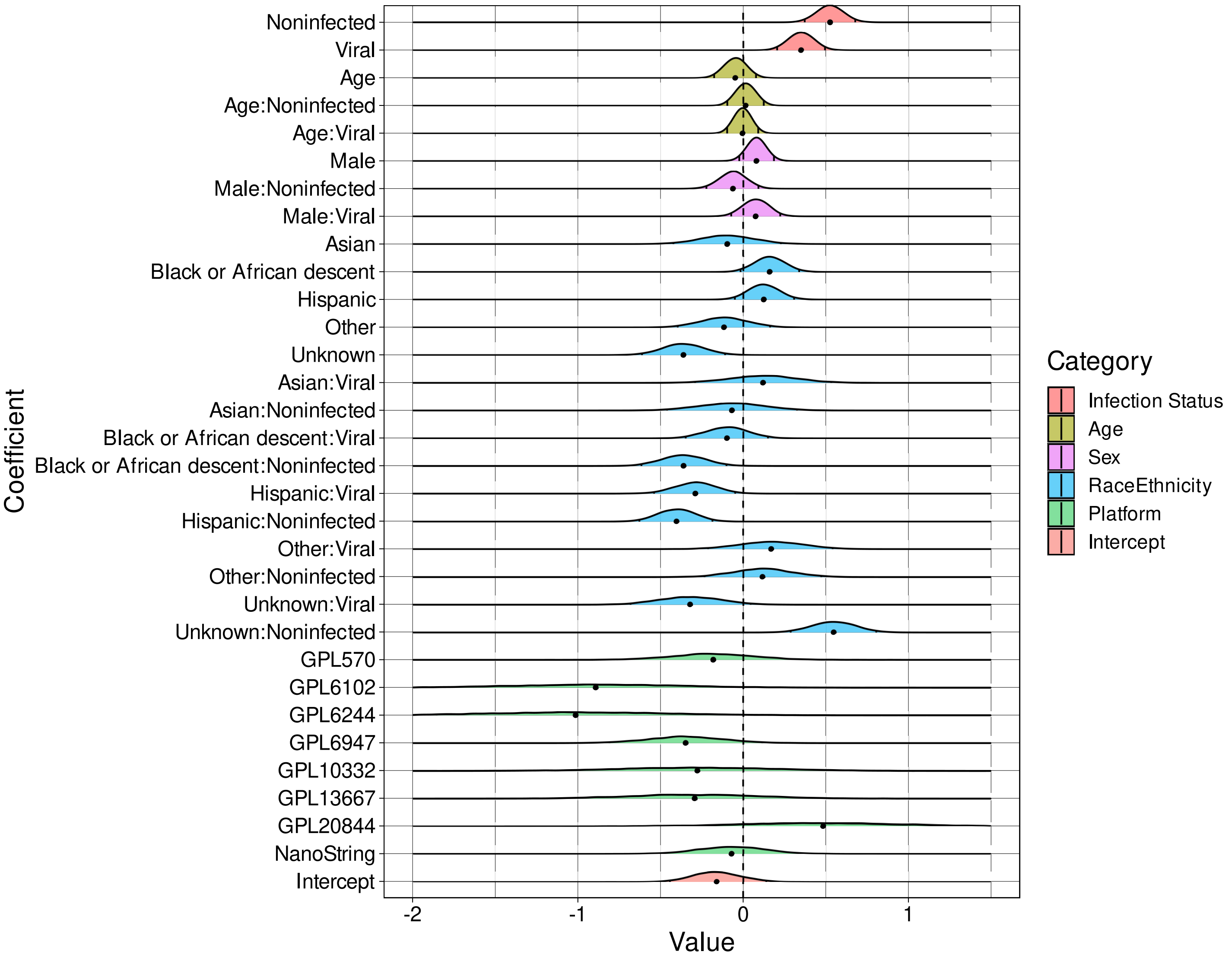


Supplementary Figure 32: **Effects inference from Bayesian multi-level model of TGFBI expression**. Shown are density plots of samples of the posterior distributions for each of the listed effects. The posterior median is indicated by the black dot at the base of the density while tick marks within the density demarcate the 2.5th and 97.5th posterior quantiles. We do detect significant association of TGFBI expression with infection status as well as with interactions between race/ethnicity and infection status. We do not observe associations between marker expression and technical platform.

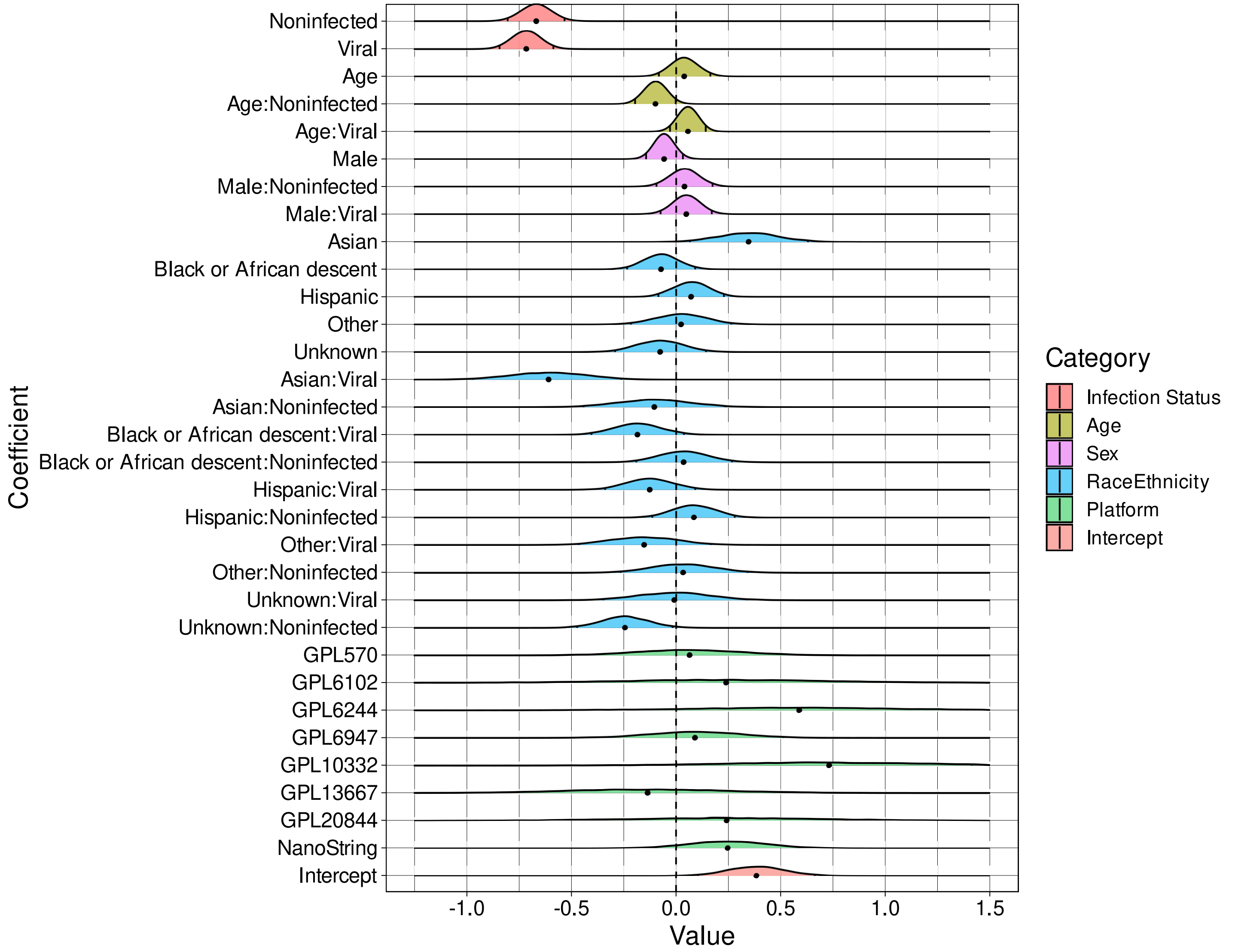


Supplementary Figure 33: **Effects inference from Bayesian multi-level model of ZDHHC19 expression**. Shown are density plots of samples of the posterior distributions for each of the listed effects. The posterior median is indicated by the black dot at the base of the density while tick marks within the density demarcate the 2.5th and 97.5th posterior quantiles. We do detect significant association of ZDHHC19 expression with infection status, main effects of race/ethnicity, and interactions between age or race/ethnicity and infection status. We do not observe associations between marker expression and technical platform.

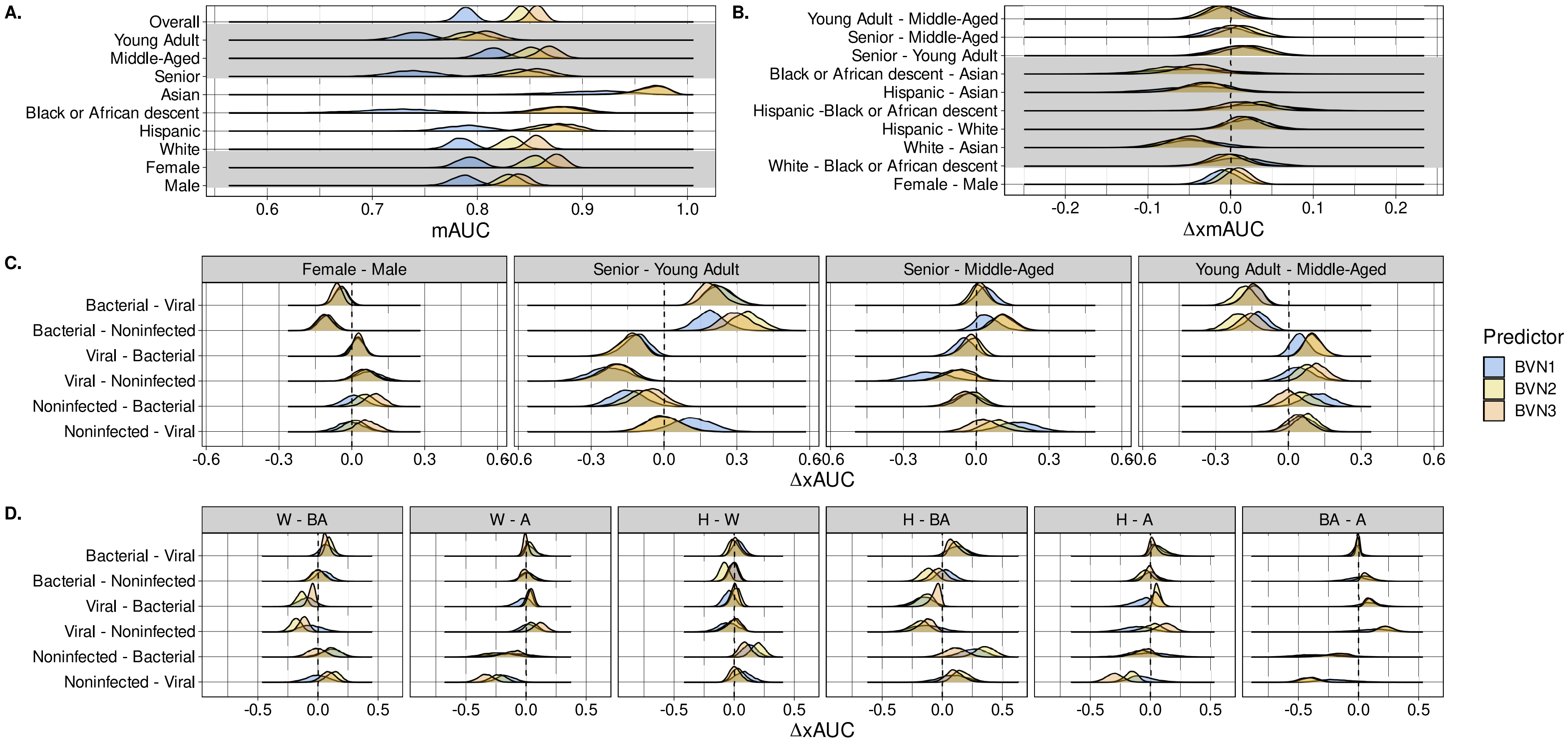


Supplementary Figure 34: **Validation performance of different generations of the BVN classifier series**. Bayesian bootstrap posterior distributions of within-group (**A.**), composite across-group (**B.**) and pairwise across-group (**C.**) performance appear for the IMX-BVN-1 [8], IMX-BVN-2 [1, 11] and IMX-BVN-3 diagnostic classifiers. A - Asian, BA - Black or African descent, H - Hispanic, W - White.

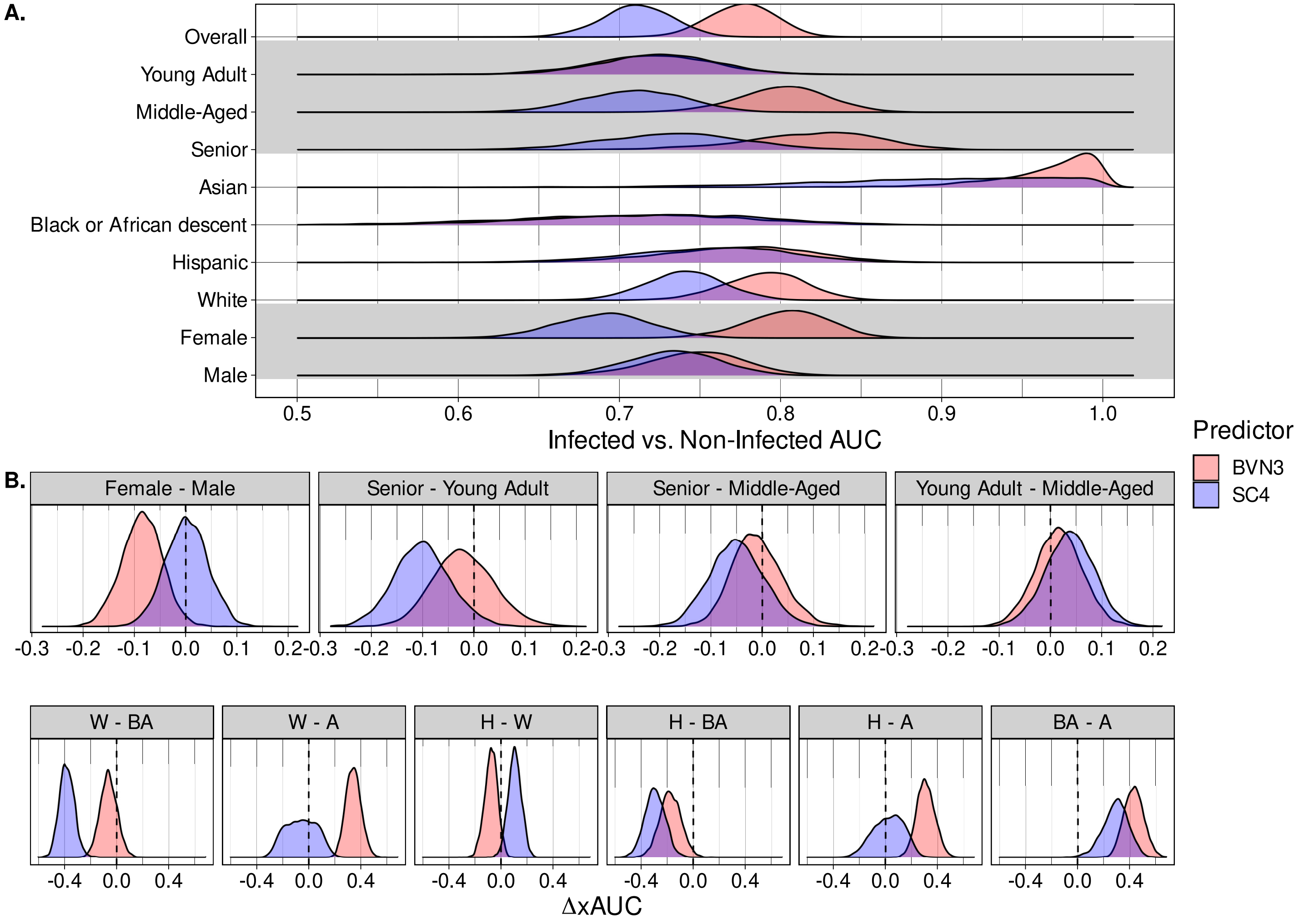


Supplementary Figure 35: **Within- and across-group performance (infected vs. non-infected) of the IMX-BVN-3 diagnostic classifier compared with SC4**. Shown are density plots of Bayesian bootstrap posterior distributions of within- (**A.**) and across- (**B.**) group measures of infected (bacterial + viral) vs. non-infected classification performance. Appearance of posterior density greater than 1.0 for IMX-BVN-3 in Asian patients is an artifact of the density estimation; nearly every posterior sample was 1.0. A - Asian, BA - Black or African descent, H - Hispanic, W - White.

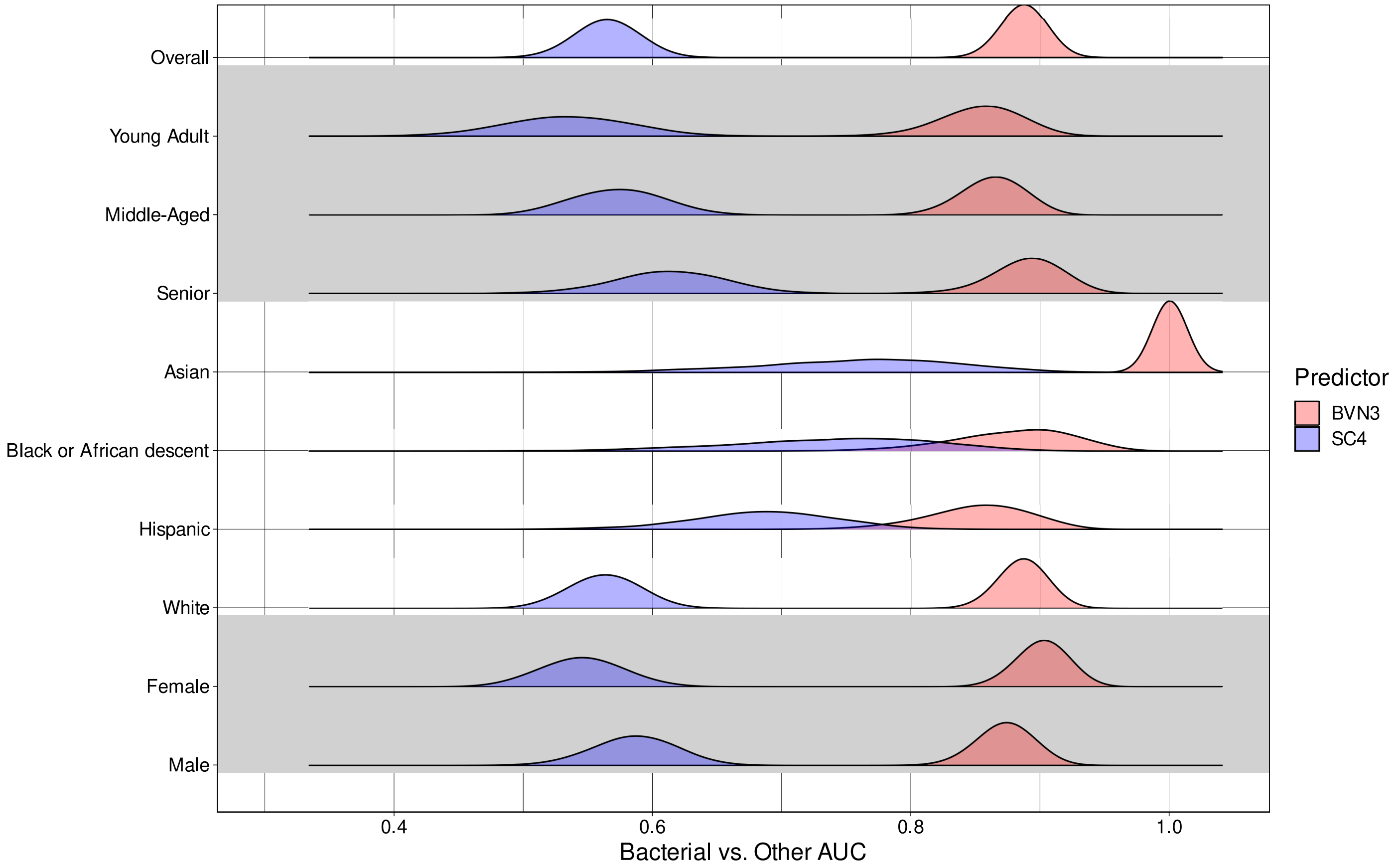


Supplementary Figure 36: **Within-group performance (bacterial infection vs. non-bacterial inflammation) of the IMX-BVN-3 diagnostic classifier compared with SC4**. Shown are density plots of Bayesian bootstrap posterior distributions of within-group measures of bacterial infection vs. non-bacterial inflammation (viral + non-infected) classification performance. Appearance of posterior density greater than 1.0 for IMX-BVN-3 in Asian patients is an artifact of the density estimation; nearly every posterior sample was 1.0. Across-group performance measures are not shown due to comparatively lower within-group performance of SC4.

Supplementary Table 1: **Infection status of patient samples in public studies.** A = adult, P = pediatric, B = bacterial infection, V = viral infection, N = non-infectious inflammation. NCBI GEO or EBI ArrayExpress IDs appear in the Study ID column.

| **Study ID** | **A/P** | **B** | **V** | **N** |
|---|---|---|---|---|
| E-MEXP-3589 | A | 4 | 5 | 14 |
| E-MTAB-1548 | A | 82 | 0 | 58 |
| E-MTAB-3162 | A | 0 | 21 | 0 |
| E-MTAB-5273 | A | 119 | 0 | 0 |
| E-MTAB-5274 | A | 108 | 0 | 0 |
| E-MTAB-5638 | A | 0 | 0 | 17 |
| GlueBuffyHCSS | A | 46 | 0 | 274 |
| GSE13015 (GPL6102) | A | 45 | 0 | 0 |
| GSE13015 (GPL6947) | A | 15 | 0 | 0 |
| GSE21802 | A | 0 | 10 | 0 |
| GSE22098 | A | 0 | 0 | 71 |
| GSE22098 | P | 52 | 0 | 70 |
| GSE25504 (GPL13667) | P | 9 | 3 | 0 |
| GSE25504 (GPL6947) | P | 20 | 1 | 0 |
| GSE27131 | A | 0 | 7 | 0 |
| GSE28750 | A | 10 | 0 | 11 |
| GSE28991 | A | 0 | 11 | 0 |
| GSE29385 | A | 0 | 80 | 0 |
| GSE30119 | P | 59 | 0 | 0 |
| GSE32707 | A | 0 | 0 | 44 |
| GSE40012 | A | 16 | 8 | 12 |
| GSE40165 | P | 0 | 123 | 0 |
| GSE40396 | P | 8 | 22 | 0 |
| GSE40586 | A | 15 | 0 | 0 |
| GSE42026 | P | 18 | 41 | 0 |
| GSE42834 | A | 14 | 0 | 68 |
| GSE47655 | A | 0 | 0 | 6 |
| GSE51808 | A | 0 | 28 | 0 |
| GSE57065 | A | 28 | 0 | 0 |
| GSE57183 | P | 0 | 0 | 11 |
| GSE60244 | A | 22 | 71 | 0 |
| GSE61821 | A | 0 | 40 | 0 |
| GSE61821 | P | 0 | 8 | 0 |
| GSE63881 | P | 0 | 0 | 171 |
| GSE64456 | P | 89 | 111 | 0 |
| GSE65682 | A | 0 | 0 | 33 |
| GSE66099 | P | 109 | 11 | 30 |
| GSE67059 | P | 0 | 80 | 0 |
| GSE68310 | A | 0 | 104 | 0 |
| GSE69528 | A | 83 | 0 | 0 |
| GSE72810 | P | 5 | 10 | 0 |
| GSE73461 | P | 52 | 94 | 162 |
| GSE77791 | A | 0 | 0 | 30 |
| GSE77087 | P | 0 | 41 | 0 |
| GSE82050 | A | 0 | 24 | 0 |
| GSE103842 | P | 0 | 62 | 0 |
| GSE111368 | A | 0 | 33 | 0 |

Supplementary Table 2: **Infection status of patient samples in NanoString-profiled studies.** B = bacterial infection, V = viral infection, N = non-infected inflammation. All studies consisted of adult patients.

| **Study ID** | **B** | **V** | **N** |
|---|---|---|---|
| INF-02-100 | 25 | 6 | 14 |
| INF-02-200 | 32 | 11 | 51 |
| INF-02-300 | 34 | 18 | 34 |
| INF-02-400 | 10 | 8 | 16 |
| INF-02-500 | 8 | 3 | 5 |
| INF-02-600 | 20 | 6 | 35 |
| INF-02-700 | 1 | 0 | 8 |
| INF-03 | 0 | 27 | 0 |
| INF-IIS-01 | 70 | 14 | 25 |
| INF-IIS-03 | 56 | 45 | 1 |
| INF-IIS-04 | 30 | 0 | 0 |
| INF-IIS-10 | 42 | 0 | 0 |
| INF-IIS-11 | 151 | 32 | 8 |
| INF-IIS-19 | 11 | 0 | 9 |
| INF-IIS-21 | 0 | 65 | 0 |

Supplementary Table 3: **Demographic characteristics of public study patients.** Country information was identified from the study's corresponding manuscript. F = female, M = male, A = Asian, BA = Black or African descent, H = Hispanic, O = Other, W = White, U = Unknown. Age data is shown as 'median (IQR)'. Studies appearing below the double horizontal line were used for IMX-BVN-3 classifier training but not included in our classifier performance analyses due to missing data.

| Study ID | Country | Sex | | | Age | Race/Ethnicity | | | | | | Platform |
|---|---|---|---|---|---|---|---|---|---|---|---|---|
| | | F | M | U | | A | BA | H | O | W | U | |
| GSE29385 | - | 23 | 48 | 9 | 25 (22,40) | 59 | 0 | 0 | 12 | 0 | 9 | GPL10558 |
| GSE40012 | Australia | 16 | 20 | 0 | 59 (46.5,67) | 0 | 0 | 0 | 0 | 0 | 36 | GPL6947 |
| GSE40586 | Estonia | 0 | 0 | 15 | 57 (53,70.5) | 0 | 0 | 0 | 0 | 0 | 15 | GPL6244 |
| GSE57065 | France | 9 | 19 | 0 | 62 (54.25,76) | 0 | 0 | 0 | 0 | 0 | 28 | GPL570 |
| GSE77791 | France | 9 | 21 | 0 | 48 (40.25,55) | 0 | 0 | 0 | 0 | 0 | 30 | GPL570 |
| GSE82050 | Germany | 9 | 13 | 2 | 64.5 (48.5,74.25) | 0 | 0 | 0 | 0 | 0 | 24 | GPL20844 |
| E-MTAB-3162 | Indonesia | 10 | 11 | 0 | 20 (17,28) | 21 | 0 | 0 | 0 | 0 | 0 | GPL570 |
| GSE65682 | Netherlands | 11 | 22 | 0 | 59 (48,67) | 3 | 4 | 0 | 0 | 24 | 2 | GPL13667 |
| GSE27131 | Norway | 1 | 6 | 0 | 38 (33,50) | 0 | 0 | 0 | 0 | 7 | 0 | GPL6244 |
| E-MTAB-1548 | Spain | 44 | 95 | 1 | 72 (71,78) | 0 | 0 | 0 | 0 | 0 | 140 | GPL10332 |
| GSE13015 | Thailand | 19 | 26 | 0 | 54 (48,61) | 45 | 0 | 0 | 0 | 0 | 0 | GPL6102 |
| GSE13015 | Thailand | 9 | 6 | 0 | 49 (43.5,59.5) | 15 | 0 | 0 | 0 | 0 | 0 | GPL6947 |
| GSE22098 | UK/S. Africa | 134 | 59 | 0 | 16 (11,26) | 9 | 46 | 73 | 2 | 63 | 0 | GPL6947 |
| E-MTAB-5273.4 | UK | 99 | 128 | 0 | 69 (54,77) | 4 | 2 | 1 | 0 | 220 | 0 | GPL10558 |
| GSE25504 | UK | 10 | 11 | 0 | 0 (0,0) | 2 | 0 | 0 | 0 | 13 | 6 | GPL6947 |
| GSE25504 | UK | 4 | 8 | 0 | 0 (0,0) | 0 | 0 | 0 | 0 | 1 | 11 | GPL13667 |
| GSE42026 | UK | 26 | 33 | 0 | 1.25 (0.375,4) | 3 | 8 | 0 | 9 | 26 | 13 | GPL6947 |
| GSE72810 | UK | 7 | 8 | 0 | 1.83 (0.88,3.29) | 0 | 3 | 0 | 2 | 8 | 2 | GPL6947 |
| GSE73461 | UK | 143 | 165 | 0 | 2.79 (0.92,8.81) | 23 | 18 | 51 | 43 | 160 | 13 | GPL10558 |
| GSE111368 | UK | 18 | 15 | 0 | 38 (29,49) | 3 | 4 | 0 | 2 | 24 | 0 | GPL10558 |
| GlueBuffyHCSS | USA | 43 | 76 | 0 | 33 (25,43) | 5 | 9 | 10 | 4 | 91 | 0 | GPL570 |
| GSE30119 | USA | 25 | 34 | 0 | 6.5 (1.92,11) | 0 | 19 | 18 | 7 | 15 | 0 | GPL6947 |
| GSE32707 | USA | 8 | 13 | 23 | 56 (45,59) | 0 | 5 | 1 | 0 | 15 | 23 | GPL10558 |
| GSE40396 | USA | 13 | 17 | 0 | 0.92 (0.33,1.60) | 0 | 15 | 0 | 2 | 13 | 0 | GPL10558 |
| GSE57183 | USA | 6 | 5 | 0 | 3.6 (3.3,7.3) | 0 | 0 | 0 | 0 | 0 | 11 | GPL10558 |
| GSE60244 | USA | 56 | 37 | 0 | 63 (50,77) | 1 | 17 | 14 | 0 | 61 | 0 | GPL10558 |
| GSE63881 | USA | 69 | 102 | 0 | 2.75 (1.42,4.25) | 25 | 6 | 38 | 41 | 36 | 25 | GPL10558 |
| GSE64456 | USA | 94 | 106 | 0 | 0.10 (0.06,0.13) | 3 | 57 | 62 | 4 | 74 | 0 | GPL10558 |
| GSE66099 | USA | 56 | 94 | 0 | 2.45 (1.00,5.88) | 4 | 26 | 2 | 8 | 99 | 11 | GPL570 |
| GSE67059 | USA | 27 | 53 | 0 | 0.83 (0.30,1.29) | 0 | 4 | 17 | 4 | 55 | 0 | GPL6947 |
| GSE68310 | USA | 54 | 50 | 0 | 20.96 (20.09,22.77) | 17 | 2 | 14 | 3 | 68 | 0 | GPL10558 |
| GSE77087 | USA | 16 | 25 | 0 | 0.45 (0.14,0.69) | 0 | 14 | 1 | 5 | 19 | 2 | GPL10558 |
| GSE103842 | USA | 23 | 39 | 0 | 0.25 (0.17,0.44) | 0 | 0 | 0 | 0 | 0 | 62 | GPL10558 |
| GSE40165 | Vietnam | 38 | 85 | 0 | 12 (10,14) | 123 | 0 | 0 | 0 | 0 | 0 | GPL10558 |
| GSE61821 | Vietnam | 24 | 24 | 0 | 40 (19.75,51) | 48 | 0 | 0 | 0 | 0 | 0 | GPL10558 |
| GSE28991 | - | 0 | 0 | 11 | - | 0 | 0 | 0 | 0 | 0 | 11 | GPL10558 |
| GSE28750 | Australia | 0 | 0 | 21 | - | 0 | 0 | 0 | 0 | 11 | 10 | GPL570 |
| GSE47655 | Australia | 0 | 0 | 6 | - | 0 | 0 | 0 | 0 | 0 | 6 | GPL6244 |
| GSE21802 | Spain/Canada | 0 | 0 | 10 | - | 0 | 0 | 0 | 0 | 0 | 10 | GPL6102 |
| E-MEXP-3589 | Spain | 5 | 16 | 2 | - | 0 | 0 | 0 | 0 | 0 | 23 | GPL10332 |
| E-MTAB-5638 | Spain | 0 | 0 | 17 | - | 0 | 0 | 0 | 0 | 0 | 17 | GPL10332 |
| GSE51808 | Thailand | 0 | 0 | 28 | - | 28 | 0 | 0 | 0 | 0 | 0 | GPL13158 |
| GSE69528 | Thailand | 0 | 0 | 83 | - | 83 | 0 | 0 | 0 | 0 | 0 | GPL10558 |
| GSE42834 | UK/France | 35 | 32 | 15 | - | 12 | 17 | 0 | 0 | 40 | 13 | GPL10558 |

Supplementary Table 4: **Demographic characteristics of NanoString study patients.** F = female, M = male, A = Asian, BA = Black or African descent, H = Hispanic, O = Other, W = White, U = Unknown. Age data is shown as 'median (IQR)'. All study patients had known biological sex.

| **Study ID** | **Country** | **Sex** | | **Age** | **Race/Ethnicity** | | | | | |
|---|---|---|---|---|---|---|---|---|---|---|
| | | **F** | **M** | | **A** | **BA** | **H** | **O** | **W** | **U** |
| INF-IIS-11 | Germany | 83 | 108 | 72 (57,81) | 0 | 0 | 0 | 0 | 191 | 0 |
| INF-02-600 | Greece | 35 | 26 | 57 (35,74) | 0 | 0 | 0 | 0 | 61 | 0 |
| INF-IIS-03 | Greece | 61 | 41 | 65.5 (38,79.75) | 0 | 0 | 0 | 0 | 102 | 0 |
| INF-IIS-04 | Greece | 12 | 18 | 72 (56.5,76.75) | 0 | 0 | 0 | 0 | 30 | 0 |
| INF-03 | India | 10 | 17 | 28 (24,37) | 27 | 0 | 0 | 0 | 0 | 0 |
| INF-IIS-21 | Spain | 25 | 40 | 81 (73,87) | 0 | 0 | 0 | 0 | 0 | 65 |
| INF-02-100 | USA | 23 | 22 | 57 (49,73) | 0 | 4 | 11 | 0 | 30 | 0 |
| INF-02-200 | USA | 53 | 41 | 39 (26.25,50) | 0 | 1 | 85 | 0 | 8 | 0 |
| INF-02-300 | USA | 39 | 47 | 43.5 (30.75,56) | 0 | 8 | 3 | 2 | 73 | 0 |
| INF-02-400 | USA | 22 | 12 | 48.5 (35.75,61) | 0 | 23 | 2 | 2 | 7 | 0 |
| INF-02-500 | USA | 9 | 7 | 48 (43.25,55.25) | 0 | 3 | 0 | 1 | 12 | 0 |
| INF-02-700 | USA | 2 | 7 | 38 (31,48) | 0 | 9 | 0 | 0 | 0 | 0 |
| INF-IIS-01 | USA | 48 | 61 | 66 (56,75) | 6 | 5 | 20 | 6 | 70 | 2 |
| INF-IIS-10 | USA | 24 | 18 | 68.5 (60.25,79.75) | 0 | 0 | 0 | 2 | 40 | 0 |
| INF-IIS-19 | USA | 6 | 14 | 59 (41.5,66.25) | 3 | 4 | 0 | 5 | 8 | 0 |

Supplementary Table 5: **Within-group composite performance of the BVN series of diagnostic classifiers**. Performance is shown in terms of the median as well as the 2.5th (LB) and 97.5th (UB) quantiles of 5000 Bayesian bootstrap samples of the posterior distribution of multi-class AUC (mAUC). Together, the LB and UB columns specify a 95% posterior confidence interval on within-group performance.

| **Subgroup Variable** | **Subgroup Value** | **Post. Median** | **95% Post. CI LB** | **95% Post. CI UB** | **Predictor** |
|---|---|---|---|---|---|
| Overall | Overall | 0.789 | 0.772 | 0.804 | IMX-BVN-1 |
| AgeBracket | Senior | 0.738 | 0.695 | 0.779 | IMX-BVN-1 |
| AgeBracket | Young Adult | 0.741 | 0.709 | 0.772 | IMX-BVN-1 |
| AgeBracket | Middle-Aged | 0.815 | 0.790 | 0.837 | IMX-BVN-1 |
| Sex | Female | 0.792 | 0.768 | 0.815 | IMX-BVN-1 |
| Sex | Male | 0.787 | 0.763 | 0.810 | IMX-BVN-1 |
| RaceEthnicity | Hispanic | 0.790 | 0.744 | 0.831 | IMX-BVN-1 |
| RaceEthnicity | White | 0.784 | 0.762 | 0.805 | IMX-BVN-1 |
| RaceEthnicity | Black or African descent | 0.725 | 0.653 | 0.789 | IMX-BVN-1 |
| RaceEthnicity | Asian | 0.909 | 0.828 | 0.962 | IMX-BVN-1 |
| Overall | Overall | 0.842 | 0.826 | 0.856 | IMX-BVN-2 |
| AgeBracket | Senior | 0.839 | 0.800 | 0.871 | IMX-BVN-2 |
| AgeBracket | Young Adult | 0.792 | 0.763 | 0.821 | IMX-BVN-2 |
| AgeBracket | Middle-Aged | 0.850 | 0.826 | 0.871 | IMX-BVN-2 |
| Sex | Female | 0.854 | 0.833 | 0.873 | IMX-BVN-2 |
| Sex | Male | 0.830 | 0.808 | 0.850 | IMX-BVN-2 |
| RaceEthnicity | Hispanic | 0.868 | 0.828 | 0.900 | IMX-BVN-2 |
| RaceEthnicity | White | 0.832 | 0.813 | 0.850 | IMX-BVN-2 |
| RaceEthnicity | Black or African descent | 0.883 | 0.838 | 0.919 | IMX-BVN-2 |
| RaceEthnicity | Asian | 0.966 | 0.918 | 0.989 | IMX-BVN-2 |
| Overall | Overall | 0.857 | 0.842 | 0.870 | IMX-BVN-3 |
| AgeBracket | Senior | 0.856 | 0.819 | 0.886 | IMX-BVN-3 |
| AgeBracket | Young Adult | 0.807 | 0.778 | 0.834 | IMX-BVN-3 |
| AgeBracket | Middle-Aged | 0.868 | 0.846 | 0.887 | IMX-BVN-3 |
| Sex | Female | 0.875 | 0.856 | 0.892 | IMX-BVN-3 |
| Sex | Male | 0.840 | 0.818 | 0.859 | IMX-BVN-3 |
| RaceEthnicity | Hispanic | 0.880 | 0.842 | 0.911 | IMX-BVN-3 |
| RaceEthnicity | White | 0.856 | 0.838 | 0.872 | IMX-BVN-3 |
| RaceEthnicity | Black or African descent | 0.876 | 0.832 | 0.913 | IMX-BVN-3 |
| RaceEthnicity | Asian | 0.967 | 0.931 | 0.988 | IMX-BVN-3 |

Supplementary Table 6: **Across-group composite performance of the BVN series of diagnostic classifiers**. Performance is shown in terms of the median as well as the 2.5th (LB) and 97.5th (UB) quantiles of 5000 Bayesian bootstrap samples of the posterior distribution of the differences in across-group multi-class AUC ($\Delta\mathsf{xmAUC}$). The 'Subgroup Value' column gives the patient subgroups used in each comparison. The order of the patient subgroups shown in this column is the same as the order of the $\mathsf{xmAUC}$ terms in the computation of $\Delta\mathsf{xmAUC}$: 'Hispanic - Black or African descent' $\rightarrow$ $\mathsf{xmAUC}_{H,BA} - \mathsf{xmAUC}_{BA,H}$. A 95% posterior confidence interval (given by the LB and UB columns) that does not include 0 would suggest a disparity in performance between the two subgroups.

| **Subgroup Variable** | **Subgroup Value** | **Post. Median** | **95% Post. CI LB** | **95% Post. CI UB** | **Predictor** |
|---|---|---|---|---|---|
| AgeBracket | Senior - Middle-Aged | -0.006 | -0.051 | 0.042 | IMX-BVN-1 |
| AgeBracket | Senior - Young Adult | 0.006 | -0.047 | 0.058 | IMX-BVN-1 |
| AgeBracket | Young Adult - Middle-Aged | -0.004 | -0.043 | 0.036 | IMX-BVN-1 |
| Sex | Female - Male | -0.010 | -0.043 | 0.022 | IMX-BVN-1 |
| RaceEthnicity | Hispanic - Asian | -0.040 | -0.126 | 0.045 | IMX-BVN-1 |
| RaceEthnicity | Hispanic - Black or African descent | 0.034 | -0.046 | 0.115 | IMX-BVN-1 |
| RaceEthnicity | Hispanic - White | 0.022 | -0.026 | 0.070 | IMX-BVN-1 |
| RaceEthnicity | White - Asian | -0.045 | -0.113 | 0.020 | IMX-BVN-1 |
| RaceEthnicity | White - Black or African descent | 0.010 | -0.054 | 0.072 | IMX-BVN-1 |
| RaceEthnicity | Black or African descent - Asian | -0.046 | -0.150 | 0.058 | IMX-BVN-1 |
| AgeBracket | Senior - Middle-Aged | 0.015 | -0.025 | 0.056 | IMX-BVN-2 |
| AgeBracket | Senior - Young Adult | 0.023 | -0.026 | 0.074 | IMX-BVN-2 |
| AgeBracket | Young Adult - Middle-Aged | -0.016 | -0.053 | 0.021 | IMX-BVN-2 |
| Sex | Female - Male | -0.000 | -0.029 | 0.029 | IMX-BVN-2 |
| RaceEthnicity | Hispanic - Asian | -0.026 | -0.081 | 0.025 | IMX-BVN-2 |
| RaceEthnicity | Hispanic - Black or African descent | 0.031 | -0.025 | 0.087 | IMX-BVN-2 |
| RaceEthnicity | Hispanic - White | 0.021 | -0.019 | 0.060 | IMX-BVN-2 |
| RaceEthnicity | White - Asian | -0.058 | -0.110 | -0.006 | IMX-BVN-2 |
| RaceEthnicity | White - Black or African descent | 0.001 | -0.041 | 0.043 | IMX-BVN-2 |
| RaceEthnicity | Black or African descent - Asian | -0.066 | -0.129 | -0.001 | IMX-BVN-2 |
| AgeBracket | Senior - Middle-Aged | 0.002 | -0.038 | 0.042 | IMX-BVN-3 |
| AgeBracket | Senior - Young Adult | 0.012 | -0.036 | 0.062 | IMX-BVN-3 |
| AgeBracket | Young Adult - Middle-Aged | -0.010 | -0.045 | 0.026 | IMX-BVN-3 |
| Sex | Female - Male | 0.009 | -0.018 | 0.037 | IMX-BVN-3 |
| RaceEthnicity | Hispanic - Asian | -0.032 | -0.085 | 0.017 | IMX-BVN-3 |
| RaceEthnicity | Hispanic - Black or African descent | 0.012 | -0.042 | 0.065 | IMX-BVN-3 |
| RaceEthnicity | Hispanic - White | 0.013 | -0.024 | 0.051 | IMX-BVN-3 |
| RaceEthnicity | White - Asian | -0.050 | -0.093 | -0.010 | IMX-BVN-3 |
| RaceEthnicity | White - Black or African descent | -0.006 | -0.047 | 0.033 | IMX-BVN-3 |
| RaceEthnicity | Black or African descent - Asian | -0.039 | -0.093 | 0.009 | IMX-BVN-3 |

Supplementary Table 7: **Across-group pairwise performance of the IMX-BVN-1 diagnostic classifier [8]**. Performance is shown in terms of the median as well as the 2.5th (LB) and 97.5th (UB) quantiles of 5000 Bayesian bootstrap samples of the posterior distribution of the differences in across-group class AUC ($\Delta$xAUC) for the 6 different pairwise comparisons (indicated in the 'Class Pair' column) in the BVN classification task. The 'Subgroup Value' column gives the patient subgroups used in each comparison. The order of the patient subgroups shown in this column combined with the order of the classes in the class pair column gives the order of the xAUC terms in the computation of $\Delta$xAUC for that class pair: 'Hispanic - Black or African descent', 'B-V' $\rightarrow \mathsf{xAUC}^{(B,V)}_{H,BA} - \mathsf{xAUC}^{(B,V)}_{BA,H}$. For the given example, the first xAUC term compares bacterial score distributions of Hispanic patients with bacterial infection to bacterial score distributions of Black or African descent patients with viral infection. The second term compares bacterial score distributions of Black or African descent patients with bacterial infection to bacterial score distributions of Hispanic viral patients. A 95% posterior confidence interval (given by the LB and UB columns) that does not include 0 would suggest a disparity in performance between the two subgroups for the corresponding pairwise class comparison.

| **Subgroup Variable** | **Subgroup Value** | **Class Pair** | **Post. Median** | **95% Post. CI LB** | **95% Post. CI UB** |
|---|---|---|---|---|---|
| AgeBracket | Senior - Middle-Aged | B-V | 0.039 | -0.035 | 0.118 |
| AgeBracket | Senior - Middle-Aged | B-N | 0.038 | -0.045 | 0.139 |
| AgeBracket | Senior - Middle-Aged | V-B | -0.053 | -0.139 | 0.021 |
| AgeBracket | Senior - Middle-Aged | V-N | -0.196 | -0.351 | -0.041 |
| AgeBracket | Senior - Middle-Aged | N-B | -0.034 | -0.150 | 0.075 |
| AgeBracket | Senior - Middle-Aged | N-V | 0.170 | 0.022 | 0.322 |
| AgeBracket | Senior - Young Adult | B-V | 0.226 | 0.126 | 0.343 |
| AgeBracket | Senior - Young Adult | B-N | 0.187 | 0.077 | 0.306 |
| AgeBracket | Senior - Young Adult | V-B | -0.111 | -0.222 | -0.021 |
| AgeBracket | Senior - Young Adult | V-N | -0.231 | -0.385 | -0.070 |
| AgeBracket | Senior - Young Adult | N-B | -0.153 | -0.282 | -0.022 |
| AgeBracket | Senior - Young Adult | N-V | 0.115 | -0.044 | 0.270 |
| AgeBracket | Young Adult - Middle-Aged | B-V | -0.145 | -0.232 | -0.071 |
| AgeBracket | Young Adult - Middle-Aged | B-N | -0.127 | -0.217 | -0.043 |
| AgeBracket | Young Adult - Middle-Aged | V-B | 0.044 | -0.022 | 0.121 |
| AgeBracket | Young Adult - Middle-Aged | V-N | 0.036 | -0.078 | 0.153 |
| AgeBracket | Young Adult - Middle-Aged | N-B | 0.117 | 0.004 | 0.236 |
| AgeBracket | Young Adult - Middle-Aged | N-V | 0.050 | -0.056 | 0.156 |
| Sex | Female - Male | B-V | -0.042 | -0.096 | 0.012 |
| Sex | Female - Male | B-N | -0.114 | -0.178 | -0.051 |
| Sex | Female - Male | V-B | 0.023 | -0.035 | 0.078 |
| Sex | Female - Male | V-N | 0.071 | -0.037 | 0.173 |
| Sex | Female - Male | N-B | 0.008 | -0.083 | 0.094 |
| Sex | Female - Male | N-V | -0.007 | -0.107 | 0.095 |
| RaceEthnicity | Hispanic - Asian | B-V | 0.046 | -0.028 | 0.234 |
| RaceEthnicity | Hispanic - Asian | B-N | -0.004 | -0.154 | 0.156 |
| RaceEthnicity | Hispanic - Asian | V-B | -0.072 | -0.278 | 0.030 |
| RaceEthnicity | Hispanic - Asian | V-N | -0.118 | -0.362 | 0.083 |
| RaceEthnicity | Hispanic - Asian | N-B | -0.018 | -0.356 | 0.294 |
| RaceEthnicity | Hispanic - Asian | N-V | -0.082 | -0.248 | 0.166 |
| RaceEthnicity | Hispanic - Black or African descent | B-V | 0.128 | -0.022 | 0.318 |
| RaceEthnicity | Hispanic - Black or African descent | B-N | 0.026 | -0.105 | 0.181 |
| RaceEthnicity | Hispanic - Black or African descent | V-B | -0.161 | -0.355 | -0.000 |
| RaceEthnicity | Hispanic - Black or African descent | V-N | -0.137 | -0.366 | 0.117 |
| RaceEthnicity | Hispanic - Black or African descent | N-B | 0.255 | 0.052 | 0.432 |
| RaceEthnicity | Hispanic - Black or African descent | N-V | 0.086 | -0.150 | 0.311 |
| RaceEthnicity | Hispanic - White | B-V | 0.033 | -0.055 | 0.135 |
| RaceEthnicity | Hispanic - White | B-N | -0.003 | -0.083 | 0.068 |
| RaceEthnicity | Hispanic - White | V-B | -0.046 | -0.138 | 0.038 |
| RaceEthnicity | Hispanic - White | V-N | -0.059 | -0.222 | 0.094 |
| RaceEthnicity | Hispanic - White | N-B | 0.125 | 0.016 | 0.244 |
| RaceEthnicity | Hispanic - White | N-V | 0.076 | -0.056 | 0.225 |
| RaceEthnicity | White - Asian | B-V | 0.028 | -0.037 | 0.152 |

**Supplementary Table 7 – continued from previous page**

| Subgroup Variable | Subgroup Value | Class Pair | Post. Median | 95% Post. CI LB | 95% Post. CI UB |
|---|---|---|---|---|---|
| RaceEthnicity | White - Asian | B-N | 0.016 | -0.102 | 0.149 |
| RaceEthnicity | White - Asian | V-B | -0.027 | -0.166 | 0.057 |
| RaceEthnicity | White - Asian | V-N | 0.015 | -0.103 | 0.137 |
| RaceEthnicity | White - Asian | N-B | -0.134 | -0.449 | 0.155 |
| RaceEthnicity | White - Asian | N-V | -0.174 | -0.309 | -0.026 |
| RaceEthnicity | White - Black or African descent | B-V | 0.068 | -0.036 | 0.184 |
| RaceEthnicity | White - Black or African descent | B-N | 0.049 | -0.061 | 0.176 |
| RaceEthnicity | White - Black or African descent | V-B | -0.096 | -0.249 | 0.042 |
| RaceEthnicity | White - Black or African descent | V-N | -0.061 | -0.235 | 0.144 |
| RaceEthnicity | White - Black or African descent | N-B | 0.109 | -0.062 | 0.256 |
| RaceEthnicity | White - Black or African descent | N-V | -0.010 | -0.194 | 0.156 |
| RaceEthnicity | Black or African descent - Asian | B-V | -0.021 | -0.104 | 0.080 |
| RaceEthnicity | Black or African descent - Asian | B-N | -0.015 | -0.218 | 0.168 |
| RaceEthnicity | Black or African descent - Asian | V-B | 0.084 | -0.111 | 0.282 |
| RaceEthnicity | Black or African descent - Asian | V-N | 0.119 | -0.197 | 0.328 |
| RaceEthnicity | Black or African descent - Asian | N-B | -0.251 | -0.624 | 0.126 |
| RaceEthnicity | Black or African descent - Asian | N-V | -0.193 | -0.409 | 0.147 |

Supplementary Table 8: **Across-group pairwise performance of the IMX-BVN-2 diagnostic classifier [1, 11].** Performance is shown in terms of the median as well as the 2.5th (LB) and 97.5th (UB) quantiles of 5000 Bayesian bootstrap samples of the posterior distribution of the differences in across-group class AUC (ΔxAUC) for the 6 different pairwise comparisons (indicated in the 'Class Pair' column) in the BVN classification task. The 'Subgroup Value' column gives the patient subgroups used in each comparison. The order of the patient subgroups shown in this column combined with the order of the classes in the class pair column gives the order of the xAUC terms in the computation of ΔxAUC for that class pair (as in Supplementary Table 7). A 95% posterior confidence interval (given by the LB and UB columns) that does not include 0 would suggest a disparity in performance between the two subgroups for the corresponding pairwise class comparison.

| Subgroup Variable | Subgroup Value | Class Pair | Post. Median | 95% Post. CI LB | 95% Post. CI UB |
|---|---|---|---|---|---|
| AgeBracket | Senior - Middle-Aged | B-V | 0.006 | -0.059 | 0.078 |
| AgeBracket | Senior - Middle-Aged | B-N | 0.106 | 0.020 | 0.207 |
| AgeBracket | Senior - Middle-Aged | V-B | -0.016 | -0.105 | 0.054 |
| AgeBracket | Senior - Middle-Aged | V-N | -0.082 | -0.218 | 0.042 |
| AgeBracket | Senior - Middle-Aged | N-B | -0.018 | -0.120 | 0.075 |
| AgeBracket | Senior - Middle-Aged | N-V | 0.093 | -0.018 | 0.232 |
| AgeBracket | Senior - Young Adult | B-V | 0.221 | 0.129 | 0.336 |
| AgeBracket | Senior - Young Adult | B-N | 0.341 | 0.227 | 0.460 |
| AgeBracket | Senior - Young Adult | V-B | -0.134 | -0.254 | -0.047 |
| AgeBracket | Senior - Young Adult | V-N | -0.190 | -0.330 | -0.069 |
| AgeBracket | Senior - Young Adult | N-B | -0.094 | -0.228 | 0.032 |
| AgeBracket | Senior - Young Adult | N-V | -0.004 | -0.138 | 0.145 |
| AgeBracket | Young Adult - Middle-Aged | B-V | -0.184 | -0.275 | -0.111 |
| AgeBracket | Young Adult - Middle-Aged | B-N | -0.211 | -0.309 | -0.123 |
| AgeBracket | Young Adult - Middle-Aged | V-B | 0.097 | 0.031 | 0.179 |
| AgeBracket | Young Adult - Middle-Aged | V-N | 0.074 | -0.023 | 0.177 |
| AgeBracket | Young Adult - Middle-Aged | N-B | 0.051 | -0.056 | 0.162 |
| AgeBracket | Young Adult - Middle-Aged | N-V | 0.075 | -0.014 | 0.164 |
| Sex | Female - Male | B-V | -0.045 | -0.098 | 0.004 |
| Sex | Female - Male | B-N | -0.102 | -0.165 | -0.039 |
| Sex | Female - Male | V-B | 0.025 | -0.026 | 0.078 |
| Sex | Female - Male | V-N | 0.059 | -0.028 | 0.146 |
| Sex | Female - Male | N-B | 0.052 | -0.032 | 0.134 |
| Sex | Female - Male | N-V | 0.013 | -0.072 | 0.103 |

**Supplementary Table 8 – continued from previous page**

| Subgroup Variable | Subgroup Value | Class Pair | Post. Median | 95% Post. CI LB | 95% Post. CI UB |
|---|---|---|---|---|---|
| RaceEthnicity | Hispanic - Asian | B-V | 0.070 | -0.007 | 0.235 |
| RaceEthnicity | Hispanic - Asian | B-N | -0.050 | -0.167 | 0.062 |
| RaceEthnicity | Hispanic - Asian | V-B | 0.051 | 0.019 | 0.120 |
| RaceEthnicity | Hispanic - Asian | V-N | 0.017 | -0.179 | 0.144 |
| RaceEthnicity | Hispanic - Asian | N-B | -0.085 | -0.285 | 0.087 |
| RaceEthnicity | Hispanic - Asian | N-V | -0.159 | -0.276 | -0.053 |
| RaceEthnicity | Hispanic - Black or African descent | B-V | 0.120 | 0.037 | 0.269 |
| RaceEthnicity | Hispanic - Black or African descent | B-N | -0.118 | -0.241 | 0.004 |
| RaceEthnicity | Hispanic - Black or African descent | V-B | -0.141 | -0.302 | -0.046 |
| RaceEthnicity | Hispanic - Black or African descent | V-N | -0.187 | -0.360 | -0.063 |
| RaceEthnicity | Hispanic - Black or African descent | N-B | 0.354 | 0.204 | 0.503 |
| RaceEthnicity | Hispanic - Black or African descent | N-V | 0.154 | 0.036 | 0.312 |
| RaceEthnicity | Hispanic - White | B-V | -0.012 | -0.090 | 0.084 |
| RaceEthnicity | Hispanic - White | B-N | -0.086 | -0.165 | -0.016 |
| RaceEthnicity | Hispanic - White | V-B | 0.013 | -0.067 | 0.081 |
| RaceEthnicity | Hispanic - White | V-N | -0.015 | -0.158 | 0.095 |
| RaceEthnicity | Hispanic - White | N-B | 0.196 | 0.093 | 0.298 |
| RaceEthnicity | Hispanic - White | N-V | 0.030 | -0.064 | 0.145 |
| RaceEthnicity | White - Asian | B-V | 0.035 | -0.017 | 0.121 |
| RaceEthnicity | White - Asian | B-N | 0.009 | -0.085 | 0.126 |
| RaceEthnicity | White - Asian | V-B | 0.026 | -0.032 | 0.078 |
| RaceEthnicity | White - Asian | V-N | 0.043 | -0.059 | 0.144 |
| RaceEthnicity | White - Asian | N-B | -0.236 | -0.459 | -0.024 |
| RaceEthnicity | White - Asian | N-V | -0.231 | -0.357 | -0.114 |
| RaceEthnicity | White - Black or African descent | B-V | 0.093 | 0.038 | 0.168 |
| RaceEthnicity | White - Black or African descent | B-N | -0.003 | -0.108 | 0.109 |
| RaceEthnicity | White - Black or African descent | V-B | -0.139 | -0.227 | -0.070 |
| RaceEthnicity | White - Black or African descent | V-N | -0.184 | -0.279 | -0.094 |
| RaceEthnicity | White - Black or African descent | N-B | 0.111 | -0.027 | 0.242 |
| RaceEthnicity | White - Black or African descent | N-V | 0.131 | -0.001 | 0.234 |
| RaceEthnicity | Black or African descent - Asian | B-V | -0.004 | -0.030 | -0.000 |
| RaceEthnicity | Black or African descent - Asian | B-N | 0.056 | -0.092 | 0.215 |
| RaceEthnicity | Black or African descent - Asian | V-B | 0.098 | 0.036 | 0.205 |
| RaceEthnicity | Black or African descent - Asian | V-N | 0.223 | 0.124 | 0.353 |
| RaceEthnicity | Black or African descent - Asian | N-B | -0.373 | -0.667 | -0.093 |
| RaceEthnicity | Black or African descent - Asian | N-V | -0.400 | -0.546 | -0.266 |

Supplementary Table 9: **Across-group pairwise performance of the IMX-BVN-3 diagnostic classifier**. Performance is shown in terms of the median as well as the 2.5th (LB) and 97.5th (UB) quantiles of 5000 Bayesian bootstrap samples of the posterior distribution of the differences in across-group class AUC ($\Delta$xAUC) for the 6 different pairwise comparisons (indicated in the 'Class Pair' column) in the BVN classification task. The 'Subgroup Value' column gives the patient subgroups used in each comparison. The order of the patient subgroups shown in this column combined with the order of the classes in the class pair column gives the order of the xAUC terms in the computation of $\Delta$xAUC for that class pair (as in Supplementary Table 7). A 95% posterior confidence interval (given by the LB and UB columns) that does not include 0 would suggest a disparity in performance between the two subgroups for the corresponding pairwise class comparison.

| Subgroup Variable | Subgroup Value | Class Pair | Post. Median | 95% Post. CI LB | 95% Post. CI UB |
|---|---|---|---|---|---|
| AgeBracket | Senior - Middle-Aged | B-V | 0.016 | -0.045 | 0.084 |
| AgeBracket | Senior - Middle-Aged | B-N | 0.113 | 0.029 | 0.208 |
| AgeBracket | Senior - Middle-Aged | V-B | -0.027 | -0.105 | 0.030 |
| AgeBracket | Senior - Middle-Aged | V-N | -0.066 | -0.200 | 0.051 |
| AgeBracket | Senior - Middle-Aged | N-B | -0.048 | -0.148 | 0.044 |

**Supplementary Table 9 – continued from previous page**

| Subgroup Variable | Subgroup Value | Class Pair | Post. Median | 95% Post. CI LB | 95% Post. CI UB |
|---|---|---|---|---|---|
| AgeBracket | Senior - Middle-Aged | N-V | 0.028 | -0.087 | 0.156 |
| AgeBracket | Senior - Young Adult | B-V | 0.186 | 0.106 | 0.291 |
| AgeBracket | Senior - Young Adult | B-N | 0.291 | 0.178 | 0.414 |
| AgeBracket | Senior - Young Adult | V-B | -0.136 | -0.250 | -0.058 |
| AgeBracket | Senior - Young Adult | V-N | -0.202 | -0.341 | -0.090 |
| AgeBracket | Senior - Young Adult | N-B | -0.055 | -0.187 | 0.062 |
| AgeBracket | Senior - Young Adult | N-V | -0.006 | -0.141 | 0.137 |
| AgeBracket | Young Adult - Middle-Aged | B-V | -0.147 | -0.225 | -0.081 |
| AgeBracket | Young Adult - Middle-Aged | B-N | -0.156 | -0.248 | -0.069 |
| AgeBracket | Young Adult - Middle-Aged | V-B | 0.099 | 0.037 | 0.175 |
| AgeBracket | Young Adult - Middle-Aged | V-N | 0.115 | 0.028 | 0.209 |
| AgeBracket | Young Adult - Middle-Aged | N-B | -0.007 | -0.109 | 0.097 |
| AgeBracket | Young Adult - Middle-Aged | N-V | 0.037 | -0.055 | 0.127 |
| Sex | Female - Male | B-V | -0.061 | -0.105 | -0.019 |
| Sex | Female - Male | B-N | -0.112 | -0.174 | -0.051 |
| Sex | Female - Male | V-B | 0.027 | -0.013 | 0.069 |
| Sex | Female - Male | V-N | 0.047 | -0.038 | 0.126 |
| Sex | Female - Male | N-B | 0.096 | 0.017 | 0.170 |
| Sex | Female - Male | N-V | 0.060 | -0.025 | 0.148 |
| RaceEthnicity | Hispanic - Asian | B-V | 0.014 | -0.041 | 0.132 |
| RaceEthnicity | Hispanic - Asian | B-N | -0.009 | -0.092 | 0.092 |
| RaceEthnicity | Hispanic - Asian | V-B | 0.037 | -0.075 | 0.123 |
| RaceEthnicity | Hispanic - Asian | V-N | 0.121 | -0.047 | 0.255 |
| RaceEthnicity | Hispanic - Asian | N-B | -0.053 | -0.236 | 0.074 |
| RaceEthnicity | Hispanic - Asian | N-V | -0.304 | -0.443 | -0.172 |
| RaceEthnicity | Hispanic - Black or African descent | B-V | 0.083 | 0.019 | 0.234 |
| RaceEthnicity | Hispanic - Black or African descent | B-N | -0.033 | -0.161 | 0.099 |
| RaceEthnicity | Hispanic - Black or African descent | V-B | -0.050 | -0.158 | -0.004 |
| RaceEthnicity | Hispanic - Black or African descent | V-N | -0.128 | -0.283 | -0.024 |
| RaceEthnicity | Hispanic - Black or African descent | N-B | 0.107 | -0.074 | 0.276 |
| RaceEthnicity | Hispanic - Black or African descent | N-V | 0.094 | -0.035 | 0.240 |
| RaceEthnicity | Hispanic - White | B-V | 0.011 | -0.057 | 0.119 |
| RaceEthnicity | Hispanic - White | B-N | -0.014 | -0.097 | 0.055 |
| RaceEthnicity | Hispanic - White | V-B | -0.002 | -0.087 | 0.054 |
| RaceEthnicity | Hispanic - White | V-N | 0.002 | -0.134 | 0.095 |
| RaceEthnicity | Hispanic - White | N-B | 0.084 | -0.007 | 0.184 |
| RaceEthnicity | Hispanic - White | N-V | 0.003 | -0.079 | 0.112 |
| RaceEthnicity | White - Asian | B-V | -0.006 | -0.045 | 0.043 |
| RaceEthnicity | White - Asian | B-N | -0.006 | -0.068 | 0.095 |
| RaceEthnicity | White - Asian | V-B | 0.040 | -0.014 | 0.089 |
| RaceEthnicity | White - Asian | V-N | 0.117 | 0.015 | 0.221 |
| RaceEthnicity | White - Asian | N-B | -0.108 | -0.300 | 0.025 |
| RaceEthnicity | White - Asian | N-V | -0.338 | -0.463 | -0.222 |
| RaceEthnicity | White - Black or African descent | B-V | 0.061 | 0.017 | 0.123 |
| RaceEthnicity | White - Black or African descent | B-N | 0.002 | -0.103 | 0.114 |
| RaceEthnicity | White - Black or African descent | V-B | -0.050 | -0.110 | -0.010 |
| RaceEthnicity | White - Black or African descent | V-N | -0.118 | -0.203 | -0.039 |
| RaceEthnicity | White - Black or African descent | N-B | -0.013 | -0.160 | 0.122 |
| RaceEthnicity | White - Black or African descent | N-V | 0.081 | -0.043 | 0.191 |
| RaceEthnicity | Black or African descent - Asian | B-V | -0.011 | -0.064 | -0.002 |
| RaceEthnicity | Black or African descent - Asian | B-N | 0.063 | 0.011 | 0.186 |
| RaceEthnicity | Black or African descent - Asian | V-B | 0.084 | 0.029 | 0.182 |
| RaceEthnicity | Black or African descent - Asian | V-N | 0.226 | 0.125 | 0.361 |
| RaceEthnicity | Black or African descent - Asian | N-B | -0.179 | -0.428 | -0.018 |
| RaceEthnicity | Black or African descent - Asian | N-V | -0.422 | -0.567 | -0.288 |

Supplementary Table 10: **Within-group performance of the IMX-BVN-3 diagnostic classifier compared with procalcitonin (PCT)**. Performance is shown in terms of the median as well as the 2.5th (LB) and 97.5th (UB) quantiles of 5000 Bayesian bootstrap samples of the posterior distribution of bacterial-vs.-non-bacterial AUC (BAO AUC). Together, the LB and UB columns specify a 95% posterior confidence interval on within-group performance.

| Subgroup Variable | Subgroup Value | Post. Median | 95% Post. CI LB | 95% Post. CI UB | Predictor |
|---|---|---|---|---|---|
| Overall | Overall | 0.887 | 0.863 | 0.909 | IMX-BVN-3 |
| Overall | Overall | 0.834 | 0.805 | 0.862 | PCT |
| AgeBracket | Senior | 0.866 | 0.805 | 0.912 | IMX-BVN-3 |
| AgeBracket | Young Adult | 0.867 | 0.805 | 0.912 | IMX-BVN-3 |
| AgeBracket | Middle-Aged | 0.870 | 0.828 | 0.906 | IMX-BVN-3 |
| AgeBracket | Senior | 0.810 | 0.733 | 0.869 | PCT |
| AgeBracket | Young Adult | 0.784 | 0.703 | 0.848 | PCT |
| AgeBracket | Middle-Aged | 0.821 | 0.770 | 0.865 | PCT |
| Sex | Female | 0.906 | 0.874 | 0.932 | IMX-BVN-3 |
| Sex | Male | 0.871 | 0.833 | 0.904 | IMX-BVN-3 |
| Sex | Female | 0.848 | 0.806 | 0.884 | PCT |
| Sex | Male | 0.820 | 0.777 | 0.860 | PCT |
| RaceEthnicity | Hispanic | 0.874 | 0.801 | 0.927 | IMX-BVN-3 |
| RaceEthnicity | White | 0.883 | 0.852 | 0.909 | IMX-BVN-3 |
| RaceEthnicity | Black or African descent | 0.859 | 0.742 | 0.937 | IMX-BVN-3 |
| RaceEthnicity | Asian | 0.994 | 0.944 | 1.000 | IMX-BVN-3 |
| RaceEthnicity | Hispanic | 0.794 | 0.701 | 0.868 | PCT |
| RaceEthnicity | White | 0.832 | 0.797 | 0.865 | PCT |
| RaceEthnicity | Black or African descent | 0.734 | 0.546 | 0.868 | PCT |
| RaceEthnicity | Asian | 1.000 | 1.000 | 1.000 | PCT |

Supplementary Table 11: **Across-group performance of the IMX-BVN-3 diagnostic classifier compared with procalcitonin (PCT)**. Performance is shown in terms of the median as well as the 2.5th (LB) and 97.5th (UB) quantiles of 5000 Bayesian bootstrap samples of the posterior distribution of across-group bacterial-vs.-non-bacterial AUC. A 95% posterior confidence interval (given by the LB and UB columns) that does not include 0 would suggest a disparity in performance between the two subgroups for the comparison of patients with bacterial infection to patients without bacterial infection.

| Subgroup Variable | Subgroup Value | Post. Median | 95% Post. CI LB | 95% Post. CI UB | Predictor |
|---|---|---|---|---|---|
| AgeBracket | Senior - Middle-Aged | 0.074 | 0.008 | 0.149 | IMX-BVN-3 |
| AgeBracket | Senior - Young Adult | 0.230 | 0.141 | 0.335 | IMX-BVN-3 |
| AgeBracket | Young Adult - Middle-Aged | -0.133 | -0.210 | -0.064 | IMX-BVN-3 |
| AgeBracket | Senior - Middle-Aged | 0.041 | -0.035 | 0.125 | PCT |
| AgeBracket | Senior - Young Adult | 0.272 | 0.167 | 0.384 | PCT |
| AgeBracket | Young Adult - Middle-Aged | -0.221 | -0.315 | -0.134 | PCT |
| Sex | Female - Male | -0.077 | -0.126 | -0.033 | IMX-BVN-3 |
| Sex | Female - Male | -0.095 | -0.154 | -0.036 | PCT |
| RaceEthnicity | Hispanic - Asian | 0.027 | -0.040 | 0.155 | IMX-BVN-3 |
| RaceEthnicity | Hispanic - Black or African descent | -0.001 | -0.116 | 0.121 | IMX-BVN-3 |
| RaceEthnicity | Hispanic - White | 0.003 | -0.059 | 0.060 | IMX-BVN-3 |
| RaceEthnicity | White - Asian | 0.023 | -0.033 | 0.121 | IMX-BVN-3 |
| RaceEthnicity | White - Black or African descent | 0.007 | -0.081 | 0.099 | IMX-BVN-3 |
| RaceEthnicity | Black or African descent - Asian | 0.048 | -0.025 | 0.186 | IMX-BVN-3 |
| RaceEthnicity | Hispanic - Asian | -0.030 | -0.105 | 0.019 | PCT |
| RaceEthnicity | Hispanic - Black or African descent | -0.019 | -0.171 | 0.169 | PCT |
| RaceEthnicity | Hispanic - White | -0.038 | -0.135 | 0.046 | PCT |

**Supplementary Table 11 – continued from previous page**

| Subgroup Variable | Subgroup Value | Post. Median | 95% Post. CI LB | 95% Post. CI UB | Predictor |
|---|---|---|---|---|---|
| RaceEthnicity | White - Asian | -0.032 | -0.105 | -0.000 | PCT |
| RaceEthnicity | White - Black or African descent | 0.019 | -0.110 | 0.189 | PCT |
| RaceEthnicity | Black or African descent - Asian | -0.049 | -0.140 | -0.011 | PCT |

Supplementary Table 12: **Within-group performance of the IMX-BVN-3 diagnostic classifier compared with the SeptiCyte 4-marker panel (SC4).** Performance is shown in terms of the median as well as the 2.5th (LB) and 97.5th (UB) quantiles of 5000 Bayesian bootstrap samples of the posterior distribution of infected-vs.-non-infected AUC. Together, the LB and UB columns specify a 95% posterior confidence interval on within-group performance.

| Subgroup Variable | Subgroup Value | Post. Median | 95% Post. CI LB | 95% Post. CI UB | Predictor |
|---|---|---|---|---|---|
| Overall | Overall | 0.776 | 0.736 | 0.813 | IMX-BVN-3 |
| Overall | Overall | 0.711 | 0.667 | 0.749 | SC4 |
| AgeBracket | Senior | 0.822 | 0.720 | 0.890 | IMX-BVN-3 |
| AgeBracket | Young Adult | 0.722 | 0.641 | 0.790 | IMX-BVN-3 |
| AgeBracket | Middle-Aged | 0.802 | 0.745 | 0.850 | IMX-BVN-3 |
| AgeBracket | Senior | 0.735 | 0.639 | 0.815 | SC4 |
| AgeBracket | Young Adult | 0.726 | 0.649 | 0.790 | SC4 |
| AgeBracket | Middle-Aged | 0.710 | 0.644 | 0.768 | SC4 |
| Sex | Female | 0.805 | 0.752 | 0.850 | IMX-BVN-3 |
| Sex | Male | 0.747 | 0.686 | 0.802 | IMX-BVN-3 |
| Sex | Female | 0.691 | 0.632 | 0.745 | SC4 |
| Sex | Male | 0.734 | 0.674 | 0.786 | SC4 |
| RaceEthnicity | Hispanic | 0.774 | 0.677 | 0.853 | IMX-BVN-3 |
| RaceEthnicity | White | 0.791 | 0.737 | 0.837 | IMX-BVN-3 |
| RaceEthnicity | Black or African descent | 0.706 | 0.555 | 0.829 | IMX-BVN-3 |
| RaceEthnicity | Asian | 0.974 | 0.872 | 0.999 | IMX-BVN-3 |
| RaceEthnicity | Hispanic | 0.756 | 0.652 | 0.844 | SC4 |
| RaceEthnicity | White | 0.741 | 0.691 | 0.784 | SC4 |
| RaceEthnicity | Black or African descent | 0.721 | 0.569 | 0.842 | SC4 |
| RaceEthnicity | Asian | 0.903 | 0.712 | 0.995 | SC4 |

Supplementary Table 13: **Across-group performance of the IMX-BVN-3 diagnostic classifier compared with the SeptiCyte 4-marker panel (SC4).** Performance is shown in terms of the median as well as the 2.5th (LB) and 97.5th (UB) quantiles of 5000 Bayesian bootstrap samples of the posterior distribution of across-group infected-vs.-non-infected AUC. A 95% posterior confidence interval (given by the LB and UB columns) that does not include 0 would suggest a disparity in performance between the two subgroups for the comparison of patients with infection to patients without infection.

| Subgroup Variable | Subgroup Value | Post. Median | 95% Post. CI LB | 95% Post. CI UB | Predictor |
|---|---|---|---|---|---|
| AgeBracket | Senior - Middle-Aged | -0.011 | -0.100 | 0.096 | IMX-BVN-3 |
| AgeBracket | Senior - Young Adult | -0.023 | -0.131 | 0.101 | IMX-BVN-3 |
| AgeBracket | Young Adult - Middle-Aged | 0.015 | -0.077 | 0.103 | IMX-BVN-3 |
| AgeBracket | Senior - Middle-Aged | -0.049 | -0.151 | 0.052 | SC4 |
| AgeBracket | Senior - Young Adult | -0.104 | -0.207 | 0.006 | SC4 |
| AgeBracket | Young Adult - Middle-Aged | 0.037 | -0.058 | 0.126 | SC4 |
| Sex | Female - Male | -0.085 | -0.165 | -0.012 | IMX-BVN-3 |
| Sex | Female - Male | 0.003 | -0.080 | 0.081 | SC4 |
| RaceEthnicity | Hispanic - Asian | 0.306 | 0.180 | 0.454 | IMX-BVN-3 |
| RaceEthnicity | Hispanic - Black or African descent | -0.175 | -0.331 | -0.014 | IMX-BVN-3 |
| RaceEthnicity | Hispanic - White | -0.073 | -0.174 | 0.018 | IMX-BVN-3 |

**Supplementary Table 13 – continued from previous page**

| Subgroup Variable | Subgroup Value | Post. Median | 95% Post. CI LB | 95% Post. CI UB | Predictor |
|---|---|---|---|---|---|
| RaceEthnicity | White - Asian | 0.342 | 0.239 | 0.461 | IMX-BVN-3 |
| RaceEthnicity | White - Black or African descent | -0.065 | -0.194 | 0.077 | IMX-BVN-3 |
| RaceEthnicity | Black or African descent - Asian | 0.426 | 0.283 | 0.577 | IMX-BVN-3 |
| RaceEthnicity | Hispanic - Asian | 0.024 | -0.212 | 0.223 | SC4 |
| RaceEthnicity | Hispanic - Black or African descent | -0.291 | -0.445 | -0.133 | SC4 |
| RaceEthnicity | Hispanic - White | 0.107 | 0.002 | 0.203 | SC4 |
| RaceEthnicity | White - Asian | -0.058 | -0.274 | 0.148 | SC4 |
| RaceEthnicity | White - Black or African descent | -0.386 | -0.492 | -0.271 | SC4 |
| RaceEthnicity | Black or African descent - Asian | 0.295 | 0.087 | 0.477 | SC4 |

Supplementary Table 14: **Multi-level model inferences for demographic and technical effects on marker expression**. Shown are the posterior medians and 95% credible intervals of samples of the posterior distribution of each model coefficient. The inferences provided are for the 'PP, All + Sex:Inf. + Age:Inf. + RaceEthn:Inf' model. For identifiability, the reference category was set to a White female bacterial-infected patient of average age profiled on the GPL10558 microarray platform. Inferences were generated for each marker independently.

| Marker | Effect | Post. Median | 95% Post. CI LB | 95% Post. CI UB |
|---|---|---|---|---|
| IFI27 | Intercept | -0.436 | -0.647 | -0.226 |
| IFI27 | Age | -0.044 | -0.141 | 0.061 |
| IFI27 | Asian | 0.263 | -0.011 | 0.530 |
| IFI27 | Black or African descent | 0.153 | -0.004 | 0.302 |
| IFI27 | Hispanic | -0.110 | -0.260 | 0.038 |
| IFI27 | Other | 0.118 | -0.122 | 0.354 |
| IFI27 | Unknown | 0.120 | -0.083 | 0.329 |
| IFI27 | Male | -0.033 | -0.139 | 0.070 |
| IFI27 | Viral | 1.174 | 1.051 | 1.302 |
| IFI27 | Noninfected | 0.156 | 0.022 | 0.288 |
| IFI27 | GPL570 | 0.133 | -0.234 | 0.483 |
| IFI27 | GPL6947 | 0.146 | -0.190 | 0.453 |
| IFI27 | GPL6244 | -0.265 | -1.071 | 0.530 |
| IFI27 | GPL13667 | -0.058 | -0.681 | 0.560 |
| IFI27 | GPL20844 | 0.254 | -0.535 | 1.032 |
| IFI27 | GPL6102 | 0.200 | -0.519 | 0.968 |
| IFI27 | GPL10332 | 0.092 | -0.634 | 0.768 |
| IFI27 | NanoString | -0.151 | -0.431 | 0.124 |
| IFI27 | Age:Viral | -0.060 | -0.139 | 0.018 |
| IFI27 | Age:Noninfected | 0.010 | -0.086 | 0.103 |
| IFI27 | Male:Viral | 0.006 | -0.124 | 0.140 |
| IFI27 | Male:Noninfected | -0.119 | -0.256 | 0.019 |
| IFI27 | Asian:Viral | -0.162 | -0.488 | 0.169 |
| IFI27 | Black or African descent:Viral | -0.083 | -0.295 | 0.132 |
| IFI27 | Hispanic:Viral | 0.277 | 0.072 | 0.487 |
| IFI27 | Other:Viral | -0.036 | -0.339 | 0.290 |
| IFI27 | Unknown:Viral | 0.163 | -0.147 | 0.473 |
| IFI27 | Asian:Noninfected | -0.320 | -0.655 | 0.011 |
| IFI27 | Black or African descent:Noninfected | 0.193 | -0.023 | 0.423 |
| IFI27 | Hispanic:Noninfected | 0.268 | 0.078 | 0.458 |
| IFI27 | Other:Noninfected | 0.017 | -0.278 | 0.306 |
| IFI27 | Unknown:Noninfected | -0.065 | -0.277 | 0.142 |
| ZDHHC19 | Intercept | 0.384 | 0.104 | 0.662 |
| ZDHHC19 | Age | 0.037 | -0.083 | 0.163 |
| ZDHHC19 | Asian | 0.348 | 0.065 | 0.629 |

**Supplementary Table 14 – continued from previous page**

| Marker | Effect | Post. Median | 95% Post. CI LB | 95% Post. CI UB |
|---|---|---|---|---|
| ZDHHC19 | Black or African descent | -0.072 | -0.234 | 0.091 |
| ZDHHC19 | Hispanic | 0.072 | -0.085 | 0.228 |
| ZDHHC19 | Other | 0.025 | -0.216 | 0.263 |
| ZDHHC19 | Unknown | -0.077 | -0.292 | 0.142 |
| ZDHHC19 | Male | -0.058 | -0.144 | 0.032 |
| ZDHHC19 | Viral | -0.716 | -0.843 | -0.586 |
| ZDHHC19 | Noninfected | -0.668 | -0.805 | -0.533 |
| ZDHHC19 | GPL570 | 0.061 | -0.451 | 0.578 |
| ZDHHC19 | GPL6947 | 0.092 | -0.356 | 0.511 |
| ZDHHC19 | GPL6244 | 0.583 | -0.462 | 1.663 |
| ZDHHC19 | GPL13667 | -0.138 | -0.897 | 0.627 |
| ZDHHC19 | GPL20844 | 0.240 | -0.797 | 1.283 |
| ZDHHC19 | GPL6102 | 0.238 | -0.765 | 1.242 |
| ZDHHC19 | GPL10332 | 0.714 | -0.258 | 1.754 |
| ZDHHC19 | NanoString | 0.246 | -0.145 | 0.644 |
| ZDHHC19 | Age:Viral | 0.057 | -0.029 | 0.141 |
| ZDHHC19 | Age:Noninfected | -0.099 | -0.196 | -0.002 |
| ZDHHC19 | Male:Viral | 0.049 | -0.074 | 0.170 |
| ZDHHC19 | Male:Noninfected | 0.040 | -0.094 | 0.174 |
| ZDHHC19 | Asian:Viral | -0.608 | -0.971 | -0.262 |
| ZDHHC19 | Black or African descent:Viral | -0.186 | -0.404 | 0.037 |
| ZDHHC19 | Hispanic:Viral | -0.126 | -0.339 | 0.091 |
| ZDHHC19 | Other:Viral | -0.153 | -0.465 | 0.165 |
| ZDHHC19 | Unknown:Viral | -0.006 | -0.348 | 0.338 |
| ZDHHC19 | Asian:Noninfected | -0.104 | -0.442 | 0.235 |
| ZDHHC19 | Black or African descent:Noninfected | 0.036 | -0.190 | 0.267 |
| ZDHHC19 | Hispanic:Noninfected | 0.084 | -0.114 | 0.279 |
| ZDHHC19 | Other:Noninfected | 0.033 | -0.265 | 0.341 |
| ZDHHC19 | Unknown:Noninfected | -0.245 | -0.473 | -0.017 |
| TGFBI | Intercept | -0.165 | -0.444 | 0.139 |
| TGFBI | Age | -0.048 | -0.176 | 0.077 |
| TGFBI | Asian | -0.099 | -0.423 | 0.227 |
| TGFBI | Black or African descent | 0.159 | -0.017 | 0.337 |
| TGFBI | Hispanic | 0.122 | -0.051 | 0.307 |
| TGFBI | Other | -0.116 | -0.397 | 0.162 |
| TGFBI | Unknown | -0.363 | -0.613 | -0.110 |
| TGFBI | Male | 0.079 | -0.025 | 0.185 |
| TGFBI | Viral | 0.350 | 0.204 | 0.494 |
| TGFBI | Noninfected | 0.525 | 0.372 | 0.678 |
| TGFBI | GPL570 | -0.188 | -0.679 | 0.324 |
| TGFBI | GPL6947 | -0.352 | -0.809 | 0.114 |
| TGFBI | GPL6244 | -1.009 | -2.133 | 0.066 |
| TGFBI | GPL13667 | -0.300 | -1.052 | 0.475 |
| TGFBI | GPL20844 | 0.470 | -0.521 | 1.516 |
| TGFBI | GPL6102 | -0.889 | -1.955 | 0.145 |
| TGFBI | GPL10332 | -0.281 | -1.260 | 0.652 |
| TGFBI | NanoString | -0.069 | -0.484 | 0.334 |
| TGFBI | Age:Viral | -0.004 | -0.098 | 0.090 |
| TGFBI | Age:Noninfected | 0.014 | -0.097 | 0.123 |
| TGFBI | Male:Viral | 0.075 | -0.073 | 0.223 |
| TGFBI | Male:Noninfected | -0.061 | -0.223 | 0.091 |
| TGFBI | Asian:Viral | 0.122 | -0.292 | 0.531 |
| TGFBI | Black or African descent:Viral | -0.096 | -0.348 | 0.149 |
| TGFBI | Hispanic:Viral | -0.288 | -0.540 | -0.049 |
| TGFBI | Other:Viral | 0.169 | -0.213 | 0.540 |

**Supplementary Table 14 – continued from previous page**

| Marker | Effect | Post. Median | 95% Post. CI LB | 95% Post. CI UB |
|---|---|---|---|---|
| TGFBI | Unknown:Viral | -0.321 | -0.682 | 0.044 |
| TGFBI | Asian:Noninfected | -0.068 | -0.461 | 0.324 |
| TGFBI | Black or African descent:Noninfected | -0.362 | -0.616 | -0.102 |
| TGFBI | Hispanic:Noninfected | -0.402 | -0.628 | -0.187 |
| TGFBI | Other:Noninfected | 0.117 | -0.235 | 0.471 |
| TGFBI | Unknown:Noninfected | 0.547 | 0.286 | 0.803 |
| CTSB | Intercept | 0.173 | -0.114 | 0.473 |
| CTSB | Age | 0.089 | -0.017 | 0.190 |
| CTSB | Asian | 0.044 | -0.279 | 0.367 |
| CTSB | Black or African descent | 0.348 | 0.173 | 0.523 |
| CTSB | Hispanic | 0.063 | -0.111 | 0.241 |
| CTSB | Other | 0.017 | -0.256 | 0.283 |
| CTSB | Unknown | -0.185 | -0.429 | 0.067 |
| CTSB | Male | 0.030 | -0.065 | 0.124 |
| CTSB | Viral | -0.393 | -0.530 | -0.250 |
| CTSB | Noninfected | -0.175 | -0.324 | -0.031 |
| CTSB | GPL570 | 0.459 | -0.116 | 1.018 |
| CTSB | GPL6947 | 0.279 | -0.233 | 0.752 |
| CTSB | GPL6244 | -0.320 | -1.416 | 0.767 |
| CTSB | GPL13667 | -0.061 | -0.873 | 0.776 |
| CTSB | GPL20844 | 0.169 | -0.825 | 1.200 |
| CTSB | GPL6102 | -0.184 | -1.199 | 0.844 |
| CTSB | GPL10332 | 0.165 | -0.803 | 1.236 |
| CTSB | NanoString | -0.160 | -0.585 | 0.239 |
| CTSB | Age:Viral | 0.158 | 0.066 | 0.250 |
| CTSB | Age:Noninfected | 0.077 | -0.035 | 0.185 |
| CTSB | Male:Viral | 0.026 | -0.110 | 0.160 |
| CTSB | Male:Noninfected | -0.076 | -0.224 | 0.068 |
| CTSB | Asian:Viral | 0.121 | -0.271 | 0.519 |
| CTSB | Black or African descent:Viral | -0.088 | -0.332 | 0.161 |
| CTSB | Hispanic:Viral | 0.027 | -0.217 | 0.274 |
| CTSB | Other:Viral | -0.045 | -0.405 | 0.320 |
| CTSB | Unknown:Viral | -0.053 | -0.415 | 0.316 |
| CTSB | Asian:Noninfected | 0.073 | -0.320 | 0.468 |
| CTSB | Black or African descent:Noninfected | -0.026 | -0.286 | 0.224 |
| CTSB | Hispanic:Noninfected | 0.119 | -0.103 | 0.343 |
| CTSB | Other:Noninfected | 0.269 | -0.080 | 0.612 |
| CTSB | Unknown:Noninfected | 0.172 | -0.082 | 0.423 |
| CD163 | Intercept | 0.132 | -0.106 | 0.383 |
| CD163 | Age | 0.092 | -0.060 | 0.239 |
| CD163 | Asian | 0.004 | -0.312 | 0.315 |
| CD163 | Black or African descent | -0.105 | -0.287 | 0.069 |
| CD163 | Hispanic | -0.188 | -0.361 | -0.020 |
| CD163 | Other | 0.006 | -0.267 | 0.283 |
| CD163 | Unknown | -0.021 | -0.262 | 0.212 |
| CD163 | Male | -0.083 | -0.181 | 0.022 |
| CD163 | Viral | -0.309 | -0.451 | -0.168 |
| CD163 | Noninfected | -0.423 | -0.574 | -0.268 |
| CD163 | GPL570 | 0.241 | -0.195 | 0.644 |
| CD163 | GPL6947 | 0.001 | -0.383 | 0.380 |
| CD163 | GPL6244 | -0.748 | -1.682 | 0.145 |
| CD163 | GPL13667 | 0.481 | -0.206 | 1.179 |
| CD163 | GPL20844 | -0.149 | -0.988 | 0.683 |
| CD163 | GPL6102 | -0.132 | -0.969 | 0.681 |
| CD163 | GPL10332 | -0.337 | -1.184 | 0.462 |

**Supplementary Table 14 – continued from previous page**

| Marker | Effect | Post. Median | 95% Post. CI LB | 95% Post. CI UB |
|---|---|---|---|---|
| CD163 | NanoString | -0.004 | -0.319 | 0.308 |
| CD163 | Age:Viral | 0.274 | 0.177 | 0.370 |
| CD163 | Age:Noninfected | 0.194 | 0.082 | 0.306 |
| CD163 | Male:Viral | 0.034 | -0.113 | 0.179 |
| CD163 | Male:Noninfected | 0.019 | -0.138 | 0.174 |
| CD163 | Asian:Viral | -0.010 | -0.391 | 0.385 |
| CD163 | Black or African descent:Viral | -0.009 | -0.256 | 0.241 |
| CD163 | Hispanic:Viral | 0.089 | -0.143 | 0.326 |
| CD163 | Other:Viral | 0.038 | -0.322 | 0.398 |
| CD163 | Unknown:Viral | -0.240 | -0.614 | 0.135 |
| CD163 | Asian:Noninfected | -0.080 | -0.459 | 0.304 |
| CD163 | Black or African descent:Noninfected | 0.172 | -0.088 | 0.431 |
| CD163 | Hispanic:Noninfected | 0.170 | -0.045 | 0.392 |
| CD163 | Other:Noninfected | -0.189 | -0.530 | 0.151 |
| CD163 | Unknown:Noninfected | -0.009 | -0.257 | 0.237 |
| GADD45A | Intercept | 0.054 | -0.241 | 0.342 |
| GADD45A | Age | 0.226 | 0.107 | 0.338 |
| GADD45A | Asian | 0.100 | -0.159 | 0.350 |
| GADD45A | Black or African descent | 0.032 | -0.110 | 0.169 |
| GADD45A | Hispanic | -0.089 | -0.229 | 0.049 |
| GADD45A | Other | -0.007 | -0.224 | 0.206 |
| GADD45A | Unknown | -0.121 | -0.311 | 0.072 |
| GADD45A | Male | -0.017 | -0.099 | 0.066 |
| GADD45A | Viral | -0.604 | -0.717 | -0.490 |
| GADD45A | Noninfected | -0.455 | -0.578 | -0.333 |
| GADD45A | GPL570 | 1.188 | 0.651 | 1.694 |
| GADD45A | GPL6947 | 0.070 | -0.381 | 0.519 |
| GADD45A | GPL6244 | 0.540 | -0.463 | 1.574 |
| GADD45A | GPL13667 | 0.698 | -0.176 | 1.550 |
| GADD45A | GPL20844 | 0.043 | -0.984 | 1.042 |
| GADD45A | GPL6102 | 0.192 | -0.776 | 1.152 |
| GADD45A | GPL10332 | 1.287 | 0.245 | 2.334 |
| GADD45A | NanoString | 0.178 | -0.231 | 0.598 |
| GADD45A | Age:Viral | -0.062 | -0.139 | 0.015 |
| GADD45A | Age:Noninfected | -0.131 | -0.215 | -0.043 |
| GADD45A | Male:Viral | 0.008 | -0.106 | 0.121 |
| GADD45A | Male:Noninfected | 0.017 | -0.104 | 0.138 |
| GADD45A | Asian:Viral | -0.111 | -0.438 | 0.213 |
| GADD45A | Black or African descent:Viral | -0.047 | -0.241 | 0.152 |
| GADD45A | Hispanic:Viral | 0.074 | -0.112 | 0.267 |
| GADD45A | Other:Viral | -0.019 | -0.295 | 0.273 |
| GADD45A | Unknown:Viral | 0.272 | -0.033 | 0.579 |
| GADD45A | Asian:Noninfected | 0.070 | -0.231 | 0.387 |
| GADD45A | Black or African descent:Noninfected | 0.081 | -0.133 | 0.292 |
| GADD45A | Hispanic:Noninfected | 0.155 | -0.022 | 0.335 |
| GADD45A | Other:Noninfected | -0.025 | -0.295 | 0.248 |
| GADD45A | Unknown:Noninfected | -0.024 | -0.216 | 0.169 |
| CTSL1 | Intercept | 0.000 | -0.306 | 0.301 |
| CTSL1 | Age | 0.011 | -0.125 | 0.147 |
| CTSL1 | Asian | 0.345 | 0.036 | 0.651 |
| CTSL1 | Black or African descent | -0.020 | -0.183 | 0.146 |
| CTSL1 | Hispanic | -0.165 | -0.327 | -0.000 |
| CTSL1 | Other | -0.058 | -0.319 | 0.203 |
| CTSL1 | Unknown | -0.365 | -0.602 | -0.133 |
| CTSL1 | Male | 0.022 | -0.066 | 0.111 |

**Supplementary Table 14 – continued from previous page**

| Marker | Effect | Post. Median | 95% Post. CI LB | 95% Post. CI UB |
|---|---|---|---|---|
| CTSL1 | Viral | 0.110 | -0.021 | 0.248 |
| CTSL1 | Noninfected | -0.402 | -0.538 | -0.261 |
| CTSL1 | GPL570 | 0.287 | -0.261 | 0.854 |
| CTSL1 | GPL6947 | -0.306 | -0.766 | 0.166 |
| CTSL1 | GPL6244 | -0.270 | -1.421 | 0.869 |
| CTSL1 | GPL13667 | -0.238 | -1.053 | 0.574 |
| CTSL1 | GPL20844 | 0.302 | -0.815 | 1.498 |
| CTSL1 | GPL6102 | 0.183 | -1.003 | 1.374 |
| CTSL1 | GPL10332 | -0.104 | -1.256 | 1.003 |
| CTSL1 | NanoString | 0.227 | -0.191 | 0.641 |
| CTSL1 | Age:Viral | 0.002 | -0.087 | 0.089 |
| CTSL1 | Age:Noninfected | -0.083 | -0.187 | 0.021 |
| CTSL1 | Male:Viral | 0.034 | -0.093 | 0.165 |
| CTSL1 | Male:Noninfected | 0.084 | -0.055 | 0.221 |
| CTSL1 | Asian:Viral | -0.116 | -0.492 | 0.268 |
| CTSL1 | Black or African descent:Viral | 0.315 | 0.078 | 0.542 |
| CTSL1 | Hispanic:Viral | 0.257 | 0.033 | 0.475 |
| CTSL1 | Other:Viral | 0.090 | -0.260 | 0.448 |
| CTSL1 | Unknown:Viral | 0.385 | 0.015 | 0.745 |
| CTSL1 | Asian:Noninfected | -0.024 | -0.389 | 0.355 |
| CTSL1 | Black or African descent:Noninfected | 0.178 | -0.060 | 0.417 |
| CTSL1 | Hispanic:Noninfected | 0.111 | -0.095 | 0.317 |
| CTSL1 | Other:Noninfected | 0.060 | -0.264 | 0.382 |
| CTSL1 | Unknown:Noninfected | 0.237 | -0.000 | 0.477 |
| DEFA4 | Intercept | 0.141 | -0.192 | 0.454 |
| DEFA4 | Age | -0.062 | -0.226 | 0.100 |
| DEFA4 | Asian | 0.186 | -0.155 | 0.522 |
| DEFA4 | Black or African descent | -0.041 | -0.230 | 0.148 |
| DEFA4 | Hispanic | -0.183 | -0.374 | 0.009 |
| DEFA4 | Other | -0.155 | -0.442 | 0.144 |
| DEFA4 | Unknown | 0.198 | -0.072 | 0.454 |
| DEFA4 | Male | 0.122 | 0.013 | 0.235 |
| DEFA4 | Viral | -0.130 | -0.285 | 0.028 |
| DEFA4 | Noninfected | -0.215 | -0.375 | -0.049 |
| DEFA4 | GPL570 | -0.175 | -0.722 | 0.377 |
| DEFA4 | GPL6947 | 0.134 | -0.409 | 0.645 |
| DEFA4 | GPL6244 | 0.019 | -1.110 | 1.114 |
| DEFA4 | GPL13667 | -0.158 | -1.054 | 0.716 |
| DEFA4 | GPL20844 | -0.404 | -1.725 | 0.768 |
| DEFA4 | GPL6102 | -0.464 | -1.636 | 0.582 |
| DEFA4 | GPL10332 | 0.322 | -0.682 | 1.418 |
| DEFA4 | NanoString | -0.543 | -0.983 | -0.127 |
| DEFA4 | Age:Viral | 0.031 | -0.069 | 0.133 |
| DEFA4 | Age:Noninfected | 0.104 | -0.019 | 0.224 |
| DEFA4 | Male:Viral | -0.101 | -0.265 | 0.061 |
| DEFA4 | Male:Noninfected | -0.011 | -0.173 | 0.152 |
| DEFA4 | Asian:Viral | -0.210 | -0.627 | 0.212 |
| DEFA4 | Black or African descent:Viral | -0.153 | -0.415 | 0.114 |
| DEFA4 | Hispanic:Viral | 0.138 | -0.118 | 0.400 |
| DEFA4 | Other:Viral | 0.035 | -0.357 | 0.411 |
| DEFA4 | Unknown:Viral | -0.227 | -0.644 | 0.178 |
| DEFA4 | Asian:Noninfected | -0.387 | -0.800 | 0.022 |
| DEFA4 | Black or African descent:Noninfected | -0.049 | -0.324 | 0.229 |
| DEFA4 | Hispanic:Noninfected | 0.031 | -0.205 | 0.272 |
| DEFA4 | Other:Noninfected | -0.042 | -0.402 | 0.313 |

**Supplementary Table 14 – continued from previous page**

| Marker | Effect | Post. Median | 95% Post. CI LB | 95% Post. CI UB |
|---|---|---|---|---|
| DEFA4 | Unknown:Noninfected | -0.305 | -0.568 | -0.037 |
| PDE4B | Intercept | -0.128 | -0.459 | 0.223 |
| PDE4B | Age | -0.063 | -0.202 | 0.069 |
| PDE4B | Asian | 0.208 | -0.086 | 0.501 |
| PDE4B | Black or African descent | -0.207 | -0.363 | -0.048 |
| PDE4B | Hispanic | -0.015 | -0.174 | 0.140 |
| PDE4B | Other | 0.004 | -0.238 | 0.242 |
| PDE4B | Unknown | -0.253 | -0.470 | -0.025 |
| PDE4B | Male | -0.154 | -0.247 | -0.065 |
| PDE4B | Viral | 0.070 | -0.060 | 0.200 |
| PDE4B | Noninfected | 0.196 | 0.061 | 0.328 |
| PDE4B | GPL570 | 0.035 | -0.604 | 0.656 |
| PDE4B | GPL6947 | 0.245 | -0.324 | 0.801 |
| PDE4B | GPL6244 | 0.240 | -0.883 | 1.383 |
| PDE4B | GPL13667 | 0.257 | -0.695 | 1.193 |
| PDE4B | GPL20844 | 0.189 | -0.915 | 1.286 |
| PDE4B | GPL6102 | 0.513 | -0.568 | 1.693 |
| PDE4B | GPL10332 | -0.301 | -1.422 | 0.796 |
| PDE4B | NanoString | 0.711 | 0.232 | 1.165 |
| PDE4B | Age:Viral | -0.002 | -0.086 | 0.082 |
| PDE4B | Age:Noninfected | -0.040 | -0.137 | 0.060 |
| PDE4B | Male:Viral | 0.047 | -0.076 | 0.175 |
| PDE4B | Male:Noninfected | 0.094 | -0.039 | 0.233 |
| PDE4B | Asian:Viral | -0.081 | -0.443 | 0.289 |
| PDE4B | Black or African descent:Viral | 0.210 | -0.018 | 0.425 |
| PDE4B | Hispanic:Viral | 0.041 | -0.165 | 0.256 |
| PDE4B | Other:Viral | -0.151 | -0.457 | 0.178 |
| PDE4B | Unknown:Viral | 0.144 | -0.184 | 0.493 |
| PDE4B | Asian:Noninfected | 0.087 | -0.269 | 0.440 |
| PDE4B | Black or African descent:Noninfected | 0.223 | -0.009 | 0.452 |
| PDE4B | Hispanic:Noninfected | 0.114 | -0.075 | 0.312 |
| PDE4B | Other:Noninfected | 0.024 | -0.271 | 0.325 |
| PDE4B | Unknown:Noninfected | 0.369 | 0.149 | 0.588 |
| FURIN | Intercept | 0.385 | 0.055 | 0.707 |
| FURIN | Age | 0.142 | 0.014 | 0.267 |
| FURIN | Asian | -0.026 | -0.336 | 0.278 |
| FURIN | Black or African descent | 0.127 | -0.040 | 0.297 |
| FURIN | Hispanic | -0.016 | -0.184 | 0.148 |
| FURIN | Other | 0.060 | -0.200 | 0.313 |
| FURIN | Unknown | -0.120 | -0.356 | 0.108 |
| FURIN | Male | -0.034 | -0.117 | 0.059 |
| FURIN | Viral | -0.564 | -0.696 | -0.427 |
| FURIN | Noninfected | -0.237 | -0.381 | -0.095 |
| FURIN | GPL570 | -0.124 | -0.692 | 0.477 |
| FURIN | GPL6947 | 0.393 | -0.185 | 1.014 |
| FURIN | GPL6244 | -0.381 | -1.493 | 0.654 |
| FURIN | GPL13667 | 0.165 | -0.721 | 1.035 |
| FURIN | GPL20844 | -0.539 | -1.642 | 0.508 |
| FURIN | GPL6102 | 0.078 | -0.915 | 1.138 |
| FURIN | GPL10332 | 0.566 | -0.403 | 1.647 |
| FURIN | NanoString | -0.522 | -0.973 | -0.106 |
| FURIN | Age:Viral | 0.054 | -0.034 | 0.139 |
| FURIN | Age:Noninfected | 0.052 | -0.049 | 0.156 |
| FURIN | Male:Viral | 0.080 | -0.048 | 0.202 |
| FURIN | Male:Noninfected | 0.028 | -0.108 | 0.161 |

**Supplementary Table 14 – continued from previous page**

| Marker | Effect | Post. Median | 95% Post. CI LB | 95% Post. CI UB |
|---|---|---|---|---|
| FURIN | Asian:Viral | -0.058 | -0.435 | 0.331 |
| FURIN | Black or African descent:Viral | -0.240 | -0.474 | -0.006 |
| FURIN | Hispanic:Viral | -0.070 | -0.307 | 0.160 |
| FURIN | Other:Viral | -0.074 | -0.422 | 0.271 |
| FURIN | Unknown:Viral | -0.029 | -0.399 | 0.325 |
| FURIN | Asian:Noninfected | 0.077 | -0.284 | 0.450 |
| FURIN | Black or African descent:Noninfected | -0.053 | -0.312 | 0.191 |
| FURIN | Hispanic:Noninfected | 0.163 | -0.043 | 0.378 |
| FURIN | Other:Noninfected | 0.041 | -0.283 | 0.373 |
| FURIN | Unknown:Noninfected | 0.001 | -0.246 | 0.242 |
| GNA15 | Intercept | 0.555 | 0.317 | 0.795 |
| GNA15 | Age | -0.020 | -0.124 | 0.082 |
| GNA15 | Asian | 0.197 | -0.112 | 0.518 |
| GNA15 | Black or African descent | 0.056 | -0.134 | 0.241 |
| GNA15 | Hispanic | 0.004 | -0.178 | 0.190 |
| GNA15 | Other | -0.085 | -0.381 | 0.201 |
| GNA15 | Unknown | -0.062 | -0.303 | 0.182 |
| GNA15 | Male | 0.032 | -0.065 | 0.128 |
| GNA15 | Viral | -0.692 | -0.830 | -0.546 |
| GNA15 | Noninfected | -0.658 | -0.810 | -0.508 |
| GNA15 | GPL570 | 0.300 | -0.107 | 0.727 |
| GNA15 | GPL6947 | 0.070 | -0.312 | 0.452 |
| GNA15 | GPL6244 | -0.604 | -1.526 | 0.293 |
| GNA15 | GPL13667 | -0.339 | -0.997 | 0.302 |
| GNA15 | GPL20844 | -0.492 | -1.351 | 0.334 |
| GNA15 | GPL6102 | -0.094 | -0.923 | 0.744 |
| GNA15 | GPL10332 | -0.081 | -0.829 | 0.660 |
| GNA15 | NanoString | -0.516 | -0.827 | -0.209 |
| GNA15 | Age:Viral | 0.295 | 0.203 | 0.389 |
| GNA15 | Age:Noninfected | -0.028 | -0.141 | 0.084 |
| GNA15 | Male:Viral | 0.081 | -0.057 | 0.220 |
| GNA15 | Male:Noninfected | 0.020 | -0.126 | 0.170 |
| GNA15 | Asian:Viral | -0.372 | -0.780 | 0.027 |
| GNA15 | Black or African descent:Viral | -0.079 | -0.333 | 0.188 |
| GNA15 | Hispanic:Viral | -0.171 | -0.428 | 0.082 |
| GNA15 | Other:Viral | 0.022 | -0.378 | 0.399 |
| GNA15 | Unknown:Viral | 0.029 | -0.339 | 0.394 |
| GNA15 | Asian:Noninfected | -0.022 | -0.424 | 0.366 |
| GNA15 | Black or African descent:Noninfected | -0.109 | -0.384 | 0.156 |
| GNA15 | Hispanic:Noninfected | -0.002 | -0.235 | 0.224 |
| GNA15 | Other:Noninfected | 0.310 | -0.046 | 0.680 |
| GNA15 | Unknown:Noninfected | 0.016 | -0.238 | 0.268 |
| OASL | Intercept | -0.267 | -0.507 | -0.049 |
| OASL | Age | -0.025 | -0.151 | 0.114 |
| OASL | Asian | 0.247 | -0.029 | 0.523 |
| OASL | Black or African descent | -0.281 | -0.431 | -0.123 |
| OASL | Hispanic | -0.159 | -0.308 | -0.012 |
| OASL | Other | 0.114 | -0.113 | 0.356 |
| OASL | Unknown | 0.018 | -0.189 | 0.232 |
| OASL | Male | -0.029 | -0.115 | 0.052 |
| OASL | Viral | 1.165 | 1.047 | 1.285 |
| OASL | Noninfected | 0.235 | 0.104 | 0.363 |
| OASL | GPL570 | -0.136 | -0.531 | 0.253 |
| OASL | GPL6947 | 0.152 | -0.199 | 0.499 |
| OASL | GPL6244 | -0.531 | -1.423 | 0.338 |

**Supplementary Table 14 – continued from previous page**

| Marker | Effect | Post. Median | 95% Post. CI LB | 95% Post. CI UB |
|---|---|---|---|---|
| OASL | GPL13667 | -0.410 | -1.074 | 0.240 |
| OASL | GPL20844 | 0.185 | -0.653 | 1.025 |
| OASL | GPL6102 | -0.030 | -0.863 | 0.793 |
| OASL | GPL10332 | -0.234 | -0.956 | 0.509 |
| OASL | NanoString | -0.147 | -0.435 | 0.166 |
| OASL | Age:Viral | -0.086 | -0.167 | -0.004 |
| OASL | Age:Noninfected | -0.170 | -0.265 | -0.075 |
| OASL | Male:Viral | -0.047 | -0.160 | 0.074 |
| OASL | Male:Noninfected | -0.144 | -0.272 | -0.018 |
| OASL | Asian:Viral | -0.001 | -0.338 | 0.342 |
| OASL | Black or African descent:Viral | 0.335 | 0.115 | 0.553 |
| OASL | Hispanic:Viral | 0.438 | 0.226 | 0.646 |
| OASL | Other:Viral | -0.018 | -0.340 | 0.293 |
| OASL | Unknown:Viral | -0.217 | -0.556 | 0.115 |
| OASL | Asian:Noninfected | -0.074 | -0.414 | 0.275 |
| OASL | Black or African descent:Noninfected | 0.396 | 0.172 | 0.618 |
| OASL | Hispanic:Noninfected | 0.298 | 0.111 | 0.488 |
| OASL | Other:Noninfected | -0.008 | -0.307 | 0.282 |
| OASL | Unknown:Noninfected | -0.095 | -0.321 | 0.124 |
| C9orf95 | Intercept | 0.064 | -0.221 | 0.346 |
| C9orf95 | Age | 0.067 | -0.070 | 0.208 |
| C9orf95 | Asian | -0.036 | -0.365 | 0.300 |
| C9orf95 | Black or African descent | 0.245 | 0.048 | 0.442 |
| C9orf95 | Hispanic | 0.143 | -0.054 | 0.334 |
| C9orf95 | Other | -0.155 | -0.457 | 0.140 |
| C9orf95 | Unknown | 0.174 | -0.094 | 0.439 |
| C9orf95 | Male | -0.087 | -0.192 | 0.016 |
| C9orf95 | Viral | 0.095 | -0.065 | 0.251 |
| C9orf95 | Noninfected | -0.322 | -0.484 | -0.160 |
| C9orf95 | GPL570 | 0.463 | -0.082 | 0.992 |
| C9orf95 | GPL6947 | -0.200 | -0.685 | 0.280 |
| C9orf95 | GPL6244 | 0.346 | -0.681 | 1.410 |
| C9orf95 | GPL13667 | -0.056 | -0.898 | 0.782 |
| C9orf95 | GPL20844 | -0.181 | -1.223 | 0.809 |
| C9orf95 | GPL6102 | 0.327 | -0.650 | 1.317 |
| C9orf95 | GPL10332 | 0.131 | -0.778 | 1.103 |
| C9orf95 | NanoString | 0.076 | -0.297 | 0.455 |
| C9orf95 | Age:Viral | -0.087 | -0.192 | 0.013 |
| C9orf95 | Age:Noninfected | -0.162 | -0.280 | -0.041 |
| C9orf95 | Male:Viral | -0.065 | -0.217 | 0.089 |
| C9orf95 | Male:Noninfected | 0.112 | -0.042 | 0.273 |
| C9orf95 | Asian:Viral | -0.100 | -0.535 | 0.326 |
| C9orf95 | Black or African descent:Viral | -0.012 | -0.291 | 0.263 |
| C9orf95 | Hispanic:Viral | -0.024 | -0.286 | 0.249 |
| C9orf95 | Other:Viral | 0.035 | -0.364 | 0.444 |
| C9orf95 | Unknown:Viral | 0.037 | -0.384 | 0.445 |
| C9orf95 | Asian:Noninfected | 0.194 | -0.210 | 0.598 |
| C9orf95 | Black or African descent:Noninfected | 0.059 | -0.221 | 0.341 |
| C9orf95 | Hispanic:Noninfected | 0.103 | -0.141 | 0.352 |
| C9orf95 | Other:Noninfected | 0.159 | -0.218 | 0.535 |
| C9orf95 | Unknown:Noninfected | -0.154 | -0.429 | 0.124 |
| CEACAM1 | Intercept | 0.258 | -0.034 | 0.545 |
| CEACAM1 | Age | -0.101 | -0.213 | 0.012 |
| CEACAM1 | Asian | 0.108 | -0.208 | 0.426 |
| CEACAM1 | Black or African descent | -0.182 | -0.365 | -0.000 |

**Supplementary Table 14 – continued from previous page**

| Marker | Effect | Post. Median | 95% Post. CI LB | 95% Post. CI UB |
|---|---|---|---|---|
| CEACAM1 | Hispanic | -0.178 | -0.359 | -0.002 |
| CEACAM1 | Other | 0.028 | -0.243 | 0.303 |
| CEACAM1 | Unknown | -0.288 | -0.537 | -0.043 |
| CEACAM1 | Male | -0.047 | -0.137 | 0.044 |
| CEACAM1 | Viral | -0.334 | -0.476 | -0.192 |
| CEACAM1 | Noninfected | -0.659 | -0.806 | -0.508 |
| CEACAM1 | GPL570 | 0.379 | -0.125 | 0.908 |
| CEACAM1 | GPL6947 | -0.250 | -0.691 | 0.214 |
| CEACAM1 | GPL6244 | -0.301 | -1.350 | 0.699 |
| CEACAM1 | GPL13667 | 0.270 | -0.481 | 1.047 |
| CEACAM1 | GPL20844 | -0.321 | -1.357 | 0.638 |
| CEACAM1 | GPL6102 | 0.106 | -0.887 | 1.074 |
| CEACAM1 | GPL10332 | 0.207 | -0.719 | 1.163 |
| CEACAM1 | NanoString | 0.174 | -0.216 | 0.562 |
| CEACAM1 | Age:Viral | 0.156 | 0.060 | 0.251 |
| CEACAM1 | Age:Noninfected | -0.317 | -0.427 | -0.208 |
| CEACAM1 | Male:Viral | -0.045 | -0.175 | 0.085 |
| CEACAM1 | Male:Noninfected | 0.014 | -0.129 | 0.156 |
| CEACAM1 | Asian:Viral | 0.054 | -0.350 | 0.448 |
| CEACAM1 | Black or African descent:Viral | 0.026 | -0.236 | 0.272 |
| CEACAM1 | Hispanic:Viral | 0.276 | 0.029 | 0.524 |
| CEACAM1 | Other:Viral | 0.061 | -0.299 | 0.440 |
| CEACAM1 | Unknown:Viral | 0.152 | -0.209 | 0.527 |
| CEACAM1 | Asian:Noninfected | 0.228 | -0.164 | 0.605 |
| CEACAM1 | Black or African descent:Noninfected | 0.090 | -0.177 | 0.356 |
| CEACAM1 | Hispanic:Noninfected | 0.436 | 0.208 | 0.656 |
| CEACAM1 | Other:Noninfected | 0.206 | -0.140 | 0.551 |
| CEACAM1 | Unknown:Noninfected | 0.360 | 0.107 | 0.612 |
| OLFM4 | Intercept | -0.053 | -0.311 | 0.191 |
| OLFM4 | Age | 0.077 | -0.058 | 0.211 |
| OLFM4 | Asian | 0.033 | -0.245 | 0.310 |
| OLFM4 | Black or African descent | 0.068 | -0.092 | 0.228 |
| OLFM4 | Hispanic | -0.128 | -0.284 | 0.032 |
| OLFM4 | Other | -0.041 | -0.277 | 0.203 |
| OLFM4 | Unknown | 0.111 | -0.107 | 0.330 |
| OLFM4 | Male | 0.042 | -0.053 | 0.145 |
| OLFM4 | Viral | -0.491 | -0.618 | -0.365 |
| OLFM4 | Noninfected | -0.507 | -0.647 | -0.366 |
| OLFM4 | GPL570 | 1.307 | 0.871 | 1.725 |
| OLFM4 | GPL6947 | 0.053 | -0.342 | 0.451 |
| OLFM4 | GPL6244 | 0.417 | -0.425 | 1.296 |
| OLFM4 | GPL13667 | 0.555 | -0.157 | 1.251 |
| OLFM4 | GPL20844 | -0.168 | -1.173 | 0.770 |
| OLFM4 | GPL6102 | -0.219 | -1.193 | 0.658 |
| OLFM4 | GPL10332 | 1.021 | 0.204 | 1.835 |
| OLFM4 | NanoString | 0.487 | 0.108 | 0.864 |
| OLFM4 | Age:Viral | -0.028 | -0.112 | 0.055 |
| OLFM4 | Age:Noninfected | -0.070 | -0.169 | 0.029 |
| OLFM4 | Male:Viral | -0.021 | -0.154 | 0.109 |
| OLFM4 | Male:Noninfected | -0.003 | -0.154 | 0.137 |
| OLFM4 | Asian:Viral | -0.285 | -0.628 | 0.060 |
| OLFM4 | Black or African descent:Viral | -0.167 | -0.390 | 0.055 |
| OLFM4 | Hispanic:Viral | 0.131 | -0.087 | 0.348 |
| OLFM4 | Other:Viral | -0.073 | -0.390 | 0.251 |
| OLFM4 | Unknown:Viral | -0.048 | -0.382 | 0.290 |

**Supplementary Table 14 – continued from previous page**

| Marker | Effect | Post. Median | 95% Post. CI LB | 95% Post. CI UB |
|---|---|---|---|---|
| OLFM4 | Asian:Noninfected | -0.048 | -0.397 | 0.294 |
| OLFM4 | Black or African descent:Noninfected | -0.103 | -0.339 | 0.124 |
| OLFM4 | Hispanic:Noninfected | 0.154 | -0.048 | 0.347 |
| OLFM4 | Other:Noninfected | -0.102 | -0.406 | 0.203 |
| OLFM4 | Unknown:Noninfected | -0.240 | -0.467 | -0.014 |
| HLA-DMB | Intercept | -0.335 | -0.608 | -0.056 |
| HLA-DMB | Age | -0.216 | -0.316 | -0.121 |
| HLA-DMB | Asian | -0.029 | -0.324 | 0.277 |
| HLA-DMB | Black or African descent | 0.018 | -0.153 | 0.187 |
| HLA-DMB | Hispanic | 0.060 | -0.112 | 0.220 |
| HLA-DMB | Other | -0.143 | -0.397 | 0.119 |
| HLA-DMB | Unknown | -0.068 | -0.291 | 0.162 |
| HLA-DMB | Male | -0.000 | -0.102 | 0.099 |
| HLA-DMB | Viral | 0.526 | 0.391 | 0.662 |
| HLA-DMB | Noninfected | 0.556 | 0.418 | 0.700 |
| HLA-DMB | GPL570 | -0.348 | -0.827 | 0.122 |
| HLA-DMB | GPL6947 | -0.150 | -0.593 | 0.290 |
| HLA-DMB | GPL6244 | -0.316 | -1.283 | 0.614 |
| HLA-DMB | GPL13667 | -0.486 | -1.203 | 0.236 |
| HLA-DMB | GPL20844 | 0.434 | -0.435 | 1.354 |
| HLA-DMB | GPL6102 | -0.349 | -1.234 | 0.505 |
| HLA-DMB | GPL10332 | -0.463 | -1.311 | 0.415 |
| HLA-DMB | NanoString | 0.415 | 0.039 | 0.792 |
| HLA-DMB | Age:Viral | 0.127 | 0.040 | 0.214 |
| HLA-DMB | Age:Noninfected | 0.106 | 0.005 | 0.208 |
| HLA-DMB | Male:Viral | 0.009 | -0.134 | 0.148 |
| HLA-DMB | Male:Noninfected | -0.015 | -0.159 | 0.129 |
| HLA-DMB | Asian:Viral | 0.080 | -0.293 | 0.445 |
| HLA-DMB | Black or African descent:Viral | -0.008 | -0.242 | 0.229 |
| HLA-DMB | Hispanic:Viral | -0.057 | -0.282 | 0.177 |
| HLA-DMB | Other:Viral | 0.244 | -0.105 | 0.580 |
| HLA-DMB | Unknown:Viral | -0.024 | -0.377 | 0.323 |
| HLA-DMB | Asian:Noninfected | -0.165 | -0.526 | 0.198 |
| HLA-DMB | Black or African descent:Noninfected | -0.231 | -0.475 | 0.013 |
| HLA-DMB | Hispanic:Noninfected | -0.290 | -0.495 | -0.079 |
| HLA-DMB | Other:Noninfected | 0.095 | -0.232 | 0.416 |
| HLA-DMB | Unknown:Noninfected | 0.127 | -0.110 | 0.370 |
| RAPGEF1 | Intercept | -0.177 | -0.488 | 0.130 |
| RAPGEF1 | Age | -0.030 | -0.139 | 0.066 |
| RAPGEF1 | Asian | -0.302 | -0.592 | -0.008 |
| RAPGEF1 | Black or African descent | 0.193 | 0.030 | 0.355 |
| RAPGEF1 | Hispanic | 0.067 | -0.089 | 0.229 |
| RAPGEF1 | Other | 0.077 | -0.170 | 0.321 |
| RAPGEF1 | Unknown | 0.191 | -0.033 | 0.423 |
| RAPGEF1 | Male | -0.049 | -0.142 | 0.040 |
| RAPGEF1 | Viral | 0.053 | -0.073 | 0.181 |
| RAPGEF1 | Noninfected | 0.319 | 0.183 | 0.456 |
| RAPGEF1 | GPL570 | 0.693 | 0.091 | 1.259 |
| RAPGEF1 | GPL6947 | 0.129 | -0.357 | 0.615 |
| RAPGEF1 | GPL6244 | 0.055 | -1.009 | 1.111 |
| RAPGEF1 | GPL13667 | 0.363 | -0.463 | 1.195 |
| RAPGEF1 | GPL20844 | 0.081 | -0.978 | 1.192 |
| RAPGEF1 | GPL6102 | -0.195 | -1.224 | 0.789 |
| RAPGEF1 | GPL10332 | -0.188 | -1.176 | 0.760 |
| RAPGEF1 | NanoString | 0.244 | -0.200 | 0.677 |

**Supplementary Table 14 – continued from previous page**

| Marker | Effect | Post. Median | 95% Post. CI LB | 95% Post. CI UB |
|---|---|---|---|---|
| RAPGEF1 | Age:Viral | 0.111 | 0.030 | 0.190 |
| RAPGEF1 | Age:Noninfected | 0.198 | 0.099 | 0.295 |
| RAPGEF1 | Male:Viral | 0.113 | -0.012 | 0.240 |
| RAPGEF1 | Male:Noninfected | 0.046 | -0.085 | 0.185 |
| RAPGEF1 | Asian:Viral | 0.173 | -0.180 | 0.522 |
| RAPGEF1 | Black or African descent:Viral | -0.153 | -0.379 | 0.072 |
| RAPGEF1 | Hispanic:Viral | -0.294 | -0.514 | -0.076 |
| RAPGEF1 | Other:Viral | -0.072 | -0.413 | 0.256 |
| RAPGEF1 | Unknown:Viral | -0.480 | -0.839 | -0.145 |
| RAPGEF1 | Asian:Noninfected | -0.007 | -0.367 | 0.339 |
| RAPGEF1 | Black or African descent:Noninfected | -0.292 | -0.531 | -0.063 |
| RAPGEF1 | Hispanic:Noninfected | -0.339 | -0.534 | -0.147 |
| RAPGEF1 | Other:Noninfected | -0.287 | -0.589 | 0.019 |
| RAPGEF1 | Unknown:Noninfected | -0.511 | -0.738 | -0.284 |
| PER1 | Intercept | 0.084 | -0.139 | 0.308 |
| PER1 | Age | 0.166 | 0.038 | 0.296 |
| PER1 | Asian | -0.212 | -0.560 | 0.112 |
| PER1 | Black or African descent | -0.044 | -0.228 | 0.144 |
| PER1 | Hispanic | -0.102 | -0.287 | 0.090 |
| PER1 | Other | -0.083 | -0.369 | 0.223 |
| PER1 | Unknown | -0.067 | -0.323 | 0.182 |
| PER1 | Male | 0.026 | -0.073 | 0.126 |
| PER1 | Viral | -0.009 | -0.162 | 0.142 |
| PER1 | Noninfected | -0.106 | -0.266 | 0.050 |
| PER1 | GPL570 | 0.005 | -0.372 | 0.375 |
| PER1 | GPL6947 | -0.001 | -0.359 | 0.373 |
| PER1 | GPL6244 | -0.354 | -1.231 | 0.502 |
| PER1 | GPL13667 | 0.094 | -0.535 | 0.736 |
| PER1 | GPL20844 | 0.029 | -0.751 | 0.824 |
| PER1 | GPL6102 | 0.020 | -0.730 | 0.757 |
| PER1 | GPL10332 | -0.992 | -1.908 | -0.177 |
| PER1 | NanoString | -0.098 | -0.392 | 0.202 |
| PER1 | Age:Viral | 0.229 | 0.128 | 0.330 |
| PER1 | Age:Noninfected | 0.032 | -0.084 | 0.146 |
| PER1 | Male:Viral | 0.001 | -0.145 | 0.140 |
| PER1 | Male:Noninfected | -0.047 | -0.206 | 0.102 |
| PER1 | Asian:Viral | 0.070 | -0.326 | 0.474 |
| PER1 | Black or African descent:Viral | -0.076 | -0.341 | 0.183 |
| PER1 | Hispanic:Viral | 0.297 | 0.040 | 0.555 |
| PER1 | Other:Viral | 0.244 | -0.146 | 0.636 |
| PER1 | Unknown:Viral | 0.011 | -0.370 | 0.383 |
| PER1 | Asian:Noninfected | 0.167 | -0.236 | 0.586 |
| PER1 | Black or African descent:Noninfected | 0.139 | -0.132 | 0.418 |
| PER1 | Hispanic:Noninfected | 0.052 | -0.188 | 0.291 |
| PER1 | Other:Noninfected | -0.066 | -0.441 | 0.292 |
| PER1 | Unknown:Noninfected | -0.225 | -0.492 | 0.036 |
| ARG1 | Intercept | 0.141 | -0.170 | 0.461 |
| ARG1 | Age | 0.103 | -0.043 | 0.256 |
| ARG1 | Asian | 0.107 | -0.173 | 0.383 |
| ARG1 | Black or African descent | 0.088 | -0.063 | 0.239 |
| ARG1 | Hispanic | -0.076 | -0.224 | 0.073 |
| ARG1 | Other | 0.061 | -0.167 | 0.296 |
| ARG1 | Unknown | -0.143 | -0.357 | 0.061 |
| ARG1 | Male | 0.031 | -0.059 | 0.121 |
| ARG1 | Viral | -0.581 | -0.701 | -0.454 |

**Supplementary Table 14 – continued from previous page**

| Marker | Effect | Post. Median | 95% Post. CI LB | 95% Post. CI UB |
|---|---|---|---|---|
| ARG1 | Noninfected | -0.427 | -0.559 | -0.293 |
| ARG1 | GPL570 | 0.852 | 0.341 | 1.372 |
| ARG1 | GPL6947 | 0.012 | -0.444 | 0.453 |
| ARG1 | GPL6244 | -0.092 | -1.238 | 1.005 |
| ARG1 | GPL13667 | 0.681 | -0.149 | 1.606 |
| ARG1 | GPL20844 | -0.454 | -1.600 | 0.627 |
| ARG1 | GPL6102 | -0.575 | -1.723 | 0.465 |
| ARG1 | GPL10332 | 0.716 | -0.236 | 1.745 |
| ARG1 | NanoString | -0.272 | -0.768 | 0.185 |
| ARG1 | Age:Viral | 0.036 | -0.046 | 0.115 |
| ARG1 | Age:Noninfected | 0.003 | -0.091 | 0.098 |
| ARG1 | Male:Viral | -0.035 | -0.159 | 0.088 |
| ARG1 | Male:Noninfected | -0.037 | -0.169 | 0.090 |
| ARG1 | Asian:Viral | -0.176 | -0.520 | 0.172 |
| ARG1 | Black or African descent:Viral | -0.162 | -0.378 | 0.048 |
| ARG1 | Hispanic:Viral | 0.051 | -0.162 | 0.255 |
| ARG1 | Other:Viral | 0.033 | -0.277 | 0.339 |
| ARG1 | Unknown:Viral | 0.289 | -0.044 | 0.615 |
| ARG1 | Asian:Noninfected | 0.053 | -0.291 | 0.388 |
| ARG1 | Black or African descent:Noninfected | 0.035 | -0.182 | 0.255 |
| ARG1 | Hispanic:Noninfected | 0.157 | -0.031 | 0.350 |
| ARG1 | Other:Noninfected | -0.041 | -0.331 | 0.248 |
| ARG1 | Unknown:Noninfected | -0.012 | -0.229 | 0.198 |
| KCNJ2 | Intercept | 0.408 | 0.078 | 0.732 |
| KCNJ2 | Age | 0.038 | -0.137 | 0.216 |
| KCNJ2 | Asian | 0.005 | -0.340 | 0.357 |
| KCNJ2 | Black or African descent | -0.141 | -0.328 | 0.043 |
| KCNJ2 | Hispanic | 0.092 | -0.090 | 0.278 |
| KCNJ2 | Other | -0.044 | -0.331 | 0.242 |
| KCNJ2 | Unknown | -0.398 | -0.660 | -0.143 |
| KCNJ2 | Male | -0.064 | -0.163 | 0.040 |
| KCNJ2 | Viral | -0.340 | -0.489 | -0.189 |
| KCNJ2 | Noninfected | -0.368 | -0.522 | -0.211 |
| KCNJ2 | GPL570 | -0.368 | -1.007 | 0.258 |
| KCNJ2 | GPL6947 | 0.133 | -0.341 | 0.624 |
| KCNJ2 | GPL6244 | -0.671 | -2.106 | 0.512 |
| KCNJ2 | GPL13667 | -0.168 | -1.047 | 0.720 |
| KCNJ2 | GPL20844 | -0.609 | -1.919 | 0.558 |
| KCNJ2 | GPL6102 | -0.468 | -1.766 | 0.704 |
| KCNJ2 | GPL10332 | -0.423 | -1.609 | 0.684 |
| KCNJ2 | NanoString | -0.293 | -0.722 | 0.153 |
| KCNJ2 | Age:Viral | 0.245 | 0.146 | 0.344 |
| KCNJ2 | Age:Noninfected | -0.136 | -0.255 | -0.018 |
| KCNJ2 | Male:Viral | 0.048 | -0.098 | 0.185 |
| KCNJ2 | Male:Noninfected | -0.041 | -0.200 | 0.110 |
| KCNJ2 | Asian:Viral | 0.006 | -0.428 | 0.442 |
| KCNJ2 | Black or African descent:Viral | -0.034 | -0.290 | 0.231 |
| KCNJ2 | Hispanic:Viral | -0.094 | -0.350 | 0.169 |
| KCNJ2 | Other:Viral | 0.146 | -0.232 | 0.531 |
| KCNJ2 | Unknown:Viral | 0.296 | -0.108 | 0.690 |
| KCNJ2 | Asian:Noninfected | 0.448 | 0.030 | 0.864 |
| KCNJ2 | Black or African descent:Noninfected | 0.333 | 0.054 | 0.603 |
| KCNJ2 | Hispanic:Noninfected | 0.266 | 0.039 | 0.499 |
| KCNJ2 | Other:Noninfected | 0.425 | 0.072 | 0.774 |
| KCNJ2 | Unknown:Noninfected | 0.450 | 0.185 | 0.720 |

Supplementary Table 14 – continued from previous page

| Marker | Effect | Post. Median | 95% Post. CI LB | 95% Post. CI UB |
|---|---|---|---|---|
| BATF | Intercept | 0.483 | 0.215 | 0.774 |
| BATF | Age | -0.002 | -0.123 | 0.119 |
| BATF | Asian | 0.229 | -0.071 | 0.536 |
| BATF | Black or African descent | 0.106 | -0.060 | 0.280 |
| BATF | Hispanic | 0.044 | -0.123 | 0.210 |
| BATF | Other | 0.037 | -0.223 | 0.288 |
| BATF | Unknown | -0.192 | -0.422 | 0.038 |
| BATF | Male | -0.123 | -0.209 | -0.039 |
| BATF | Viral | -0.810 | -0.943 | -0.677 |
| BATF | Noninfected | -0.808 | -0.948 | -0.666 |
| BATF | GPL570 | 0.342 | -0.165 | 0.851 |
| BATF | GPL6947 | 0.006 | -0.445 | 0.459 |
| BATF | GPL6244 | 0.091 | -0.911 | 1.171 |
| BATF | GPL13667 | 0.335 | -0.490 | 1.134 |
| BATF | GPL20844 | -0.331 | -1.391 | 0.687 |
| BATF | GPL6102 | -0.247 | -1.254 | 0.758 |
| BATF | GPL10332 | 0.431 | -0.536 | 1.422 |
| BATF | NanoString | 0.071 | -0.308 | 0.450 |
| BATF | Age:Viral | 0.184 | 0.095 | 0.273 |
| BATF | Age:Noninfected | -0.160 | -0.258 | -0.055 |
| BATF | Male:Viral | 0.082 | -0.043 | 0.202 |
| BATF | Male:Noninfected | 0.071 | -0.067 | 0.208 |
| BATF | Asian:Viral | -0.246 | -0.624 | 0.129 |
| BATF | Black or African descent:Viral | -0.090 | -0.327 | 0.147 |
| BATF | Hispanic:Viral | -0.089 | -0.317 | 0.141 |
| BATF | Other:Viral | -0.029 | -0.363 | 0.302 |
| BATF | Unknown:Viral | 0.514 | 0.160 | 0.866 |
| BATF | Asian:Noninfected | 0.093 | -0.271 | 0.467 |
| BATF | Black or African descent:Noninfected | 0.029 | -0.221 | 0.276 |
| BATF | Hispanic:Noninfected | 0.167 | -0.045 | 0.376 |
| BATF | Other:Noninfected | 0.081 | -0.234 | 0.395 |
| BATF | Unknown:Noninfected | 0.277 | 0.039 | 0.510 |
| ISG15 | Intercept | -0.466 | -0.688 | -0.234 |
| ISG15 | Age | -0.174 | -0.264 | -0.080 |
| ISG15 | Asian | 0.249 | -0.009 | 0.517 |
| ISG15 | Black or African descent | -0.312 | -0.461 | -0.160 |
| ISG15 | Hispanic | -0.213 | -0.360 | -0.065 |
| ISG15 | Other | 0.013 | -0.219 | 0.238 |
| ISG15 | Unknown | 0.048 | -0.150 | 0.242 |
| ISG15 | Male | -0.056 | -0.152 | 0.033 |
| ISG15 | Viral | 1.207 | 1.086 | 1.323 |
| ISG15 | Noninfected | 0.202 | 0.071 | 0.334 |
| ISG15 | GPL570 | 0.091 | -0.288 | 0.480 |
| ISG15 | GPL6947 | 0.221 | -0.145 | 0.577 |
| ISG15 | GPL6244 | -0.574 | -1.411 | 0.235 |
| ISG15 | GPL13667 | 0.141 | -0.467 | 0.752 |
| ISG15 | GPL20844 | 0.238 | -0.530 | 1.022 |
| ISG15 | GPL6102 | 0.077 | -0.690 | 0.831 |
| ISG15 | GPL10332 | -0.423 | -1.121 | 0.299 |
| ISG15 | NanoString | 0.235 | -0.052 | 0.538 |
| ISG15 | Age:Viral | 0.047 | -0.028 | 0.122 |
| ISG15 | Age:Noninfected | -0.074 | -0.167 | 0.017 |
| ISG15 | Male:Viral | -0.025 | -0.142 | 0.097 |
| ISG15 | Male:Noninfected | -0.062 | -0.197 | 0.072 |
| ISG15 | Asian:Viral | -0.083 | -0.406 | 0.238 |

**Supplementary Table 14 – continued from previous page**

| Marker | Effect | Post. Median | 95% Post. CI LB | 95% Post. CI UB |
|---|---|---|---|---|
| ISG15 | Black or African descent:Viral | 0.287 | 0.076 | 0.493 |
| ISG15 | Hispanic:Viral | 0.433 | 0.236 | 0.628 |
| ISG15 | Other:Viral | 0.044 | -0.260 | 0.346 |
| ISG15 | Unknown:Viral | -0.098 | -0.407 | 0.195 |
| ISG15 | Asian:Noninfected | -0.207 | -0.535 | 0.116 |
| ISG15 | Black or African descent:Noninfected | 0.337 | 0.120 | 0.549 |
| ISG15 | Hispanic:Noninfected | 0.242 | 0.061 | 0.434 |
| ISG15 | Other:Noninfected | 0.071 | -0.214 | 0.347 |
| ISG15 | Unknown:Noninfected | -0.033 | -0.239 | 0.169 |
| KIAA1370 | Intercept | -0.407 | -0.741 | -0.057 |
| KIAA1370 | Age | -0.102 | -0.247 | 0.038 |
| KIAA1370 | Asian | 0.096 | -0.207 | 0.413 |
| KIAA1370 | Black or African descent | 0.163 | -0.006 | 0.336 |
| KIAA1370 | Hispanic | 0.176 | 0.014 | 0.344 |
| KIAA1370 | Other | 0.033 | -0.222 | 0.294 |
| KIAA1370 | Unknown | 0.020 | -0.217 | 0.255 |
| KIAA1370 | Male | -0.112 | -0.197 | -0.027 |
| KIAA1370 | Viral | 0.244 | 0.115 | 0.375 |
| KIAA1370 | Noninfected | 0.418 | 0.280 | 0.560 |
| KIAA1370 | GPL570 | 0.114 | -0.571 | 0.792 |
| KIAA1370 | GPL6947 | 0.026 | -0.525 | 0.571 |
| KIAA1370 | GPL6244 | 0.520 | -0.616 | 1.721 |
| KIAA1370 | GPL13667 | 0.028 | -0.909 | 0.992 |
| KIAA1370 | GPL20844 | -0.202 | -1.451 | 0.928 |
| KIAA1370 | GPL6102 | 0.495 | -0.642 | 1.701 |
| KIAA1370 | GPL10332 | 0.539 | -0.631 | 1.748 |
| KIAA1370 | NanoString | 0.487 | -0.004 | 0.945 |
| KIAA1370 | Age:Viral | -0.092 | -0.184 | -0.001 |
| KIAA1370 | Age:Noninfected | 0.063 | -0.042 | 0.167 |
| KIAA1370 | Male:Viral | 0.003 | -0.116 | 0.132 |
| KIAA1370 | Male:Noninfected | 0.050 | -0.083 | 0.182 |
| KIAA1370 | Asian:Viral | 0.060 | -0.333 | 0.455 |
| KIAA1370 | Black or African descent:Viral | 0.026 | -0.213 | 0.259 |
| KIAA1370 | Hispanic:Viral | -0.028 | -0.255 | 0.195 |
| KIAA1370 | Other:Viral | 0.143 | -0.194 | 0.490 |
| KIAA1370 | Unknown:Viral | 0.389 | 0.030 | 0.759 |
| KIAA1370 | Asian:Noninfected | 0.137 | -0.249 | 0.516 |
| KIAA1370 | Black or African descent:Noninfected | -0.003 | -0.263 | 0.241 |
| KIAA1370 | Hispanic:Noninfected | -0.056 | -0.263 | 0.154 |
| KIAA1370 | Other:Noninfected | -0.199 | -0.522 | 0.122 |
| KIAA1370 | Unknown:Noninfected | 0.010 | -0.227 | 0.251 |
| JUP | Intercept | -0.534 | -0.729 | -0.336 |
| JUP | Age | -0.242 | -0.327 | -0.161 |
| JUP | Asian | -0.027 | -0.306 | 0.246 |
| JUP | Black or African descent | 0.072 | -0.092 | 0.234 |
| JUP | Hispanic | -0.063 | -0.216 | 0.093 |
| JUP | Other | 0.160 | -0.082 | 0.398 |
| JUP | Unknown | 0.175 | -0.040 | 0.387 |
| JUP | Male | -0.110 | -0.196 | -0.026 |
| JUP | Viral | 1.177 | 1.052 | 1.300 |
| JUP | Noninfected | 0.333 | 0.200 | 0.462 |
| JUP | GPL570 | 0.414 | 0.062 | 0.772 |
| JUP | GPL6947 | 0.217 | -0.087 | 0.501 |
| JUP | GPL6244 | -0.155 | -0.961 | 0.666 |
| JUP | GPL13667 | 0.015 | -0.534 | 0.591 |

**Supplementary Table 14 – continued from previous page**

| Marker | Effect | Post. Median | 95% Post. CI LB | 95% Post. CI UB |
|---|---|---|---|---|
| JUP | GPL20844 | 0.031 | -0.739 | 0.870 |
| JUP | GPL6102 | 0.190 | -0.568 | 0.934 |
| JUP | GPL10332 | -0.141 | -0.889 | 0.576 |
| JUP | NanoString | 0.025 | -0.243 | 0.294 |
| JUP | Age:Viral | 0.140 | 0.059 | 0.221 |
| JUP | Age:Noninfected | 0.112 | 0.019 | 0.205 |
| JUP | Male:Viral | 0.029 | -0.094 | 0.149 |
| JUP | Male:Noninfected | 0.050 | -0.084 | 0.183 |
| JUP | Asian:Viral | 0.140 | -0.192 | 0.478 |
| JUP | Black or African descent:Viral | 0.011 | -0.216 | 0.231 |
| JUP | Hispanic:Viral | 0.159 | -0.054 | 0.373 |
| JUP | Other:Viral | -0.097 | -0.423 | 0.221 |
| JUP | Unknown:Viral | -0.103 | -0.420 | 0.210 |
| JUP | Asian:Noninfected | -0.112 | -0.445 | 0.233 |
| JUP | Black or African descent:Noninfected | 0.109 | -0.126 | 0.342 |
| JUP | Hispanic:Noninfected | -0.072 | -0.267 | 0.122 |
| JUP | Other:Noninfected | -0.057 | -0.354 | 0.237 |
| JUP | Unknown:Noninfected | -0.246 | -0.462 | -0.029 |
| PSMB9 | Intercept | -0.199 | -0.453 | 0.044 |
| PSMB9 | Age | -0.022 | -0.160 | 0.117 |
| PSMB9 | Asian | 0.332 | 0.019 | 0.639 |
| PSMB9 | Black or African descent | -0.176 | -0.351 | -0.005 |
| PSMB9 | Hispanic | 0.017 | -0.153 | 0.188 |
| PSMB9 | Other | 0.118 | -0.140 | 0.382 |
| PSMB9 | Unknown | -0.241 | -0.475 | -0.009 |
| PSMB9 | Male | -0.124 | -0.210 | -0.037 |
| PSMB9 | Viral | 0.656 | 0.521 | 0.786 |
| PSMB9 | Noninfected | 0.039 | -0.104 | 0.186 |
| PSMB9 | GPL570 | -0.359 | -0.804 | 0.082 |
| PSMB9 | GPL6947 | 0.053 | -0.309 | 0.434 |
| PSMB9 | GPL6244 | 0.012 | -0.956 | 1.001 |
| PSMB9 | GPL13667 | -0.356 | -1.088 | 0.391 |
| PSMB9 | GPL20844 | 0.552 | -0.399 | 1.565 |
| PSMB9 | GPL6102 | 0.378 | -0.568 | 1.362 |
| PSMB9 | GPL10332 | -0.417 | -1.330 | 0.448 |
| PSMB9 | NanoString | 0.517 | 0.185 | 0.852 |
| PSMB9 | Age:Viral | 0.054 | -0.039 | 0.147 |
| PSMB9 | Age:Noninfected | -0.220 | -0.328 | -0.113 |
| PSMB9 | Male:Viral | 0.021 | -0.108 | 0.143 |
| PSMB9 | Male:Noninfected | 0.030 | -0.106 | 0.168 |
| PSMB9 | Asian:Viral | -0.210 | -0.575 | 0.167 |
| PSMB9 | Black or African descent:Viral | 0.209 | -0.031 | 0.453 |
| PSMB9 | Hispanic:Viral | 0.200 | -0.038 | 0.437 |
| PSMB9 | Other:Viral | -0.030 | -0.390 | 0.328 |
| PSMB9 | Unknown:Viral | 0.188 | -0.180 | 0.553 |
| PSMB9 | Asian:Noninfected | -0.037 | -0.407 | 0.354 |
| PSMB9 | Black or African descent:Noninfected | 0.292 | 0.040 | 0.550 |
| PSMB9 | Hispanic:Noninfected | 0.178 | -0.038 | 0.395 |
| PSMB9 | Other:Noninfected | 0.135 | -0.202 | 0.460 |
| PSMB9 | Unknown:Noninfected | 0.034 | -0.202 | 0.279 |
| C3AR1 | Intercept | 0.464 | 0.196 | 0.738 |
| C3AR1 | Age | 0.042 | -0.081 | 0.156 |
| C3AR1 | Asian | -0.120 | -0.450 | 0.209 |
| C3AR1 | Black or African descent | -0.364 | -0.552 | -0.183 |
| C3AR1 | Hispanic | -0.412 | -0.594 | -0.223 |

**Supplementary Table 14 – continued from previous page**

| Marker | Effect | Post. Median | 95% Post. CI LB | 95% Post. CI UB |
|---|---|---|---|---|
| C3AR1 | Other | -0.262 | -0.556 | 0.021 |
| C3AR1 | Unknown | -0.303 | -0.546 | -0.061 |
| C3AR1 | Male | -0.022 | -0.121 | 0.082 |
| C3AR1 | Viral | -0.231 | -0.385 | -0.082 |
| C3AR1 | Noninfected | -0.790 | -0.946 | -0.631 |
| C3AR1 | GPL570 | 0.234 | -0.223 | 0.684 |
| C3AR1 | GPL6947 | -0.149 | -0.520 | 0.258 |
| C3AR1 | GPL6244 | -0.413 | -1.425 | 0.533 |
| C3AR1 | GPL13667 | -0.184 | -0.948 | 0.617 |
| C3AR1 | GPL20844 | -0.487 | -1.484 | 0.465 |
| C3AR1 | GPL6102 | -0.109 | -1.044 | 0.790 |
| C3AR1 | GPL10332 | 0.213 | -0.639 | 1.094 |
| C3AR1 | NanoString | -0.270 | -0.627 | 0.079 |
| C3AR1 | Age:Viral | 0.073 | -0.021 | 0.169 |
| C3AR1 | Age:Noninfected | -0.002 | -0.113 | 0.115 |
| C3AR1 | Male:Viral | 0.057 | -0.089 | 0.199 |
| C3AR1 | Male:Noninfected | 0.121 | -0.038 | 0.278 |
| C3AR1 | Asian:Viral | -0.112 | -0.512 | 0.287 |
| C3AR1 | Black or African descent:Viral | 0.165 | -0.100 | 0.421 |
| C3AR1 | Hispanic:Viral | 0.132 | -0.122 | 0.380 |
| C3AR1 | Other:Viral | 0.148 | -0.232 | 0.529 |
| C3AR1 | Unknown:Viral | -0.013 | -0.397 | 0.372 |
| C3AR1 | Asian:Noninfected | -0.387 | -0.790 | 0.018 |
| C3AR1 | Black or African descent:Noninfected | 0.148 | -0.121 | 0.416 |
| C3AR1 | Hispanic:Noninfected | 0.134 | -0.093 | 0.360 |
| C3AR1 | Other:Noninfected | 0.262 | -0.100 | 0.622 |
| C3AR1 | Unknown:Noninfected | 0.042 | -0.215 | 0.300 |
| S100A12 | Intercept | 0.248 | -0.018 | 0.508 |
| S100A12 | Age | 0.169 | 0.046 | 0.294 |
| S100A12 | Asian | -0.017 | -0.309 | 0.288 |
| S100A12 | Black or African descent | -0.061 | -0.231 | 0.105 |
| S100A12 | Hispanic | -0.022 | -0.183 | 0.148 |
| S100A12 | Other | 0.116 | -0.145 | 0.376 |
| S100A12 | Unknown | -0.010 | -0.242 | 0.220 |
| S100A12 | Male | -0.028 | -0.112 | 0.059 |
| S100A12 | Viral | -0.590 | -0.725 | -0.455 |
| S100A12 | Noninfected | -0.508 | -0.645 | -0.362 |
| S100A12 | GPL570 | 0.238 | -0.205 | 0.687 |
| S100A12 | GPL6947 | -0.319 | -0.738 | 0.088 |
| S100A12 | GPL6244 | -0.225 | -1.191 | 0.660 |
| S100A12 | GPL13667 | 0.323 | -0.396 | 1.052 |
| S100A12 | GPL20844 | -0.157 | -1.046 | 0.732 |
| S100A12 | GPL6102 | -0.182 | -1.061 | 0.673 |
| S100A12 | GPL10332 | 0.491 | -0.334 | 1.366 |
| S100A12 | NanoString | 0.123 | -0.225 | 0.467 |
| S100A12 | Age:Viral | -0.213 | -0.302 | -0.127 |
| S100A12 | Age:Noninfected | -0.171 | -0.272 | -0.067 |
| S100A12 | Male:Viral | 0.058 | -0.071 | 0.181 |
| S100A12 | Male:Noninfected | 0.119 | -0.018 | 0.251 |
| S100A12 | Asian:Viral | -0.144 | -0.519 | 0.219 |
| S100A12 | Black or African descent:Viral | 0.118 | -0.107 | 0.355 |
| S100A12 | Hispanic:Viral | 0.046 | -0.183 | 0.268 |
| S100A12 | Other:Viral | -0.235 | -0.581 | 0.116 |
| S100A12 | Unknown:Viral | -0.002 | -0.363 | 0.354 |
| S100A12 | Asian:Noninfected | 0.218 | -0.152 | 0.581 |

| Marker | Effect | Post. Median | 95% Post. CI LB | 95% Post. CI UB |
|---|---|---|---|---|
| S100A12 | Black or African descent:Noninfected | -0.101 | -0.343 | 0.151 |
| S100A12 | Hispanic:Noninfected | 0.127 | -0.085 | 0.340 |
| S100A12 | Other:Noninfected | 0.057 | -0.258 | 0.369 |
| S100A12 | Unknown:Noninfected | 0.129 | -0.107 | 0.364 |
| LY86 | Intercept | -0.247 | -0.491 | -0.006 |
| LY86 | Age | -0.172 | -0.266 | -0.079 |
| LY86 | Asian | -0.080 | -0.398 | 0.243 |
| LY86 | Black or African descent | 0.058 | -0.122 | 0.240 |
| LY86 | Hispanic | 0.055 | -0.119 | 0.236 |
| LY86 | Other | -0.207 | -0.478 | 0.065 |
| LY86 | Unknown | -0.121 | -0.366 | 0.123 |
| LY86 | Male | 0.047 | -0.057 | 0.152 |
| LY86 | Viral | 0.532 | 0.384 | 0.679 |
| LY86 | Noninfected | 0.401 | 0.254 | 0.548 |
| LY86 | GPL570 | -0.199 | -0.618 | 0.220 |
| LY86 | GPL6947 | -0.180 | -0.564 | 0.174 |
| LY86 | GPL6244 | -0.602 | -1.616 | 0.334 |
| LY86 | GPL13667 | -0.327 | -0.951 | 0.345 |
| LY86 | GPL20844 | 0.126 | -0.732 | 1.009 |
| LY86 | GPL6102 | -1.096 | -2.003 | -0.181 |
| LY86 | GPL10332 | 0.228 | -0.585 | 1.042 |
| LY86 | NanoString | 0.127 | -0.221 | 0.473 |
| LY86 | Age:Viral | 0.100 | 0.009 | 0.193 |
| LY86 | Age:Noninfected | 0.124 | 0.015 | 0.235 |
| LY86 | Male:Viral | 0.026 | -0.125 | 0.182 |
| LY86 | Male:Noninfected | 0.046 | -0.108 | 0.198 |
| LY86 | Asian:Viral | 0.155 | -0.259 | 0.549 |
| LY86 | Black or African descent:Viral | -0.089 | -0.331 | 0.164 |
| LY86 | Hispanic:Viral | -0.199 | -0.442 | 0.041 |
| LY86 | Other:Viral | 0.219 | -0.153 | 0.590 |
| LY86 | Unknown:Viral | -0.172 | -0.540 | 0.194 |
| LY86 | Asian:Noninfected | -0.092 | -0.500 | 0.289 |
| LY86 | Black or African descent:Noninfected | -0.320 | -0.582 | -0.065 |
| LY86 | Hispanic:Noninfected | -0.336 | -0.563 | -0.112 |
| LY86 | Other:Noninfected | 0.102 | -0.240 | 0.447 |
| LY86 | Unknown:Noninfected | 0.090 | -0.166 | 0.345 |
| HK3 | Intercept | 0.536 | 0.247 | 0.818 |
| HK3 | Age | 0.104 | -0.010 | 0.220 |
| HK3 | Asian | 0.172 | -0.126 | 0.484 |
| HK3 | Black or African descent | -0.008 | -0.170 | 0.157 |
| HK3 | Hispanic | -0.003 | -0.166 | 0.163 |
| HK3 | Other | 0.127 | -0.122 | 0.381 |
| HK3 | Unknown | -0.056 | -0.284 | 0.165 |
| HK3 | Male | 0.019 | -0.066 | 0.111 |
| HK3 | Viral | -0.733 | -0.865 | -0.600 |
| HK3 | Noninfected | -0.702 | -0.841 | -0.557 |
| HK3 | GPL570 | 0.580 | 0.060 | 1.087 |
| HK3 | GPL6947 | -0.125 | -0.548 | 0.297 |
| HK3 | GPL6244 | -0.076 | -1.164 | 0.995 |
| HK3 | GPL13667 | 0.114 | -0.637 | 0.874 |
| HK3 | GPL20844 | -0.531 | -1.647 | 0.554 |
| HK3 | GPL6102 | -0.585 | -1.693 | 0.408 |
| HK3 | GPL10332 | 0.501 | -0.413 | 1.518 |
| HK3 | NanoString | -0.338 | -0.746 | 0.065 |
| HK3 | Age:Viral | 0.109 | 0.020 | 0.197 |

**Supplementary Table 14 – continued from previous page**

| Marker | Effect | Post. Median | 95% Post. CI LB | 95% Post. CI UB |
|---|---|---|---|---|
| HK3 | Age:Noninfected | -0.226 | -0.329 | -0.125 |
| HK3 | Male:Viral | 0.067 | -0.064 | 0.191 |
| HK3 | Male:Noninfected | 0.090 | -0.046 | 0.223 |
| HK3 | Asian:Viral | -0.292 | -0.664 | 0.077 |
| HK3 | Black or African descent:Viral | -0.152 | -0.384 | 0.072 |
| HK3 | Hispanic:Viral | -0.110 | -0.346 | 0.117 |
| HK3 | Other:Viral | -0.165 | -0.511 | 0.177 |
| HK3 | Unknown:Viral | -0.310 | -0.664 | 0.043 |
| HK3 | Asian:Noninfected | 0.034 | -0.338 | 0.402 |
| HK3 | Black or African descent:Noninfected | -0.096 | -0.334 | 0.145 |
| HK3 | Hispanic:Noninfected | 0.172 | -0.045 | 0.380 |
| HK3 | Other:Noninfected | -0.063 | -0.381 | 0.251 |
| HK3 | Unknown:Noninfected | -0.230 | -0.457 | -0.006 |
